# Modeling the health and economic impact of scaling up monthly oral pre-exposure prophylaxis alone or alongside injectable Lenacapavir in Kenya and South Africa

**DOI:** 10.64898/2026.09.01.26361983

**Authors:** Akash Malhotra, Nishali Patel, David Kaftan, Edinah Mudimu, Anna Bershteyn, Monisha Sharma

**Affiliations:** Department of Global Health, University of Washington, Seattle, WA, USA; Department of Health Metrics Sciences, University of Washington, Seattle, WA, USA; Department of Population Health, New York University Grossman School of Medicine, New York, NY, USA; Department of Decision Sciences, University of South Africa, Pretoria, South Africa

**Keywords:** Pre-exposure prophylaxis, long acting, lenacapavir, monthly oral, MK-8527, HIV, Africa, modeling, cost-effectiveness

## Abstract

**Introduction:** Monthly oral HIV pre-exposure prophylaxis (PrEP) such as MK-8527 offer promise as low-cost, self-administered options that are easily delivered through community-based platforms. Economic evaluations of MK-8527 either alone or alongside other long-acting (LA) PrEP products like lenacapavir are needed for informing HIV prevention strategies.

**Methods:** We adapted an agent-based network model, EMOD-HIV, to simulate LA-PrEP scale-up in South Africa and western Kenya from 2026-2035; scenarios evaluated MK-8527 alone, lenacapavir alone and combined strategies, with varying uptake among female sex workers, their male clients, and individuals with >1 partner. We assumed 95% effectiveness of MK-8527 for 2 months (assuming individuals took 2 of 3 pills dispensed) and 95% lenacapavir effectiveness for 6 months. Scenarios were compared to a baseline of daily oral PrEP only.

**Results:** Assuming the same uptake rates, MK-8527 alone achieved lower health impacts than lenacapavir alone in both settings (6-14% vs. 11-18% of HIV infections averted) but had substantially lower costs; provision costs of MK-8527 were 58-59% lower than lenacapavir assuming $1.00/pill and 40–43% lower at $2.50/pill. In western Kenya, ICERs for MK-8527 alone were $467/DALY averted and $799/DALY averted assuming pill prices of $1.00 and $2.50 respectively, compared to $1,306/DALY averted for lenacapavir. In South Africa, all LA-PrEP strategies were cost-saving over the 35-year horizon, although near-term budget impacts were substantial ($169–348 million over five years). Service delivery accounted for the majority of MK-8527 costs (73% at $1.00 per pill). Combined strategies of lenacapavir and MK-8527 increased health benefits (13-23% infections averted) but had higher provision costs than either strategy alone.

**Conclusion:** MK-8527 can reduce HIV incidence at lower costs than lenacapavir. However, high service delivery costs limit its cost-effectiveness to scenarios in which pill prices are low (US$1.00 per pill) and provision is targeted to populations at substantial HIV risk.

## INTRODUCTION

HIV remains a leading cause of morbidity and mortality, disproportionately impacting Eastern and Southern Africa (ESA)^1^, the global region with the highest HIV incidence. While daily oral pre-exposure prophylaxis (PrEP) is effective at preventing HIV, uptake, adherence and continuation are low in ESA; reported barriers include pill burden and stigma of taking daily antiretrovirals. Studies among persons with HIV risk indication find a preference for long-acting (LA) PrEP which is perceived as a convenient and discreet alternative to daily pill taking.^2^ Further, while the effectiveness of daily oral PrEP is contingent on an individual’s adherence, LA-PrEP can achieve high long-term effectiveness after one dose. Six-monthly injectable lenacapavir is a promising LA-PrEP product, demonstrating nearly 100% efficacy in clinical trials^3^. However, the impact of LA-PrEP is dependent on equitable and affordable distribution to those at greatest HIV risk. Modeling studies suggest that focused lenacapavir scale-up to those at substantial HIV risk can avert 12-18% of HIV infections and is cost-effective in western Kenya and South Africa at a maximum price per dose of approximately $17 and $106 USD, respectively^4^. However focused lenacapavir scale-up alone is likely insufficient to reach goals for epidemic control, and broadening coverage to those at moderate HIV risk reduces lenacapavir’s maximum price threshold that achieves cost-effectiveness. Recent price negotiations have lowered lenacapavir to $20 per dose (plus a $17 loading dose), which is considerably lower than the price threshold estimated for South Africa but higher than that of Kenya (under focused coverage scenarios)^4,5^. Additionally, injectable LA PrEP is not universally preferred among those who could benefit, with some individuals reporting fear of injections and side effects^6^. Lenacapavir provision requires trained providers to administer injections which may present logistical challenges in differentiated delivery settings. To maximize the benefits of lenacapavir and achieve wider PrEP coverage, multiple HIV prevention options should be considered.

Monthly oral PrEP formulations such as MK-8527 offer promise as a low-cost, self-administered pill that is less burdensome to health systems. Since MK-8527 avoids injection-related logistics (trained providers, sharps, observation time, etc) it can be more easily integrated within community-based distribution channels. Although clinical effectiveness results from Phase 3 clinical trials are pending, Phase 2 data indicate favorable safety with protective intracellular drug levels reached within 24 hours and persisting for >28 days, pharmacology that matches a monthly schedule^7^. If effective, MK-8527 can be a scalable LA-PrEP strategy to increase coverage of biomedical HIV prevention. The cost-effectiveness of MK-8527 depends on coverage and continuation among those with HIV risk, price, and scale up of other PrEP products. We sought to evaluate plausible scenarios of MK-8527 scale-up either alone or alongside lenacapavir in South Africa and western Kenya. The model-projected health and economic impact of MK-8527 can identify implementation conditions that increase likelihood of cost-effective scale-up.

## METHODS

### Mathematical model

We adapted EMOD-HIV, an existing network-based dynamic model developed by Institute for Disease Modeling^8,9^ to simulate scale up of MK-8527 and lenacapavir. EMOD-HIV is an open-source model that incorporates population demography, HIV natural history, and heterosexual HIV transmission, configured to match age- and sex-specific propensities of forming sexual partnerships. The model uses monthly time steps and accounts for the impact of HIV treatment and prevention on the epidemic. The modeled HIV care continuum includes HIV testing, linkage and retention on antiretroviral therapy (ART). The PrEP continuum includes uptake, adherence, persistence, and re-engagement. The model tracks outcomes including HIV infections, HIV-related deaths, and healthcare use, enabling calculation of disability adjusted life years (DALYs) and HIV-related costs.

We parameterized the model with epidemiological data from South Africa (estimated HIV prevalence ages 15-49 in 2023: 17.1%)^10^, and western Kenya (11.3%) including age-specific fertility, mortality, voluntary male circumcision coverage, oral PrEP uptake, health-seeking behavior, population size by age/sex, and sizes of key populations such as female sex workers (FSWs) and male clients of FSWs ^11,12^. The model was calibrated to empiric data on age and sex-specific HIV prevalence and number of individuals on ART using an algorithm that maximizes the likelihood of matching observed data. We selected 100 good-fitting model parameter sets to conduct the analyses. See Appendix for parameterization and calibration data (pages 3-34).

### Modelled scenarios

In the baseline (no LA-PrEP) scenario, we assumed availability of only daily oral PrEP, used by individuals aged 15-65 years with HIV risk indication and scaled to currently reported coverages in both countries^13^. We assumed oral PrEP decreased the risk of HIV acquisition by 75% based on clinical trials accounting for adherence; individuals persisted on PrEP for an average of 3-months based on data from implementation studies^14^. In LA-PrEP scenarios we modeled the scale-up of MK-8527 either alone or in combination with lenacapavir; LA-PrEP was introduced in 2026 and scaled linearly to target coverage by 2029. We assumed LA-PrEP provision ends in 2035 to evaluate the impact of a ten-year implementation program. We utilized a 35-year analytic time horizon (2026-2060) to assess long-term health outcomes and calculate cost-effectiveness.

Based on clinical trials, we assumed lenacapavir reduced HIV acquisition risk by 95% for a 6-month duration^3^. In the absence of empirical data on MK-8527 effectiveness, we assumed effectiveness equivalent to that of oral PrEP when used with high adherence, which is found to reduce HIV acquisition by >95%^15^. Because the monthly oral formulation does not require daily adherence, we assumed 95% effectiveness for each 30-day period in which a pill was taken. MK-8527 was modeled to be dispensed in 3-month supplies; on average, individuals were assumed to take the first two pills. Accordingly, we assumed MK-8527 reduced HIV acquisition risk by 95% for the first two months, with no protection in the third month.

We estimated plausible scenarios of LA-PrEP uptake using previously developed assumptions based on review of the PrEP preferences literature^6^. Lenacapavir uptakes estimates by subgroup were consistent with our previous analysis^4^: FSWs (40%), male clients of FSWs (40%), adolescent girls and young women (AGYW) age 16-24 years with >1 partner (32%), women age 25-50 years with >1 partner (36%), and men age 25-50 years with >1 partner (32%) (Table 2). In the “MK-8527 only” scenario, we utilized the same uptake rates as lenacapavir, assuming persons seeking LA PrEP would utilize MK-8527 in the absence of lenacapavir availability. For all scenarios modeling both lenacapavir and MK-8527, we assumed lenacapavir uptake remained the same as the “LEN only” scenario and the impact of MK-8527 provision was largely additive. This aligns with the family planning literature which demonstrates an increase in total contraceptive coverage with introduction of new methods^16^. Further, studies find a strong preference for injectables and longer duration products among those at HIV risk so we assume most individuals would choose lenacapavir over MK-8527 if both products were available. Therefore, lenacapavir uptake is substantially higher than MK-8527 in scenarios modeling both products. In the “MK-8527 + LEN” scenario, we assume MK-8527 uptake rates increase the total LA-PrEP uptake to the midpoint of our previously estimated plausible uptake range, leading to MK-8527 uptake by subgroup of: FSWs (17%), male clients of FSWs (17%), adolescent girls and young women (AGYW) age 16-24 years with >1 partner (14%), women age 25-50 years with >1 partner (15%), and men age 25-50 years with >1 partner (14%). Due to uncertainty regarding MK-8527 uptake, we evaluated additional MK-8527 scenarios. In the “MK-8527 only at higher uptake” scenario, we assumed MK-8527 availability would expand coverage to equal that of the “MK-8527 + lenacapavir” scenario; therefore the uptake rates by subgroup were calculated as the combined uptake of both products in the “MK-8527 + LEN” scenario. In the “Expanded MK-8527 + LEN” scenario, we utilized the same MK-8527 uptake rates by subgroup as the “MK-8527 + LEN” scenario and expanded MK-8527 uptake to those with >1 partner in the last 3 months (but not concurrent) at the same sex-specific rates used for persons with concurrent partners in the single product scenarios. Similarly, in the “expanded MK-8527 only” scenario, we assessed expanded MK-8527 coverage to individuals with >1 partner in the last 3 months. Across scenarios, persons who discontinued LA PrEP were eligible to re-initiate if they met eligibility criteria. Uptake assumptions are summarized in Table 2.

### Sensitivity analyses

In addition to varying MK-8527 coverage, we also evaluated the impact of potential future PEPFAR funding disruptions. For Kenya, we modeled a scenario in which both HIV testing and ART coverage was reduced by 41%, based on assumptions utilized in a previous modeling analysis (PEPFAR accounts for 41% of Kenya’s HIV budget)^17^. For South Africa, we assumed HIV testing and ART were reduced by 18% (PEPFAR accounts for 18% of South Africa’s HIV budget)^17^. We also evaluated pessimistic MK-8527 scenarios assuming persons took only 1 of the three pills dispensed.

### Model outcomes

For LA PrEP scenarios, we estimated number of HIV infections, HIV-related deaths, DALYs and percentage of each outcome averted compared to the counterfactual scenario of oral PrEP availability only. We calculated population PrEP coverage as the total person months on PrEP divided by the total person months in the analysis period. We assumed 6 months of coverage for lenacapavir and 2 months for MK-8527 (assuming individuals took an average of two out of three pills dispensed). We calculated 90% uncertainty intervals across 100 parameter sets. Model outputs were analyzed in R version 4.2.2.

### Costs

Costs included HIV testing, ART, HIV-related hospitalizations, oral and LA-PrEP provision, and LA-PrEP product wastage (Table 1; Appendix pages 36-39). Lenacapavir drug prices were based on recent agreements enabling reduced cost generic Lenacapavir drugs for low- and middle-income countries (LMICs) ($20 per 6-monthly injection and $17 oral loading dose)^5^. We assumed 75% lenacapavir continuation among those eligible; loading dose costs were only incurred at lenacapavir initiation. As the price of MK-8527 has not yet been established, we based our estimates on expert consultation; we assumed MK-8527 would be available at $1 per pill but also evaluated an upper bound estimate of $2.50 per pill. LA-PrEP provision costs included personnel, HIV testing, and overhead. We included costs for demand generation, assumed to be 10% of the lenacapavir drug price (with the same cost assumed for MK-8527). We assumed individuals incurred a cost for a 3-month supply of MK-8527 and took an average of 2 pills, resulting in wastage of the 3^rd^ pill.

**Table 1.** Model parameters, assumptions, and cost inputs^￥^.

| Model assumptions | Value | Source |
| --- | --- | --- |
| Oral PrEP effectiveness | 75% | Baeten 2012<sup>14</sup> |
| Lenacapavir effectiveness | 95% | Bekker 2024<sup>22</sup> |
| MK-8527 effectiveness | 95% | Assumption based on oral PrEP effectiveness with high adherence |
| LA PrEP scaleup period | 2026 – 2029 | Assumption |
| LA PrEP implementation period | 2026 – 2035 | Assumption |
| Analytic time horizon | 2026 – 2060 | Assumption |
| **Costs (USD)** |  |  |
| MK-8527 (per pill) | \$1.00<br>(\$2.50, \$5.00) | Assumption9/1/2026 8:47:00 PM |
| MK-8527 provision | \$7.20 | Wanga et al. 20215, Mangale et al. 20256, Bhardwaj (unpublished)9/1/2026 8:47:00 PM |
| MK-8527 wastage | 10% | Assumption9/1/2026 8:47:00 PM |
| Lenacapavir dose (6 monthly) | \$20.00 | CHAI 2025 |
| Lenacapavir oral loading dose | \$17.50 | CHAI 2025 |
| Lenacapavir wastage | 5% | Assumption |
| LA PrEP demand generation | 10% of LEN drug cost | Assumption based on VMMC literature (same cost for MK-8527 & lenacapavir) |
| **western Kenya** |  |  |
| Annual health care costs (among those not on ART) |  |  |
| HIV-positive CD4 < 200 | \$110.30 | Eaton 2014<sup>23</sup> |
| HIV-positive CD4 200 - 349 | \$30.38 | Eaton 2014<sup>23</sup> |
| HIV-positive CD4 > 350 | \$8.59 | Eaton 2014<sup>23</sup> |
| End of life care | \$105.68 | Eaton 2014<sup>23</sup> |
| ART provision (annual) | \$196.85 | Larson 2018<sup>24</sup>, Long 2010<sup>25</sup>, The Global Fund<sup>26</sup>, and Haas 2015<sup>27</sup> |
| Oral PrEP per person month | \$10.88 | Wanga 2019<sup>28</sup> |
| Facility-based HIV-positive test | \$3.68 | Meisner 2021<sup>29</sup> 9/1/2026 8:47:00 PM |
| Facility-based HIV-negative test | \$2.64 | Meisner 2021<sup>29</sup> |
| **South Africa** |  |  |
| Annual health care costs (among those not on ART) |  |  |
| HIV-positive CD4 < 200 | \$374.08 | Eaton 2014<sup>23</sup> |
| HIV-positive CD4 200 - 349 | \$102.95 | Eaton 2014<sup>23</sup> |
| HIV-positive CD4 > 350 | \$29.10 | Eaton 2014<sup>23</sup> |
| End of life care | \$358.10 | Eaton 2014<sup>23</sup> |
| ART provision (annual) | \$189.56 | Larson 2018<sup>24</sup>, Long 2010<sup>25</sup>, The Global Fund<sup>26</sup>, and Haas 2015<sup>27</sup> |
| Oral PrEP per person month | \$15.20 | 30,31 |
| Facility-based HIV-positive test | \$5.62 | Meyer-Rath 2019<sup>32</sup> |
| Facility-based HIV-negative test | \$3.62 | Meyer-Rath 2019<sup>32</sup> |

<sup>‡</sup> PrEP: pre-exposure prophylaxis, ART: antiretroviral therapy. VMMC: voluntary medical male circumcision. Costs are adjusted for inflation and GDP per capita ratio where applicable. ART costs are of delivery costs and assume 3% of patients receive 2nd-line ART. Lenacapavir demand generation costs are adapted from the literature for voluntary medical male circumcision demand generation. Additional details on costing methods are available in Appendix.

**Table 2:** Long-acting PrEP uptake among subgroups by scenario*.

|  | LEN only |  | LEN + MK-8527 |  | LEN + Expanded MK-8527 |  | MK-8527 only |  | MK-8527 only at higher uptake |  | Expanded MK-8527 only |  |
| --- | --- | --- | --- | --- | --- | --- | --- | --- | --- | --- | --- | --- |
|  | LEN | MK-8527 | LEN | MK-8527 | LEN | MK-8527 | LEN | MK-8527 | LEN | MK-8527 | LEN | MK-8527 |
| **FSWs** | 40% | - | 40% | 17% | 40% | 17% | - | 40% | - | 57% | - | 48% |
| **Male clients of FSWs** | 40% | - | 40% | 17% | 40% | 17% | - | 40% | - | 57% | - | 48% |
| **AGYW** | 32% | - | 32% | 14% | 32% | 14% | - | 32% | - | 46% | - | 39% |
| **Women with concurrent partners** | 36% | - | 36% | 15% | 36% | 15% | - | 36% | - | 51% | - | 44% |
| **Men with concurrent partners** | 32% | - | 32% | 14% | 32% | 14% | - | 32% | - | 46% | - | 39% |
| **Women with >1 partner in last 3 months\*** | - | - | - | - | - | 36% | - | - | - | - | - | 36% |
| **Men with >1 partner in last 3 months\*** | - | - | - | - | - | 32% | - | - | - | - | - | 32% |
| **Women whose partners have other partners** | - | - | - | - | - | 36% | - | - | - | - | - | 36% |

\* LA PrEP: long acting PrEP; LEN: lenacapavir. \*Partners not concurrent

### Economic evaluation

Using the payer perspective, we calculated the incremental cost-effectiveness ratio as the difference in costs divided by the difference in DALYs for each LA PrEP scenario compared to the counterfactual scenario of oral PrEP only. We utilized a commonly referenced supply-side cost-effectiveness threshold of $500 USD per DALY averted^18^. We also estimated the undiscounted five-year budget impact of LA-PrEP scale-up from the healthcare payer perspective, disaggregated by cost category.

### Ethics and consent

Data used for this analysis were from publicly available sources, therefore did not require informed consent nor ethical approval.

## RESULTS

### Western Kenya

In western Kenya, “LEN only” scale-up at 2.4% population coverage was estimated to avert 18.0% of HIV infections and 4.4% of HIV-related deaths and had an ICER of $1,306/DALY averted (Table 3A). Scaling up both LA-PrEP products together (“LEN + MK-8527”), increased health benefits to 23.2% of HIV infections and 5.6% of HIV-related deaths averted but was associated with a higher ICER than “LEN only”: $1,412 and $1,489/DALY averted at an MK-8527 price of $1.00 and $2.50, respectively. “LEN + Expanded MK-8527,” which entailed broadening MK-8527 provision to individuals with lower HIV risk, increased health benefits slightly compared to “LEN + MK-8527” but had a higher ICER: $1,542 and $1,710, respectively at MK-8527 pill prices of $1.00 and $2.50. Compared to “LEN only”, scale-up of “MK-8527 only” at the same uptake resulted in lower population coverage (1.3%) and infections averted (13.9%) but had a considerably lower ICER: $467 and $799/DALY averted at MK-8527 pill prices of $1.00 and $2.50, respectively. In the “MK-8527 only at higher uptake” which assumed higher LA-PrEP coverage among those at highest risk, HIV infections averted were slightly lower than the “LEN only” scenario (17.2% vs 18.4%), but the ICER was considerably lower: $748/DALY averted. The “Expanded MK-8527 only” scenario achieved similar health benefits as “LEN only” (18.1% of HIV infections and 5.2% of HIV-related deaths averted) and a lower ICER at an MK-8527 pill price of $1.00: $887/DALY averted; at a higher pill price of $2.50, the ICER exceeded that of “LEN only:” $1,436/DALY averted.

**Table 3A:**
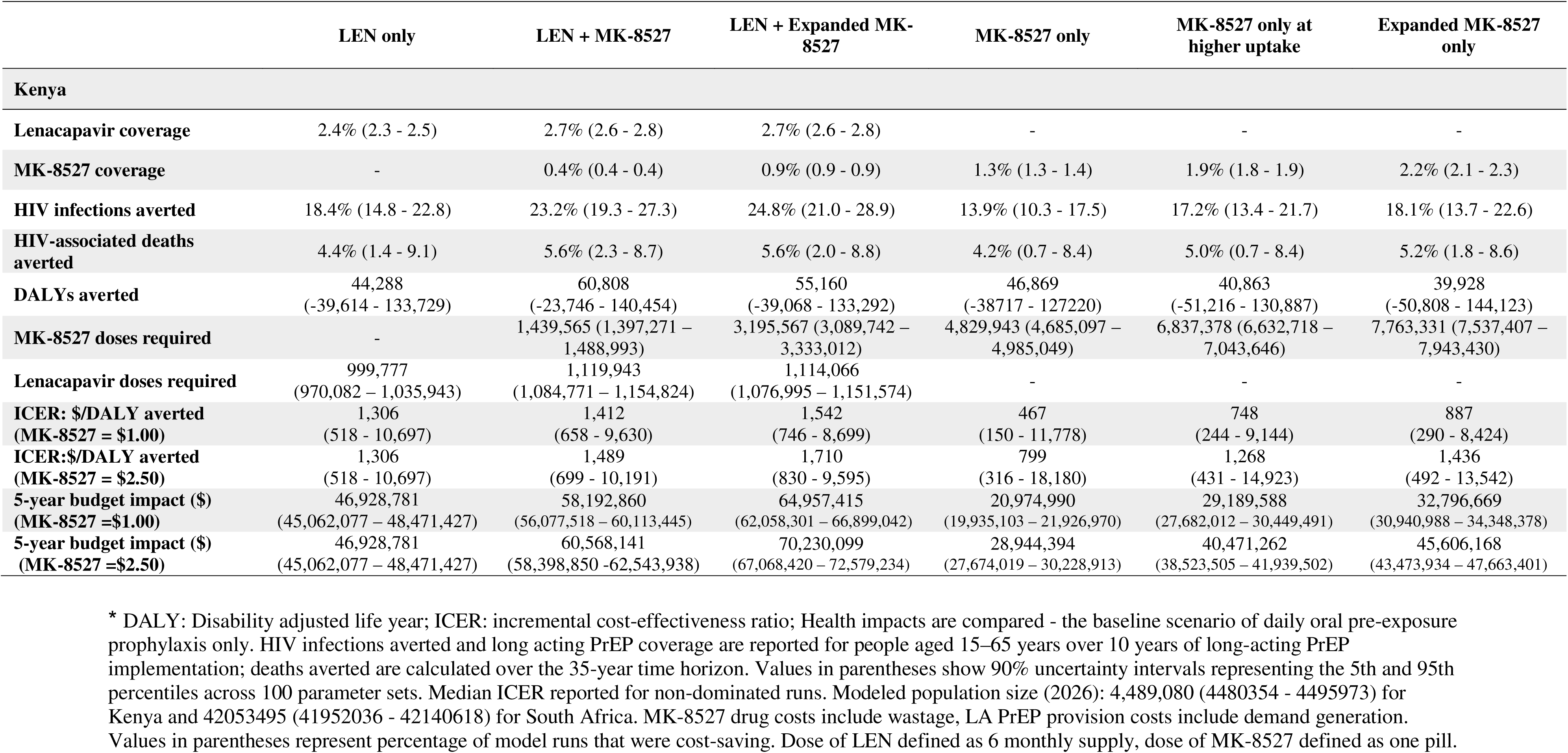
Health and economic impact of scenarios of LA PrEP scale up in western Kenya*.

At an MK-8527 price of $1.00/pill, the budget impact of all scenarios of MK-8527 alone were lower than those containing lenacapavir; for example, the “LEN only” scenario had a 5-year budget impact of $46.9 million while “Expanded MK-8527 only” had similar health benefits at a budget impact of $30.5 million and “MK-8527 only at higher uptake” had slightly lower health benefits at a budget impact of $34.5 million. Scenarios with both LA-PrEP products had the highest budget impact (e.g. $59.0 million for the “LEN + MK-8527” scenario). At a higher MK-8527 price of $2.50/pill, the budget impact of most scenarios of MK-8527 alone were lower than “LEN only” except for the “Expanded MK-8527 only” which was slightly higher ($47.4 million). Cost savings due to HIV-related illness accounted for <5% of total incremental costs across scenarios) (Tables S1 – S5, Appendix pages 46 - 56). LA-PrEP drugs and delivery made up the largest portion of the budget impact (>90%).

**Figure 1a** shows the total 5-year costs of LA PrEP provision (drugs and service delivery) by strategy in Kenya at both MK-8527 prices. For the “LEN only” scenario, the majority (74%) of the 46 million in program costs were due to drugs while delivery (including personnel, overhead, HIV testing, and demand generation) accounted for 26%. Similarly, for scenarios with both products, drugs remain the major cost driver. The opposite pattern is observed for scenarios of MK-8527 alone (assuming $1.00/pill); drug costs make up only 27% of total costs while delivery accounts for 73%. For example, in the “MK-8527 only” scenario, while drug costs were relatively low (5.3 million), service delivery costs were substantial (14.2 million). At a higher price of $2.50/pill, drugs accounted for nearly half of total LA-PrEP provision costs for scenarios of MK-8527 alone (48%) while service delivery costs remained the same as the lower priced MK-8527 scenarios and now comprised 52% of total costs.

**Figure 1.**
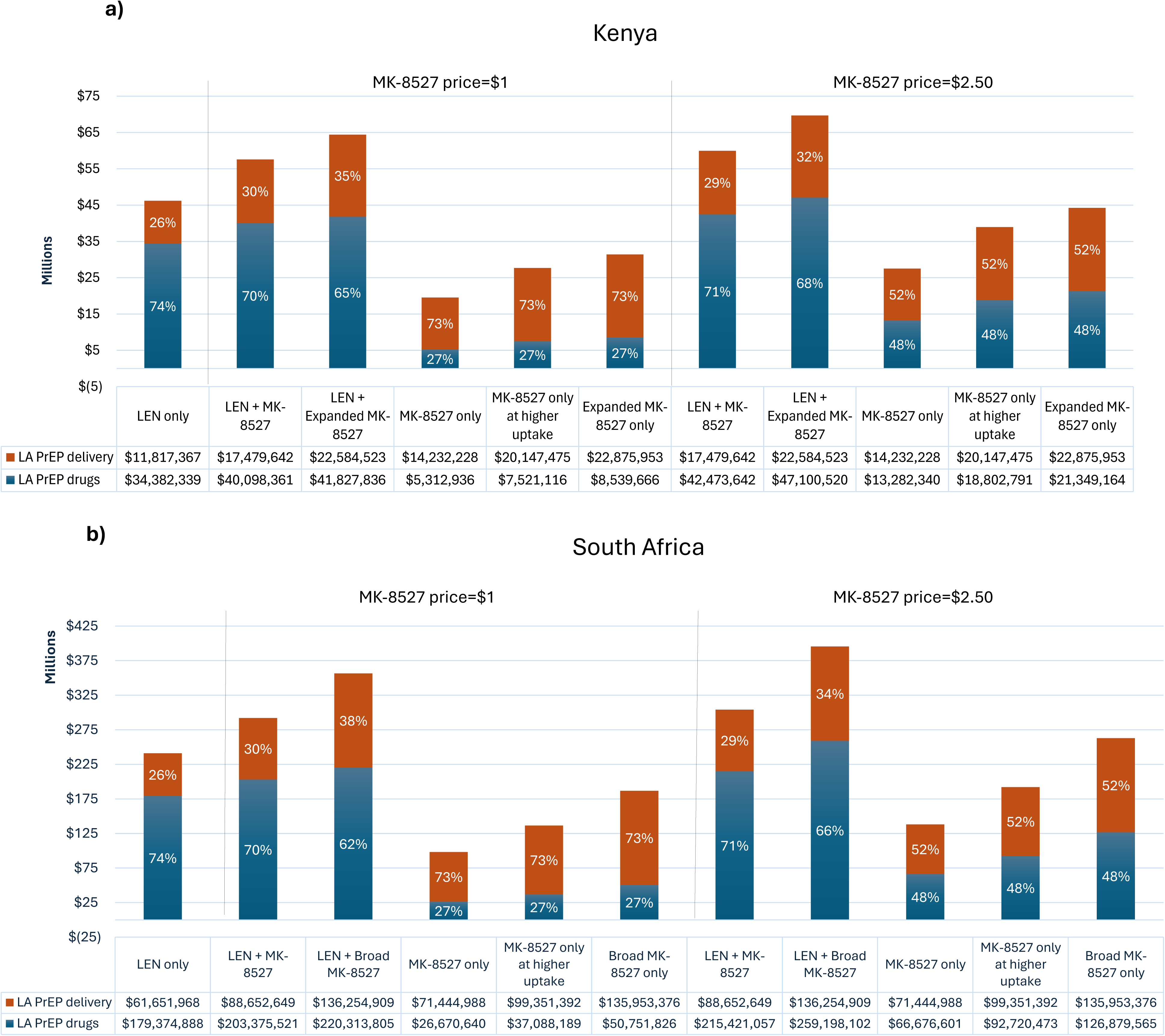

### South Africa

The relative health impacts of LA-PrEP scenarios in South Africa was similar to that of western Kenya, with strategies containing lenacapavir having the highest health benefits at the highest costs; however, all ICERs were cost-saving (Table 3B). “LEN only” scale-up at 1.4% population coverage averted 10.7% of HIV infections and 2.5% of HIV-related deaths with a budget impact of $240.5 million; the “LEN + MK-8527” scenario, increased health benefits (13.2% of HIV infections and 3.1% of HIV-related deaths averted) but had a higher budget impact: $311.4 million and $323.4 million at an MK-8527 price of $1.00 and $2.50, respectively. “LEN + Expanded MK-8527,” had slightly higher health benefits than “LEN + MK-8527” but at a higher budget impact: $393.3 million and $432.2 million, respectively at MK-8527 price/pill of $1.00 and $2.50. While “MK-8527 only” had lower health benefits than “LEN only” (13.9%) it had a significantly lower budget impact: $178 million and $218 million, at MK-8527 pill prices of $1.00 and $2.50, respectively. Both the “MK-8527 only at higher uptake” and the “Expanded MK-8527” scenarios had slightly lower health benefits than “LEN only” but at lower costs: 183 million and 240 million, respectively at MK-8527 pill prices of $1.00 and 183 million and 240 million at $2.50. Similar to Kenya, the total 5-year costs of LA-PrEP provision (drugs and service delivery) for LEN only strategies were largely comprised of PrEP drugs (74%) while the majority of MK-8527 only costs at $1.00/pill were due to service delivery (73%) **(Figure 1b)**. At $2.50/pill, PrEP provision costs for MK-8527 only strategies were equally comprised of service delivery (52%) and drugs (48%).

**Table 3B:**
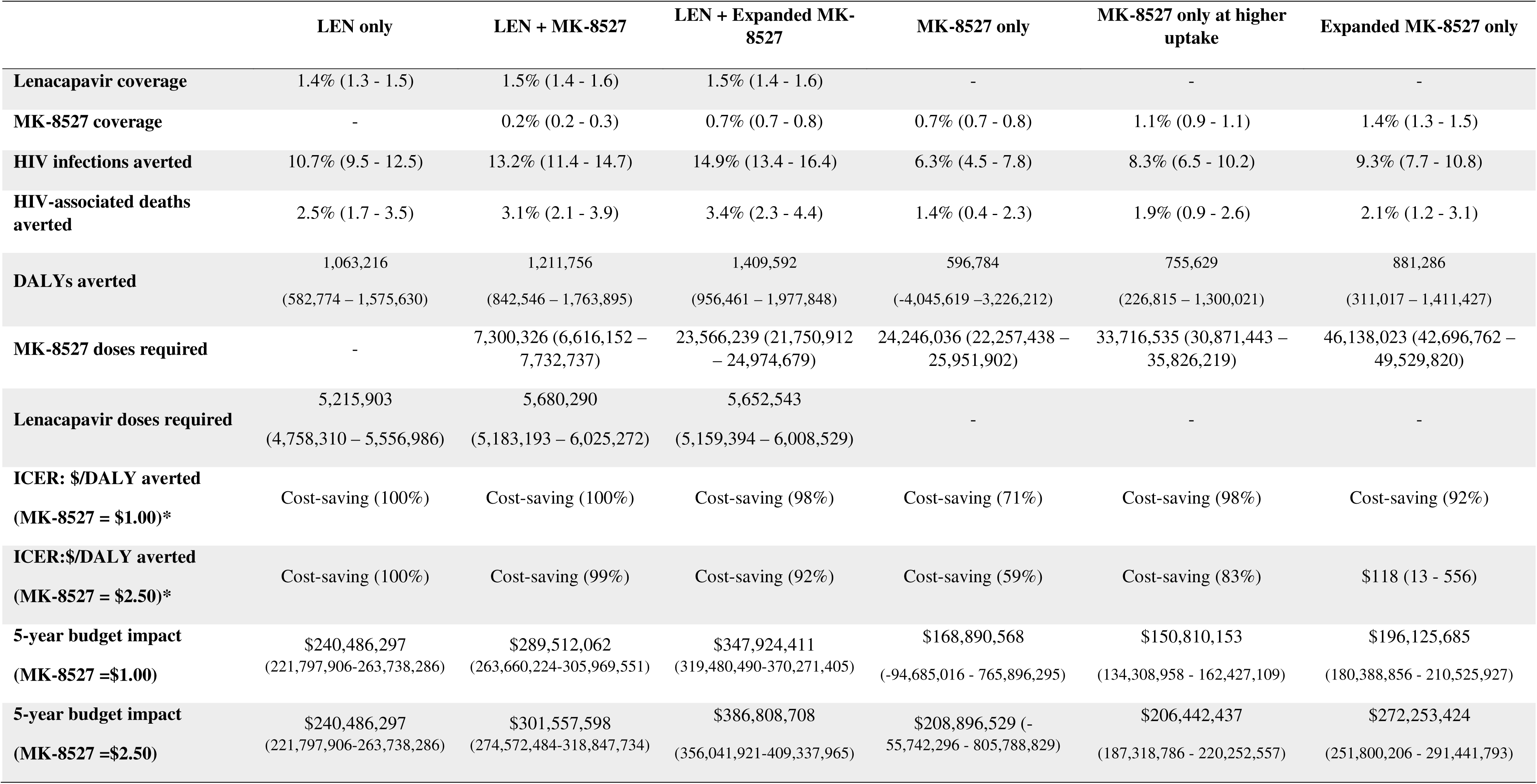
Health and economic impact of scenarios of LA PrEP scale up in South Africa*.

### Sensitivity analyses

In PEPFAR funding reduction scenarios, all LA-PrEP strategies resulted in higher HIV infections and deaths averted and had a smaller budget impact due to greater ART costs averted. In Kenya, LA PrEP strategies averted 9-13-times more DALYs than the main scenario; ICERs for strategies of MK-8527 alone were cost-saving and those containing lenacapavir had ICERs of $31-67 per DALY averted. Similarly, for South Africa, LA-PrEP averted nearly 2 times more DALYs than the main analysis and all scenarios remained cost-saving. The 5-year budget impact ranged from $21-34 million for strategies of MK-8527 at $1.00/pill and $29-37 million at $2.50/pill; lenacapavir strategies had a budget impact of $46-71 million.

Pessimistically assuming individuals using MK-8527 take only 1 of the 3 pills dispensed slightly lowered HIV infections averted for MK-8527 only scenarios to 12-16% in Kenya and 5-7% in South Africa. In Kenya, “MK-8527 only” ICER increased to $549 and $864 at MK-8527 prices of $1.00 and $2.50/pill respectively. ICERs for “MK-8527 only at higher uptake” and “Expanded MK-8527 only” were $876 and $814/DALY averted assuming $1.00 and $2.50/pill respectively; at $2.50/pill ICERs increased substantially to $1,288 and $1,423/DALY averted respectively. In South Africa, MK-8527 only strategies remained cost-saving at $1.00/pill (aside from “Expanded MK-8527 only,” ICER: $119/DALY averted); at $2.50/pill ICERs ranged from $152/DALY averted for “MK-8527 only” to $317/DALY averted for “Expanded MK-8527 only.” At higher prices of $5.00 per pill MK-8527 strategies were more costly and less effective than lenacapavir alon

## DISCUSSION

In this model-based analysis, we assessed the impact of scaling up MK-8527 in western Kenya and South Africa, either alone or alongside lenacapavir. Overall, we find adding MK-8527 to lenacapavir provision results in greater HIV infections and deaths averted compared to lenacapavir alone, but at a higher budget impact. These results reflect the assumption that providing multiple options for HIV prevention can increase overall PrEP coverage and associated health benefits by aligning with diverse user preferences. In qualitative studies among individuals at risk for HIV, participants emphasize the value of being able to choose the LA-PrEP formulation that best fits their needs and preferences^6^. Although injectable LA-PrEP has high acceptability, some individuals report fear of injections or safety concerns about a long-duration product and prefer oral formulations. Further, MK-8527 may be a low-cost option that can be more feasibly delivered through community-based channels by lower cadre providers. We find differences in health benefits and cost-effectiveness by setting, highlighting the importance of country-specific analyses. In western Kenya, ICERs for combined LA-PrEP strategies were higher than those of lenacapavir alone, while in South Africa, all LA-PrEP strategies were cost-saving, reflecting the higher disease burden and greater downstream costs averted.

Due to budget constraints, large-scale rollout of lenacapavir at 1.5-2.5% population coverage, either alone or with MK-8527, may be cost-prohibitive in some LMICs. Policymakers may consider offering limited lenacapavir provision (e.g. for pregnant women) and focusing efforts on scaling up MK-8527 distribution. When assessing strategies of MK-8527 only, we find that both strategies of “MK-8527 only” and “MK-8527 at higher uptake” provide slightly lower health benefits than “LEN only,” (eg 6.3-8.3% of infections averted vs 10.7% in “LEN only” in South Africa), but at substantially lower costs. Across settings, provision costs of both MK-8527 strategies were 41-60% lower than that of “LEN only” at $1.00/pill and 17-41% times lower times lower assuming $2.50/pill. Even in settings such as South Africa, where lenacapavir was found to be cost-saving over a 35-year time horizon, it would require high upfront provision costs ($240 million over five years) and would not be cost-saving in the near term. In an era of shrinking HIV funding, countries may prioritize maintaining high coverage of lifesaving antiretroviral therapy (ART) and allocate fewer resources to HIV prevention. In this context, MK-8527 may provide a feasible low-cost strategy to reduce HIV incidence.

In a scenario of “expanded of MK-8527 only”, we project similar health benefits and considerably lower costs than lenacapavir alone if pill prices are low ($1.00/pill). However, at a higher price of $2.50/pill, costs of expanded MK-8527 provision exceeded that of lenacapavir alone in both settings, suggesting that expanded MK-8527 distribution is likely efficient only at low MK-8527 prices. However, the feasibility of achieving expanded MK-8527 provision remains uncertain. MK-8527 required significantly higher modeled initiation rates among priority populations to achieve the same coverage as lenacapavir due to the lower assumed two months of protection (2 pills taken out of 3 dispensed), vs. six months for lenacapavir. The demand generation required to achieve these higher initiation rates may be substantial, particularly in light of the low uptake of oral PrEP in ESA. Additionally, MK-8527 strategies were projected to require over 7.5-9 times as many doses as lenacapavir to achieve the same coverage. Investments in personnel and infrastructure are likely required to dispense the volume of pills, frequency of visits, and adherence support associated with expanded provision. Indeed, we find that even at low costs ($1.00/pill), expanded MK-8527 may not be cost-effective in all settings. In Kenya, the ICER for expanded MK-8527 at $1.00/pill was $887/DALY averted, exceeded the commonly cited threshold of $500/DALY averted (although it was more efficient than lenacapavir alone, ICER: 1,306/DALY averted). This finding was primarily driven by delivery costs which comprised 73% of total MK-8527 provision costs; therefore even at low MK-8527 pill prices, the relatively high cost of personnel, infrastructure, 3-monthly dispensing, HIV testing, etc., may limit the cost-effectiveness of broadening MK-8527 scale-up. We conservatively assumed lower PrEP service delivery costs than some reported in the literature, reflecting provision through community-based channels such as pharmacies. Using higher published PrEP delivery cost estimates (e.g., $13–25 vs. $8.55 in our analysis^19,20^) would increase ICERs for MK-8527 and further reduce the efficiency of expanded MK-8527 implementation, particularly compared to lenacapavir, which requires half the number of delivery visits. This suggests that strategies to lower service delivery costs (e.g., 6-monthly dispensing with HIV self-tests) may have a greater impact on cost-effectiveness than reductions in product costs. Additionally, our expanded MK-8527 scenarios still assume targeted uptake among those with substantial HIV risk; broadening provision to those with lower HIV risk would likely further reduce cost-effectiveness, suggesting that widespread MK-8527 use is likely not cost-effective, even with a low-cost product, due to high service delivery costs.

Our analysis has several limitations. First, due to a lack of empirical data, we relied on assumptions and expert opinion to parameterize MK-8527 effectiveness, price, and pill-taking behavior. We assumed imperfect adherence in our main analysis (2 pills out of 3 taken) to account for real-world adherence challenges and varied this assumption in sensitivity analyses. In a pessimistic scenario (only 1 of 3 pills taken), overall conclusions were similar, although health benefits were reduced and ICERs were less attractive. These findings highlight the importance of adherence support to maximize duration of protection and minimize pill wastage. To address price uncertainty, we evaluated a range of plausible prices and found that MK-8527 would have to be low cost in order to be a cost-effective complement or replacement for lenacapavir. Second, LA-PrEP uptake assumptions were informed by the PrEP preference literature rather than observed use under routine implementation. However, we evaluated multiple plausible scenarios, varying uptake, product combinations, and priority populations. Third, our model only simulates heterosexual mixing, which accounts for much of the HIV epidemic in ESA; therefore, we cannot assess the impact of LA-PrEP among men who have sex with men or persons who inject drugs, both important populations for LA-PrEP. Additionally, we do not model pregnant women, who have high HIV incidence in ESA and can benefit from LA-PrEP. Fifth, we adopted a payer perspective which does not capture participant costs incurred or averted by LA-PrEP.

Strengths of this analysis include using an individual-based network model and evaluating uncertainty across 100 parameter sets in two geographic regions of ESA. We report both long-term health outcomes and cost-effectiveness and short-term provision costs. To our knowledge this is the first modeling study to evaluate the impact of MK-8527 scale-up alongside lenacapavir in ESA and the first to utilize data on PrEP preferences across subgroups to inform MK-8527 uptake. Our results are in line with a previous analysis, which finds that targeted MK-8527 scale-up at 5% coverage is cost-effective at monthly MK-8527 costs of $0.87 and $5.02 in western Kenya and South Africa, respectively^21^.

## CONCLUSION

Overall, we found that MK-8527 provision, either alone or alongside lenacapavir, can avert substantial HIV incidence and may be cost-effective in ESA if pill prices are low. Our findings can inform policy decisions regarding scale-up of novel LA PrEP products.

## Data Availability

All data produced in the present study are available upon reasonable request to the authors

## ACKNOWLEDGEMENTS

This study was funded by the Gates Foundation. The funders had no role in study design, data collection, analysis, writing of the report, nor the decision to submit for publication. We gratefully acknowledge the Institute for Disease Modeling (Seattle, WA, USA) for developing and maintaining EMOD-HIV, particularly Dan Bridenbecker and Clark Kirkman IV who provided technical and computational support.

## DATA SHARING

Data used for the modeling can be found in the Appendix. EMOD-HIV is open-source and publicly available: https://docs.idmod.org/projects/emod-hiv/en/2.20_a/.

## AUTHOR CONTRIBUTIONS

All authors contributed to conceptualization of the modeling question, provided substantive input into the modelling scenarios and assumptions, interpreted results, and critically reviewed manuscript drafts. All authors contributed to model development, and coding. AM and NP conducted analysis of model outputs and accessed and verified the data. MS wrote the first draft of the manuscript. All authors read and approved the final manuscript.

