## Supplemental appendix for "Modeling the health and economic impact of scaling up monthly oral pre-exposure prophylaxis alone or alongside injectable Lenacapavir in Kenya and South Africa"

**Accompanying the manuscript:**

#### Empiric Data for Model Calibration and Validation

##### Table S1: HIV incidence by region

| Setting | Incidence per 1000  Adults aged 15 – 49 |
| --- | --- |
| Nyanza, Kenya | 2.34 [1.90 – 3.08] |
| South Africa | 7.79 [4.58 - 10.80] |

##### HIV prevalence in counties by age and sex

###### Table S2a. HIV prevalence in counties of the former Nyanza province (Homa Bay, Kisii, Kisumu, Migori, Nyamira, Siaya), by age and sex in Kenya^¥^

| **Age group (years)** | **2003** | | **2007** | | **2008** | | **2012** | | **2018** | |
| --- | --- | --- | --- | --- | --- | --- | --- | --- | --- | --- |
|  | **Men** | **Women** | **Men** | **Women** | **Men** | **Women** | **Men** | **Women** | **Men** | **Women** |
| **15 - 19** | 0.0015 | 0.0459 | 0.0121 | 0.0773 | 0.0184 | 0.1078 | 0.0151 | 0.0486 | 0.0025 | 0.0294 |
| **20 - 24** | 0.0562 | 0.2997 | 0.0257 | 0.2056 | 0.0578 | 0.1201 | 0.0289 | 0.1411 | 0.0216 | 0.0910 |
| **25 - 29** | 0.2429 | 0.2301 | 0.1956 | 0.2454 | 0.2450 | 0.2228 | 0.2107 | 0.2454 | 0.0705 | 0.2334 |
| **30 - 34** | 0.1840 | 0.1632 | 0.2578 | 0.2576 | 0.1530 | 0.2593 | 0.2392 | 0.2047 | 0.0911 | 0.2696 |
| **35- 39** | 0.2064 | 0.1838 | 0.2384 | 0.2227 | 0.2275 | 0.2259 | 0.1955 | 0.2811 | 0.1675 | 0.2705 |
| **40 - 44** | 0.2533 | 0.3500 | 0.2024 | 0.1799 | 0.2501 | 0.0927 | 0.3132 | 0.1694 | 0.2075 | 0.2765 |
| **45 - 49** | 0.1624 | 0.1651 | 0.2103 | 0.1291 | 0.1331 | 0.1716 | 0.1623 | 0.2287 | 0.2796 | 0.1991 |
| **15 - 49** | 0.1160 | 0.1830 | 0.1140 | 0.1760 | 0.1140 | 0.1600 | 0.1340 | 0.1760 | 0.0826 | 0.1667 |

**^¥^Sources:** Kenya Demographic and Health Surveys, 2003 & 2008; Kenya AIDS Indicator Surveys, 2007 & 2012; Kenya Population-Based HIV Impact Assessment 2018.

###### Table S2b. HIV prevalence data by age and sex from population-based surveys for model calibration in South Africa^¥^

| **Sex** | **Age group**  **(years)** | **Year** | | | | |
| --- | --- | --- | --- | --- | --- | --- |
|  |  | **2002** | **2005** | **2008** | **2012** | **2017** |
| **Men** | 15-19 | 0.040 | 0.032 | 0.025 | 0.007 | 0.047 |
|  | 20-24 | 0.080 | 0.060 | 0.051 | 0.051 | 0.048 |
|  | 25-29 | 0.220 | 0.121 | 0.157 | 0.173 | 0.124 |
|  | 30-34 | 0.240 | 0.233 | 0.258 | 0.256 | 0.184 |
|  | 35-39 | 0.180 | 0.233 | 0.185 | 0.288 | 0.238 |
|  | 40-44 | 0.120 | 0.175 | 0.192 | 0.158 | 0.224 |
|  | 45-49 | 0.120 | 0.103 | 0.084 | 0.134 | 0.248 |
|  | 50-54 | 0.050 | 0.142 | 0.104 | 0.155 | 0.202 |
|  | 55-59 | 0.070 | 0.064 | 0.062 | 0.055 | 0.148 |
|  | 15-49 | 0.128 | 0.117 | 0.116 | 0.145 | 0.148 |
| **Women** | 15-19 | 0.070 | 0.094 | 0.067 | 0.056 | 0.058 |
|  | 20-24 | 0.170 | 0.239 | 0.211 | 0.174 | 0.156 |
|  | 25-29 | 0.320 | 0.333 | 0.327 | 0.284 | 0.275 |
|  | 30-34 | 0.240 | 0.260 | 0.291 | 0.360 | 0.347 |
|  | 35-39 | 0.140 | 0.193 | 0.248 | 0.316 | 0.394 |
|  | 40-44 | 0.190 | 0.124 | 0.163 | 0.280 | 0.359 |
|  | 45-49 | 0.110 | 0.087 | 0.141 | 0.197 | 0.303 |
|  | 50-54 | 0.080 | 0.075 | 0.102 | 0.148 | 0.222 |
|  | 55-59 | 0.070 | 0.030 | 0.077 | 0.097 | 0.176 |
|  | 15-49 | 0.177 | 0.202 | 0.213 | 0.232 | 0.263 |

^¥^Sources: South African National HIV Prevalence, Incidence and Behaviour Surveys (2002, 2005, 2008, 2012 and 2017) from the Human Sciences Research Council (HSRC)

###### Table S3. HIV prevalence among men and women ages 15-49, by county and sex in Kenya^¥^

| **County** | **2003** | | **2007** | | **2008** | | **2012** | | **2018** | |
| --- | --- | --- | --- | --- | --- | --- | --- | --- | --- | --- |
|  | **Men** | **Women** | **Men** | **Women** | **Men** | **Women** | **Men** | **Women** | **Men** | **Women** |
| **Homa Bay** | 0.1097 | 0.2458 | 0.2514 | 0.3259 | 0.1737 | 0.2524 | 0.2217 | 0.2787 | 0.1279 | 0.2532 |
| **Kisii** | 0.0114 | 0.0853 | 0.0445 | 0.0693 | 0.0330 | 0.0573 | 0.0346 | 0.0368 | 0.0458 | 0.0684 |
| **Kisumu** | 0.1663 | 0.1914 | 0.1139 | 0.1847 | 0.1109 | 0.1810 | 0.1940 | 0.2022 | 0.0960 | 0.2096 |
| **Migori** | 0.1804 | 0.1860 | 0.1685 | 0.2181 | 0.1923 | 0.2228 | 0.1435 | 0.1925 | 0.0706 | 0.1758 |
| **Nyamira** | 0.0029 | 0.0742 | - | - | 0.0234 | 0.0544 | 0.0419 | 0.1045 | 0.0247 | 0.0432 |
| **Siaya** | 0.1824 | 0.2424 | 0.1445 | 0.2130 | 0.1526 | 0.1921 | 0.2596 | 0.2990 | 0.0961 | 0.1905 |

**^¥^Sources:** Kenya Demographic and Health Surveys, 2003 & 2008; Kenya AIDS Indicator Surveys, 2007 & 2012; Kenya Population-Based HIV Impact Assessment 2018.

##### Number of people on ART

###### Table S4a. Number of people on ART by county, sex, and age group in Kenya^¥^

| **Sex** | **County** | **Age group**  **(years)** | **Year** | | | | | | | | | | | | | |
| --- | --- | --- | --- | --- | --- | --- | --- | --- | --- | --- | --- | --- | --- | --- | --- | --- |
|  |  |  | **2004** | **2005** | **2006** | **2007** | **2008** | **2009** | **2010** | **2011** | **2012** | **2013** | **2014** | **2015** | **2016** | **2017** |
| **Men** | **Homa Bay** | 0 - 14 | - | - | - | - | - | - | - | - | - | - | 2,945 | 3,583 | 4,109 | 4,192 |
|  |  | 15 - 99 | 1,067 | 2,313 | 5,148 | 7,194 | 10,002 | 14,436 | 17,178 | 15,954 | 17,522 | 18,279 | 19,157 | 22,834 | 26,441 | 29,220 |
|  | **Kisii** | 0 - 14 | - | - | - | - | - | - | - | - | - | - | 828 | 993 | 1,109 | 1,083 |
|  |  | 15 - 99 | - | - | - | - | - | - | - | 2,972 | - | - | 4,614 | 5,451 | 6,604 | 7,169 |
|  | **Kisumu** | 0 - 14 | - | - | - | - | - | - | - | - | - | - | 3,101 | 3,245 | 3,525 | 3,607 |
|  |  | 15 - 99 | 945 | 2,047 | 4,557 | 6,368 | 8,853 | 12,779 | 15,206 | 14,122 | 15,511 | 16,180 | 21,216 | 24,550 | 28,082 | 31,021 |
|  | **Migori** | 0 - 14 |  |  |  |  |  |  |  |  |  |  | 2,309 | 2,295 | 2,678 | 2,673 |
|  |  | 15 - 99 | 711 | 1,541 | 3,430 | 4,793 | 6,664 | 9,619 | 11,446 | 10,630 | 11,675 | 12,179 | 13,929 | 15,165 | 17,438 | 18,455 |
|  | **Nyamira** | 0 - 14 | - | - | - | - | - | - | - | - | - | - | 484 | 552 | 578 | 611 |
|  |  | 15 - 99 | - | - | - | - | - | - | - | 1,362 | - | - | 2,120 | 2,585 | 3,142 | 3,474 |
|  | **Siaya** | 0 - 14 | - | - | - | - | - | - | - | - | - | - | 2,645 | 2,950 | 3,017 | 3,197 |
|  |  | 15 - 99 | 860 | 1,864 | 4,148 | 5,797 | 8,060 | 11,633 | 13,843 | 12,856 | 14,120 | 14,730 | 16,163 | 18,611 | 21,477 | 23,762 |
| **Women** | **Homa Bay** | 0 - 14 | - | - | - | - | - | - | - | - | - | - | 3,431 | 3,835 | 4,426 | 4,535 |
|  |  | 15 - 99 | 1,359 | 2,944 | 6,551 | 9,155 | 12,522 | 17,202 | 21,798 | 31,454 | 35,182 | 38,819 | 40,118 | 49,956 | 57,286 | 61,811 |
|  | **Kisii** | 0 - 14 | - | - | - | - | - | - | - | - | - | - | 906 | 1,079 | 1,200 | 1,146 |
|  |  | 15 - 99 | - | - | - | - | - | - | - | 7,902 | - | - | 11,691 | 14,350 | 17,274 | 19,044 |
|  | **Kisumu** | 0 - 14 | - | - | - | - | - | - | - | - | - | - | 3,241 | 3,393 | 3,810 | 3,831 |
|  |  | 15 - 99 | 1,203 | 2,606 | 5,799 | 8,104 | 11,084 | 15,227 | 19,296 | 27,843 | 31,143 | 34,362 | 41,230 | 48,424 | 56,384 | 60,789 |
|  | **Migori** | 0 - 14 | - | - | - | - | - | - | - | - | - | - | 2,526 | 2,448 | 2,868 | 2,884 |
|  |  | 15 - 99 | 905 | 1,961 | 4,365 | 6,100 | 8,343 | 11,461 | 14,524 | 20,958 | 23,442 | 25,865 | 27,896 | 31,964 | 38,637 | 40,891 |
|  | **Nyamira** | 0 - 14 | - | - | - | - | - | - | - | - | - | - | 506 | 567 | 601 | 622 |
|  |  | 15 - 99 | - | - | - | - | - | - | - | 3,766 | - | - | 5,964 | 7,210 | 8,258 | 8,654 |
|  | **Siaya** | 0 - 14 | - | - | - | - | - | - | - | - | - | - | 2,778 | 3,136 | 3,299 | 3,569 |
|  |  | 15 - 99 | 1,095 | 2,372 | 5,279 | 7,377 | 10,090 | 13,862 | 17,566 | 25,347 | 28,351 | 31,281 | 33,911 | 39,853 | 44,892 | 48,808 |
| **Both** | **All** | 15-99 | 8,954 | 19,404 | 43,185 | 60,350 | 83,142 | 116,787 | 143,877 | 175,003 | 194,552 | 210,769 | 246,293 | 301,029 | 333,333 | 389,159 |

**^¥^**Source: Kenya Ministry of Health

###### Table S4b: Number of people on ART by sex (ages 15-49 years) in South Africa ^¥^

| **Year** | **Male** | **Female** |
| --- | --- | --- |
| 2001 | 2713 | 3543 |
| 2002 | 5768 | 7586 |
| 2003 | 9321 | 12313 |
| 2004 | 17717 | 24423 |
| 2005 | 34874 | 59240 |
| 2006 | 66629 | 123308 |
| 2007 | 118977 | 228753 |
| 2008 | 186564 | 367389 |
| 2009 | 277931 | 548206 |
| 2010 | 402000 | 781477 |
| 2011 | 558131 | 1095411 |
| 2012 | 713294 | 1415016 |
| 2013 | 876749 | 1732515 |
| 2014 | 1013499 | 2003014 |
| 2015 | 1130063 | 2239669 |
| 2016 | 1238815 | 2518238 |
| 2017 | 1403702 | 2998170 |

^¥^Source: South Africa Department of Health Surveys

##### Population size by age and sex

###### Table S5a. Population size by sex, county, and age group in 2009 in western Kenya^¥^

| **Age Group**  **(years)** | **Men** | | | | | | **Women** | | | | | |
| --- | --- | --- | --- | --- | --- | --- | --- | --- | --- | --- | --- | --- |
|  | **Homa Bay** | **Kisii** | **Kisumu** | **Migori** | **Nyamira** | **Siaya** | **Homa Bay** | **Kisii** | **Kisumu** | **Migori** | **Nyamira** | **Siaya** |
| **0 - < 1** | 18,335 | 18,236 | 17,457 | 19,265 | 10,313 | 15,093 | 18,354 | 17,993 | 16,926 | 19,309 | 10,263 | 14,860 |
| **1 - 4** | 69,799 | 69,529 | 63,054 | 69,921 | 41,165 | 56,269 | 69,250 | 69,023 | 63,172 | 69,519 | 40,396 | 55,901 |
| **5 - 9** | 75,926 | 76,757 | 67,083 | 73,872 | 46,450 | 60,966 | 75,973 | 75,778 | 67,779 | 74,333 | 46,867 | 60,710 |
| **10 - 14** | 68,689 | 68,473 | 62,706 | 64,300 | 42,590 | 58,296 | 67,159 | 68,072 | 63,359 | 63,249 | 42,198 | 56,248 |
| **15 - 19** | 57,430 | 59,228 | 55,597 | 53,075 | 36,604 | 49,220 | 54,119 | 60,776 | 56,741 | 52,238 | 36,786 | 47,825 |
| **20 - 24** | 39,573 | 41,898 | 47,281 | 38,690 | 24,409 | 32,725 | 50,309 | 58,225 | 57,649 | 48,004 | 34,184 | 41,443 |
| **25 - 29** | 30,437 | 32,792 | 40,964 | 30,727 | 19,515 | 25,961 | 36,016 | 42,878 | 40,614 | 34,670 | 27,273 | 30,135 |
| **30 - 34** | 23,259 | 26,678 | 30,412 | 23,344 | 16,605 | 20,359 | 26,342 | 30,031 | 27,515 | 25,630 | 19,487 | 22,328 |
| **34 - 39** | 16,013 | 21,766 | 21,251 | 17,024 | 14,039 | 14,793 | 20,010 | 26,051 | 20,611 | 19,313 | 17,106 | 17,932 |
| **40 - 44** | 11,914 | 15,718 | 15,145 | 12,170 | 10,470 | 11,118 | 16,513 | 18,360 | 16,894 | 14,773 | 11,377 | 16,082 |
| **45 - 49** | 11,124 | 16,797 | 13,361 | 10,549 | 11,318 | 10,390 | 15,248 | 19,181 | 15,298 | 12,888 | 11,886 | 15,486 |
| **50 - 54** | 9,705 | 12,789 | 11,251 | 8,565 | 8,379 | 9,079 | 12,942 | 14,136 | 12,504 | 10,314 | 8,703 | 14,541 |
| **55 - 59** | 8,159 | 9,527 | 8,718 | 6,399 | 5,999 | 8,414 | 9,833 | 9,528 | 9,175 | 7,692 | 5,819 | 12,265 |
| **60 - 64** | 6,989 | 7,395 | 7,054 | 5,250 | 5,026 | 7,712 | 8,587 | 7,654 | 7,597 | 6,000 | 5,107 | 11,081 |
| **65 - 69** | 4,325 | 4,637 | 4,163 | 3,382 | 3,094 | 5,107 | 5,957 | 5,320 | 5,402 | 4,508 | 3,322 | 7,732 |
| **70 - 74** | 4,029 | 3,945 | 3,777 | 2,907 | 2,753 | 5,175 | 5,355 | 5,017 | 4,757 | 3,524 | 3,153 | 7,173 |
| **75 - 79** | 2,835 | 2,743 | 2,392 | 2,033 | 1,778 | 3,549 | 3,891 | 3,338 | 3,356 | 2,968 | 1,919 | 5,464 |
| **80 - 99** | 3,726 | 3,701 | 2,821 | 2,624 | 2,393 | 4,159 | 5,316 | 5,891 | 4,615 | 3,636 | 3,475 | 6,155 |

**^¥^**Source: Kenya National Bureau of Statistics, 2009 Census

###### Table S5b. Population of South Africa by age and sex ^¥^

| **Sex** | **Age group**  **(years)** | **Year** | | | | |
| --- | --- | --- | --- | --- | --- | --- |
|  |  | **2002** | **2005** | **2008** | **2012** | **2017** |
| **Men** | 0 - 4 | 2634839 | 2651819 | 2692300 | 2784372 | 2886299 |
|  | 5 - 9 | 2559246 | 2570683 | 2592577 | 2644720 | 2767111 |
|  | 10 - 14 | 2582620 | 2565499 | 2557928 | 2578751 | 2636981 |
|  | 15 - 19 | 2546968 | 2596517 | 2584945 | 2572032 | 2587023 |
|  | 20 - 24 | 2348165 | 2488613 | 2572307 | 2606447 | 2594656 |
|  | 25 - 29 | 2069939 | 2215800 | 2362377 | 2530034 | 2612453 |
|  | 30 - 34 | 1759391 | 1899279 | 2018461 | 2227862 | 2488823 |
|  | 35 - 39 | 1488894 | 1568198 | 1664587 | 1845019 | 2131687 |
|  | 40 - 44 | 1270053 | 1324166 | 1367150 | 1493652 | 1731088 |
|  | 45 - 49 | 1077752 | 1122009 | 1159644 | 1234887 | 1398602 |
|  | 50 - 54 | 831061 | 925655 | 973184 | 1038560 | 1147992 |
|  | 55 - 59 | 636945 | 677128 | 760379 | 856728 | 947123 |
| **Women** | 0 - 4 | 2587465 | 2602664 | 2640997 | 2729586 | 2825703 |
|  | 5 - 9 | 2523770 | 2535299 | 2555462 | 2604517 | 2719700 |
|  | 10 - 14 | 2557891 | 2533039 | 2526253 | 2545219 | 2603685 |
|  | 15 - 19 | 2527828 | 2568409 | 2552976 | 2535920 | 2555420 |
|  | 20 - 24 | 2323512 | 2451570 | 2523985 | 2560705 | 2550570 |
|  | 25 - 29 | 2064262 | 2167428 | 2282514 | 2447992 | 2550351 |
|  | 30 - 34 | 1807453 | 1906808 | 1967958 | 2123170 | 2394266 |
|  | 35 - 39 | 1577255 | 1648612 | 1717873 | 1821711 | 2039776 |
|  | 40 - 44 | 1370835 | 1434647 | 1491937 | 1594914 | 1745050 |
|  | 45 - 49 | 1184477 | 1243314 | 1302936 | 1396666 | 1541175 |
|  | 50 - 54 | 924873 | 1054059 | 1124243 | 1214556 | 1348848 |
|  | 55 - 59 | 727478 | 783876 | 898804 | 1035986 | 1160340 |

^¥^Source: United Nations Population Database

##### Voluntary medical male circumcision

###### Table S6a. Circumcision status quo by county, age group (years), and year in Kenya ^¥^

|  | **Homa Bay** | | | **Kisii** | | | **Kisumu** | | | | **Migori** | | | | **Nyamira** | | | | **Siaya** | | |
| --- | --- | --- | --- | --- | --- | --- | --- | --- | --- | --- | --- | --- | --- | --- | --- | --- | --- | --- | --- | --- | --- |
| **Year** | **10-14** | **15-24** | **25-49** | **10-14** | **15-24** | **25-49** | **10-14** | **15-24** | **25-49** | **10-14** | | **15-24** | **25-49** | **10-14** | | **15-24** | **25-49** | **10-14** | | **15-24** | **25-49** |
| **Pre-2008** | 0.249 | 0.249 | 0.249 | 0.948 | 0.948 | 0.948 | 0.322 | 0.322 | 0.322 | 0.410 | | 0.410 | 0.410 | 0.965 | | 0.965 | 0.965 | 0.252 | | 0.252 | 0.252 |
| **2008** | 0.118 | 0.235 | 0.275 | 0.948 | 0.948 | 0.948 | 0.152 | 0.303 | 0.333 | 0.194 | | 0.385 | 0.451 | 0.965 | | 0.965 | 0.965 | 0.119 | | 0.243 | 0.270 |
| **2009** | 0.122 | 0.240 | 0.283 | 0.948 | 0.948 | 0.948 | 0.178 | 0.315 | 0.339 | 0.199 | | 0.388 | 0.463 | 0.965 | | 0.965 | 0.965 | 0.127 | | 0.250 | 0.277 |
| **2010** | 0.166 | 0.285 | 0.299 | 0.948 | 0.948 | 0.948 | 0.324 | 0.389 | 0.362 | 0.214 | | 0.395 | 0.478 | 0.965 | | 0.965 | 0.965 | 0.222 | | 0.310 | 0.293 |
| **2011** | 0.226 | 0.355 | 0.313 | 0.948 | 0.948 | 0.948 | 0.450 | 0.479 | 0.385 | 0.280 | | 0.423 | 0.486 | 0.965 | | 0.965 | 0.965 | 0.293 | | 0.375 | 0.304 |
| **2012** | 0.337 | 0.486 | 0.339 | 0.948 | 0.948 | 0.948 | 0.516 | 0.559 | 0.409 | 0.516 | | 0.530 | 0.509 | 0.965 | | 0.965 | 0.965 | 0.417 | | 0.483 | 0.324 |
| **2013** | 0.368 | 0.565 | 0.361 | 0.948 | 0.948 | 0.948 | 0.564 | 0.634 | 0.436 | 0.587 | | 0.614 | 0.528 | 0.965 | | 0.965 | 0.965 | 0.435 | | 0.549 | 0.340 |
| **2014** | 0.471 | 0.710 | 0.399 | 0.948 | 0.948 | 0.948 | 0.620 | 0.712 | 0.467 | 0.717 | | 0.726 | 0.554 | 0.965 | | 0.965 | 0.965 | 0.537 | | 0.659 | 0.368 |
| **2015** | 0.506 | 0.811 | 0.438 | 0.948 | 0.948 | 0.948 | 0.672 | 0.790 | 0.502 | 0.742 | | 0.813 | 0.580 | 0.965 | | 0.965 | 0.965 | 0.562 | | 0.739 | 0.395 |
| **2016** | 0.476 | 0.894 | 0.488 | 0.948 | 0.948 | 0.948 | 0.756 | 0.844 | 0.538 | 0.697 | | 0.891 | 0.613 | 0.965 | | 0.965 | 0.965 | 0.628 | | 0.841 | 0.425 |
| **2017** | 0.489 | 0.937 | 0.537 | 0.948 | 0.948 | 0.948 | 0.863 | 0.889 | 0.572 | 0.680 | | 0.949 | 0.648 | 0.965 | | 0.965 | 0.965 | 0.759 | | 0.917 | 0.457 |
| **2018** | 0.589 | 0.925 | 0.637 | 0.948 | 0.948 | 0.948 | 0.944 | 0.925 | 0.651 | 0.775 | | 0.976 | 0.744 | 0.965 | | 0.965 | 0.965 | 0.844 | | 0.959 | 0.562 |
| **2019** | 0.741 | 0.904 | 0.678 |  |  |  | 0.784 | 0.943 | 0.677 | 0.785 | |  | 0.775 |  | |  |  | 0.790 | | 0.976 | 0.596 |
| **2020** | 0.802 | 0.901 | 0.717 |  |  |  | 0.787 | 0.925 | 0.703 | 0.800 | |  | 0.808 |  | |  |  | 0.800 | |  | 0.633 |
| **2021** | 0.802 | 0.909 | 0.749 |  |  |  | 0.836 | 0.913 | 0.727 |  | |  | 0.836 |  | |  |  |  | |  | 0.668 |
| **2022** |  | 0.917 | 0.779 |  |  |  | 0.804 | 0.947 | 0.750 |  | |  | 0.865 |  | |  |  |  | |  | 0.704 |
| **2023** |  | 0.926 | 0.807 |  |  |  | 0.804 |  | 0.773 |  | |  | 0.892 |  | |  |  |  | |  | 0.738 |
| **2024** |  | 0.935 | 0.833 |  |  |  | 0.804 |  | 0.795 |  | |  | 0.918 |  | |  |  |  | |  | 0.772 |
| **2025** |  | 0.944 | 0.858 |  |  |  | 0.804 |  | 0.815 |  | |  | 0.942 |  | |  |  |  | |  | 0.803 |
| **2026** |  | 0.949 | 0.879 |  |  |  | 0.801 |  | 0.832 |  | |  | 0.961 |  | |  |  |  | |  | 0.832 |
| **2027** |  | 0.953 | 0.898 |  |  |  | 0.801 |  | 0.848 |  | |  | 0.976 |  | |  |  |  | |  | 0.976 |
| **2028** |  | 0.955 | 0.915 |  |  |  | 0.801 |  | 0.862 |  | |  |  |  | |  |  |  | |  |  |
| **2029** |  |  | 0.931 |  |  |  | 0.801 |  | 0.874 |  | |  |  |  | |  |  |  | |  |  |
| **2030** |  |  | 0.945 |  |  |  | 0.801 |  | 0.885 |  | |  |  |  | |  |  |  | |  |  |
| **2031** |  |  | 0.955 |  |  |  | 0.802 |  | 0.897 |  | |  |  |  | |  |  |  | |  |  |
| **2032** |  |  |  |  |  |  |  |  | 0.907 |  | |  |  |  | |  |  |  | |  |  |
| **2033** |  |  |  |  |  |  |  |  | 0.917 |  | |  |  |  | |  |  |  | |  |  |
| **2034** |  |  |  |  |  |  |  |  | 0.924 |  | |  |  |  | |  |  |  | |  |  |
| **2035** |  |  |  |  |  |  |  |  | 0.931 |  | |  |  |  | |  |  |  | |  |  |

**^¥^Source:** Circumcision prevalence prior to 2008 is obtained from the Kenya Demographic and Health Survey, 2003. Prevalence of circumcision from 2008 onward combines prevalence of traditional male circumcision and voluntary medical male circumcision estimates obtained from the Decision-Makers' Program Planning Toolkit 2.

###### Table S6b. Number of voluntary medical male circumcisions conducted in South Africa by age group^*^

| **Year** | **Age Group (years)** | | | | | |
| --- | --- | --- | --- | --- | --- | --- |
|  | **10 - 14** | **15 - 19** | **20 - 24** | **25 - 34** | **35 - 49** | **≥50** |
| **2010** | 55,431 | 30,552 | 15,856 | 18,047 | 7,864 | 1,160 |
| **2011** | 137,648 | 75,866 | 39,374 | 44,816 | 19,527 | 2,881 |
| **2012** | 175,060 | 96,487 | 50,075 | 56,996 | 24,834 | 3,664 |
| **2013** | 156,496 | 86,255 | 44,765 | 50,952 | 22,201 | 3,276 |
| **2014** | 199,750 | 110,095 | 57,138 | 65,035 | 28,337 | 4,181 |
| **2015** | 199,535 | 109,976 | 57,076 | 64,965 | 28,306 | 4,176 |
| **2016** | 165,672 | 91,312 | 47,390 | 53,940 | 23,502 | 3,468 |
| **2017** | 203,960 | 112,415 | 58,342 | 66,406 | 28,934 | 4,269 |
| **2018** | 279,500 | 154,050 | 79,950 | 91,000 | 39,650 | 5,850 |
| **2019** | 258,000 | 142,200 | 73,800 | 84,000 | 36,600 | 5,400 |
| **2020** | 236,500 | 130,350 | 67,650 | 77,000 | 33,550 | 4,950 |
| **2021** | 107,500 | 59,250 | 30,750 | 35,000 | 15,250 | 2,250 |
| **2022 onwards** | 43,000 | 23,700 | 12,300 | 14,000 | 6,100 | 900 |

^*^Source: South Africa Department of Health (unpublished data) and South Africa National Strategic Plan for HIV, TB, and STIs 2017-2022(25).

##### Age-specific population fertility rates

###### Table S7a. Age-specific population fertility rates in Kenya 1950-2044^¥^

|  | **Age-specific fertility rates (births per 1,000 women)**  **(age groups in years)** | | | | | | |
| --- | --- | --- | --- | --- | --- | --- | --- |
| **Year** | **15-19** | **20-24** | **25-29** | **30-34** | **35-39** | **40-44** | **45-49** |
| **1950-1955** | 169.1 | 351.6 | 338.1 | 284.3 | 203.5 | 110.7 | 38.9 |
| **1955-1960** | 175.9 | 365.9 | 351.9 | 295.8 | 211.8 | 115.2 | 40.5 |
| **1960-1965** | 182.3 | 379.1 | 364.5 | 306.5 | 219.4 | 119.4 | 41.9 |
| **1965-1970** | 183.3 | 381.2 | 366.6 | 308.2 | 220.6 | 120 | 42.2 |
| **1970-1975** | 180.6 | 375.5 | 361.1 | 303.6 | 217.3 | 118.3 | 41.5 |
| **1975-1980** | 172.7 | 359.1 | 345.3 | 290.3 | 207.8 | 113.1 | 39.7 |
| **1980-1985** | 163.1 | 339.2 | 326.2 | 274.2 | 196.3 | 106.8 | 37.5 |
| **1985-1990** | 147.8 | 307.3 | 295.5 | 248.4 | 177.8 | 96.8 | 34.0 |
| **1990-1995** | 115.3 | 268.9 | 252.0 | 206.8 | 161.6 | 73.4 | 52.0 |
| **1995-2000** | 111.5 | 260.7 | 253.3 | 196.2 | 143.3 | 62.4 | 42.7 |
| **2000-2005** | 104.2 | 243.6 | 236.7 | 183.4 | 133.9 | 58.3 | 39.9 |
| **2005-2010** | 97.1 | 227.1 | 221.4 | 170.6 | 123.7 | 53.6 | 36.5 |
| **2010-2015** | 86.2 | 201.9 | 202.3 | 149.2 | 102.1 | 42.4 | 27.9 |
| **2015-2020** | 75.1 | 176.5 | 179.8 | 129.6 | 85.8 | 34.8 | 22.4 |
| **2020-2024** | 69.9 | 165.1 | 171.8 | 120.2 | 76.2 | 29.9 | 18.6 |
| **2025-2029** | 65.0 | 154.7 | 164.8 | 112.6 | 68.5 | 26.0 | 15.5 |
| **2030-2034** | 60.6 | 145.7 | 159.1 | 106.7 | 62.5 | 22.9 | 13.1 |
| **2035-2039** | 56.2 | 137.2 | 153.6 | 101.8 | 57.6 | 20.4 | 11.0 |
| **2040-2044** | 52.2 | 129.7 | 149.3 | 98.2 | 53.8 | 18.5 | 9.4 |

**^¥^Source**: 2019 World Population Prospects

###### Table S7b. Age-specific population fertility rates in South Africa 1950-2044^¥^

|  | **Age-specific fertility rates (births per 1,000 women)**  **(age groups in years)** | | | | | | |
| --- | --- | --- | --- | --- | --- | --- | --- |
| **Year** | **15-19** | **20-24** | **25-29** | **30-34** | **35-39** | **40-44** | **45-49** |
| **1950-1955** | 66.8 | 265 | 291.9 | 242.2 | 189.8 | 132 | 72.3 |
| **1955-1960** | 65.7 | 260.8 | 287.3 | 238.3 | 186.7 | 130 | 71.2 |
| **1960-1965** | 64.7 | 256.6 | 282.7 | 234.5 | 183.7 | 127.9 | 70 |
| **1965-1970** | 60.4 | 239.7 | 264.1 | 219.1 | 171.7 | 119.5 | 65.4 |
| **1970-1975** | 76.1 | 233.9 | 253.6 | 211 | 160.2 | 105.1 | 54 |
| **1975-1980** | 86.1 | 217.3 | 231.8 | 193.6 | 142.3 | 87.4 | 41.5 |
| **1980-1985** | 93.6 | 201.1 | 211.3 | 177 | 125.9 | 71.7 | 30.5 |
| **1985-1990** | 95.4 | 179.4 | 185.7 | 155.9 | 107.3 | 56 | 20.4 |
| **1990-1995** | 90.8 | 152.2 | 155.2 | 130.8 | 86.9 | 41 | 11.7 |
| **1995-2000** | 80.6 | 140.5 | 142.5 | 111.5 | 74.4 | 31 | 10.3 |
| **2000-2005** | 70.7 | 139 | 141.8 | 105.6 | 67.4 | 27.1 | 8.8 |
| **2005-2010** | 59.2 | 131.7 | 135.1 | 95.9 | 58.4 | 22.6 | 7.1 |
| **2010-2015** | 50.9 | 129 | 133.1 | 90.2 | 52.3 | 19.4 | 5.9 |
| **2015-2020** | 43.6 | 127 | 131.6 | 85.3 | 47 | 16.6 | 4.8 |
| **2020-2024** | 37.2 | 125.7 | 130.8 | 81.3 | 42.4 | 14.1 | 3.8 |
| **2025-2029** | 31.4 | 124.8 | 130.4 | 77.8 | 38.3 | 11.9 | 2.9 |
| **2030-2034** | 26.2 | 124.5 | 130.4 | 74.8 | 34.6 | 9.8 | 2.1 |
| **2035-2039** | 21.5 | 124.8 | 131.3 | 72.5 | 31.4 | 7.9 | 1.4 |
| **2040-2044** | 17 | 125.7 | 132.6 | 70.6 | 28.5 | 6.2 | 0.6 |

**^¥^Source**: 2012 World Population Prospects

###### Table S8a. Age-specific HIV deleted mortality rates in Kenya 1950-2049 by sex ^¥^

|  |  | **Age-specific mortality rates (%)**  **(age groups in years)** | | | | | | |
| --- | --- | --- | --- | --- | --- | --- | --- | --- |
| **Sex** | **Year** | **15-19** | **20-24** | **25-29** | **30-34** | **35-39** | **40-44** | **45-49** |
| **Women** | **1997.5** | 0.136 | 0.191 | 0.244 | 0.294 | 0.361 | 0.453 | 0.542 |
|  | **2002.5** | 0.116 | 0.167 | 0.215 | 0.259 | 0.321 | 0.408 | 0.493 |
|  | **2007.5** | 0.100 | 0.145 | 0.188 | 0.229 | 0.286 | 0.367 | 0.449 |
|  | **2012.5** | 0.085 | 0.127 | 0.166 | 0.202 | 0.254 | 0.331 | 0.408 |
|  | **2017.5** | 0.073 | 0.110 | 0.145 | 0.178 | 0.226 | 0.298 | 0.371 |
|  | **2022.5** | 0.063 | 0.096 | 0.128 | 0.158 | 0.201 | 0.268 | 0.338 |
|  | **2027.5** | 0.054 | 0.084 | 0.112 | 0.139 | 0.179 | 0.241 | 0.307 |
|  | **2032.5** | 0.046 | 0.073 | 0.099 | 0.123 | 0.160 | 0.217 | 0.280 |
|  | **2037.5** | 0.039 | 0.064 | 0.087 | 0.108 | 0.142 | 0.196 | 0.254 |
|  | **2042.5** | 0.034 | 0.055 | 0.076 | 0.096 | 0.126 | 0.176 | 0.231 |
| **Men** | **1997.5** | 0.165 | 0.253 | 0.288 | 0.345 | 0.433 | 0.551 | 0.713 |
|  | **2002.5** | 0.142 | 0.219 | 0.251 | 0.304 | 0.385 | 0.494 | 0.648 |
|  | **2007.5** | 0.121 | 0.189 | 0.219 | 0.267 | 0.343 | 0.443 | 0.589 |
|  | **2012.5** | 0.104 | 0.163 | 0.191 | 0.235 | 0.305 | 0.398 | 0.535 |
|  | **2017.5** | 0.089 | 0.141 | 0.166 | 0.207 | 0.271 | 0.357 | 0.486 |
|  | **2022.5** | 0.076 | 0.122 | 0.145 | 0.182 | 0.241 | 0.320 | 0.441 |
|  | **2027.5** | 0.065 | 0.106 | 0.126 | 0.160 | 0.215 | 0.287 | 0.401 |
|  | **2032.5** | 0.056 | 0.091 | 0.110 | 0.141 | 0.191 | 0.258 | 0.364 |
|  | **2037.5** | 0.048 | 0.079 | 0.096 | 0.124 | 0.170 | 0.231 | 0.331 |
|  | **2042.5** | 0.041 | 0.068 | 0.083 | 0.109 | 0.151 | 0.208 | 0.300 |

**^¥^Source**: 2019 World Population Prospects

###### Table S8b. Age-specific mortality rates in South Africa 1950-2049 by sex^¥^

|  |  | **Age-specific mortality rates (%)**  **(age groups in years)** | | | | | | |
| --- | --- | --- | --- | --- | --- | --- | --- | --- |
| **Sex** | **Year** | **15-19** | **20-24** | **25-29** | **30-34** | **35-39** | **40-44** | **45-49** |
| **Women** | **1997.5** | 0.116 | 0.163 | 0.215 | 0.263 | 0.351 | 0.482 | 0.702 |
|  | **2002.5** | 0.096 | 0.134 | 0.179 | 0.222 | 0.302 | 0.424 | 0.629 |
|  | **2007.5** | 0.079 | 0.111 | 0.150 | 0.187 | 0.260 | 0.373 | 0.563 |
|  | **2012.5** | 0.066 | 0.092 | 0.125 | 0.158 | 0.223 | 0.328 | 0.504 |
|  | **2017.5** | 0.054 | 0.076 | 0.105 | 0.133 | 0.192 | 0.288 | 0.451 |
|  | **2022.5** | 0.045 | 0.063 | 0.087 | 0.113 | 0.165 | 0.254 | 0.404 |
|  | **2027.5** | 0.037 | 0.052 | 0.073 | 0.095 | 0.142 | 0.223 | 0.362 |
|  | **2032.5** | 0.031 | 0.043 | 0.061 | 0.080 | 0.122 | 0.196 | 0.324 |
|  | **2037.5** | 0.025 | 0.035 | 0.051 | 0.068 | 0.105 | 0.172 | 0.290 |
|  | **2042.5** | 0.021 | 0.029 | 0.042 | 0.057 | 0.090 | 0.152 | 0.260 |
| **Men** | **1997.5** | 0.177 | 0.312 | 0.361 | 0.437 | 0.594 | 0.872 | 1.264 |
|  | **2002.5** | 0.169 | 0.281 | 0.327 | 0.395 | 0.541 | 0.803 | 1.177 |
|  | **2007.5** | 0.173 | 0.254 | 0.296 | 0.358 | 0.494 | 0.739 | 1.095 |
|  | **2012.5** | 0.156 | 0.230 | 0.268 | 0.324 | 0.451 | 0.681 | 1.019 |
|  | **2017.5** | 0.141 | 0.207 | 0.243 | 0.294 | 0.411 | 0.627 | 0.949 |
|  | **2022.5** | 0.127 | 0.187 | 0.220 | 0.266 | 0.375 | 0.578 | 0.883 |
|  | **2027.5** | 0.115 | 0.169 | 0.199 | 0.241 | 0.342 | 0.532 | 0.822 |
|  | **2032.5** | 0.103 | 0.153 | 0.180 | 0.218 | 0.312 | 0.490 | 0.765 |
|  | **2037.5** | 0.093 | 0.138 | 0.163 | 0.197 | 0.285 | 0.451 | 0.713 |
|  | **2042.5** | 0.082 | 0.124 | 0.148 | 0.179 | 0.260 | 0.415 | 0.663 |

**^¥^Source**: 2012 World Population Prospects

##### Model fit to age-specific and overall prevalence from population-based surveys by sex

###### Figure S1a Model fit to age-specific and overall prevalence from population-based surveys by sex in Kenya

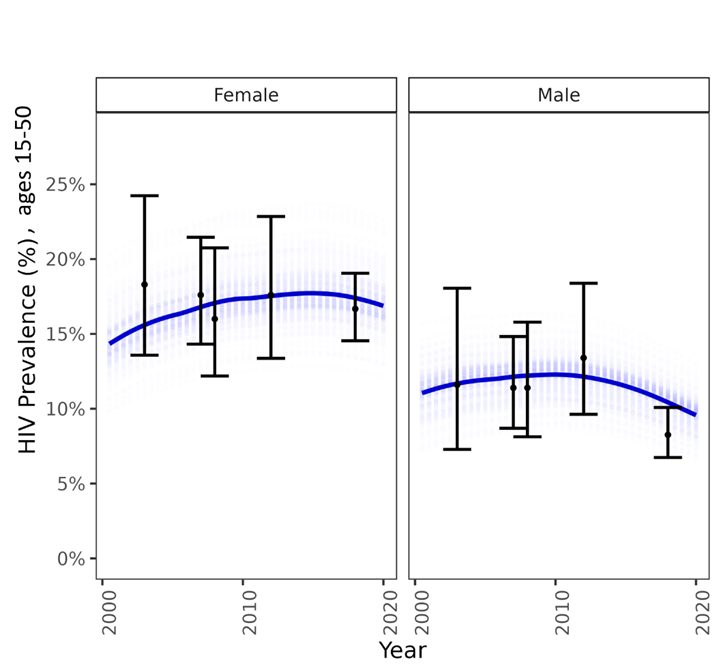

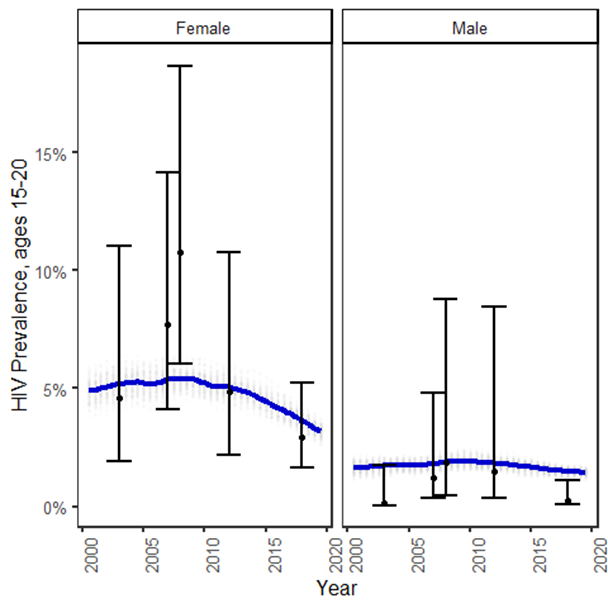

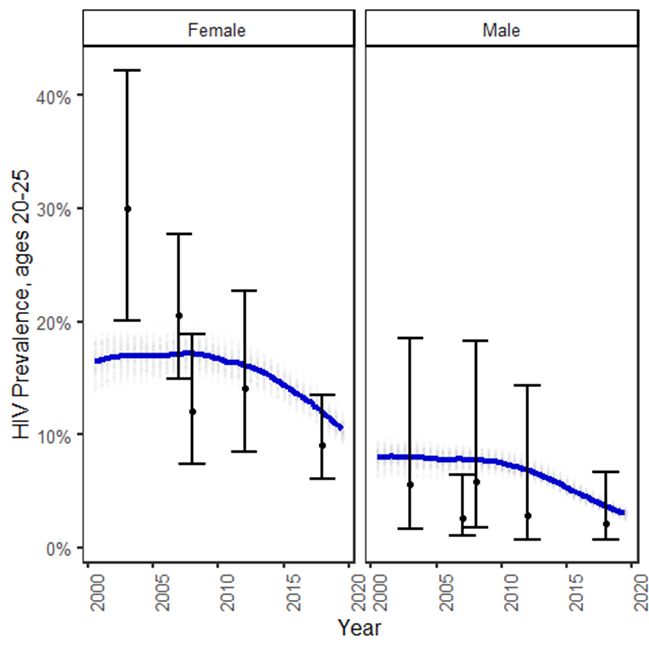

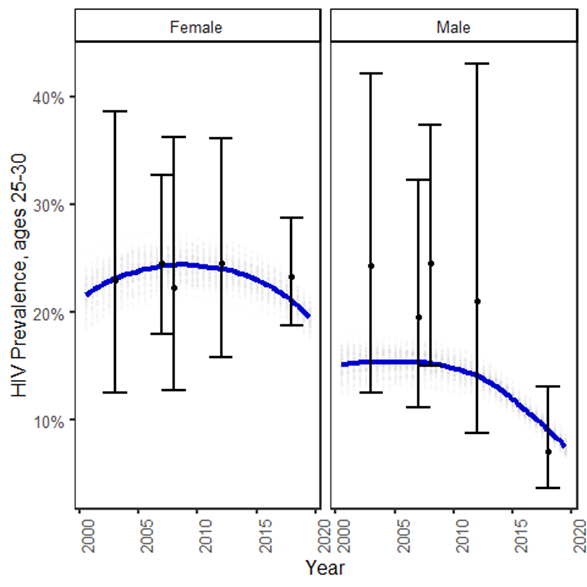

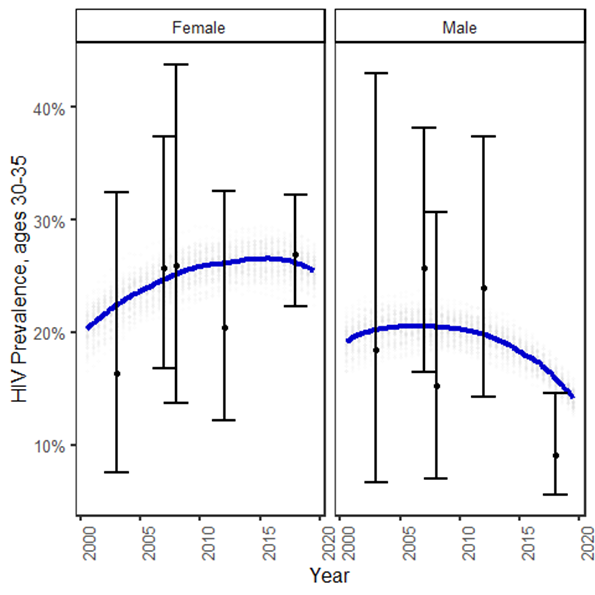

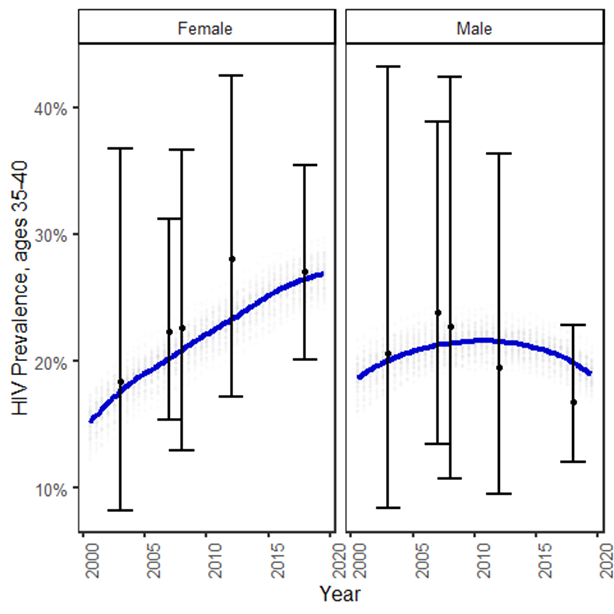

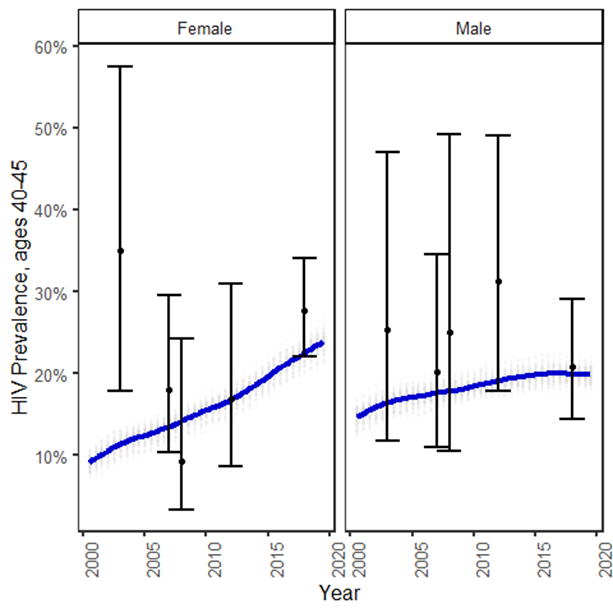

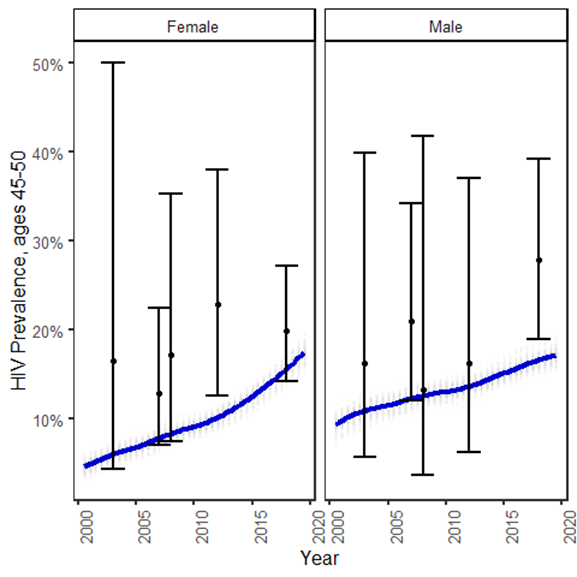

* Blue curves refer to the average over the 100 model simulations; the error bars refer to the empirical estimates and 95% confidence intervals for HIV prevalence obtained from Kenya Demographic and Health Surveys and Kenya AIDS Indicator Surveys; 95% confidence intervals were calculated using DHS survey weights and accounting for strata.

###### Figure S1b Model fit to age-specific and overall prevalence from population-based surveys by sex in South Africa

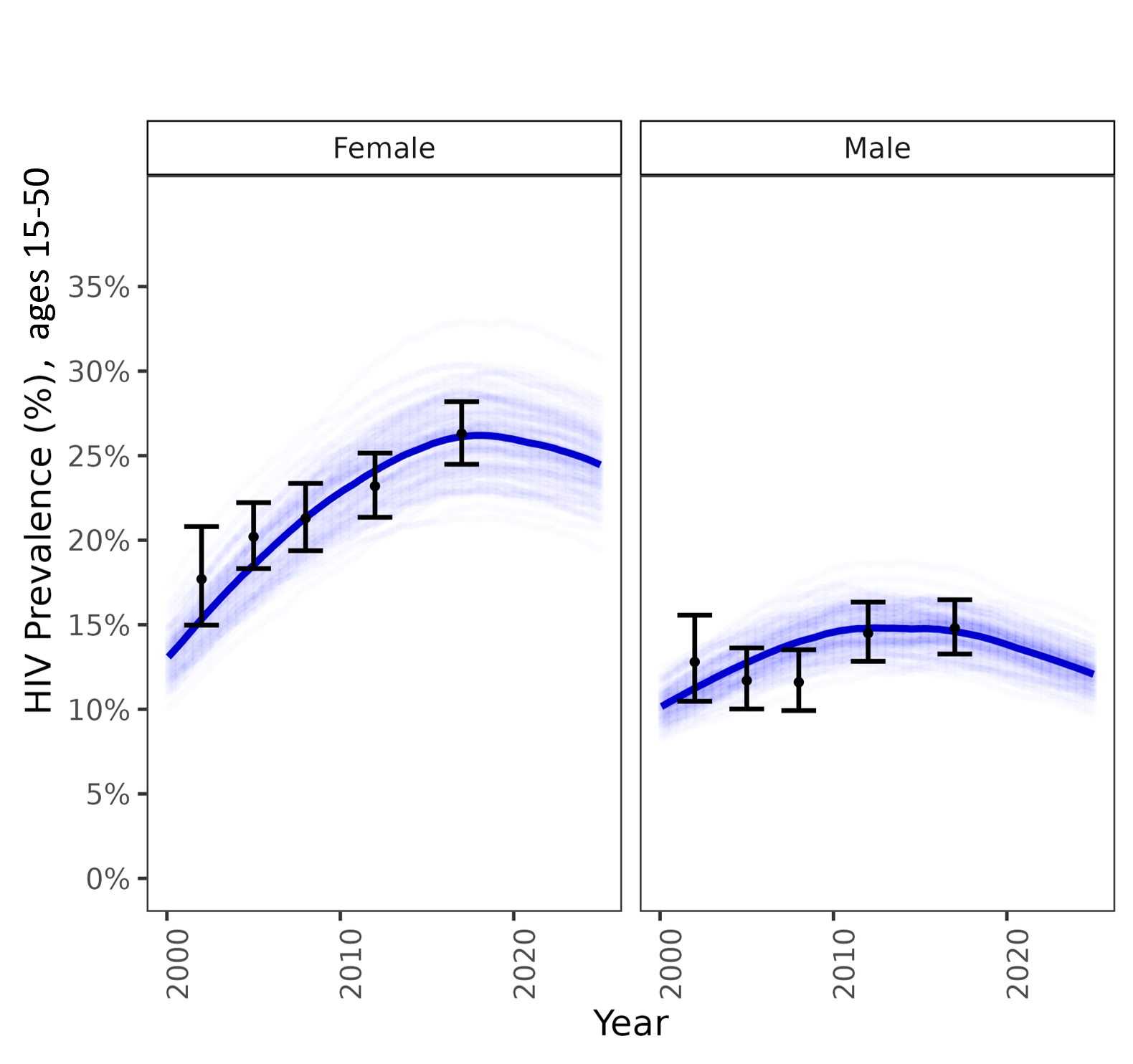

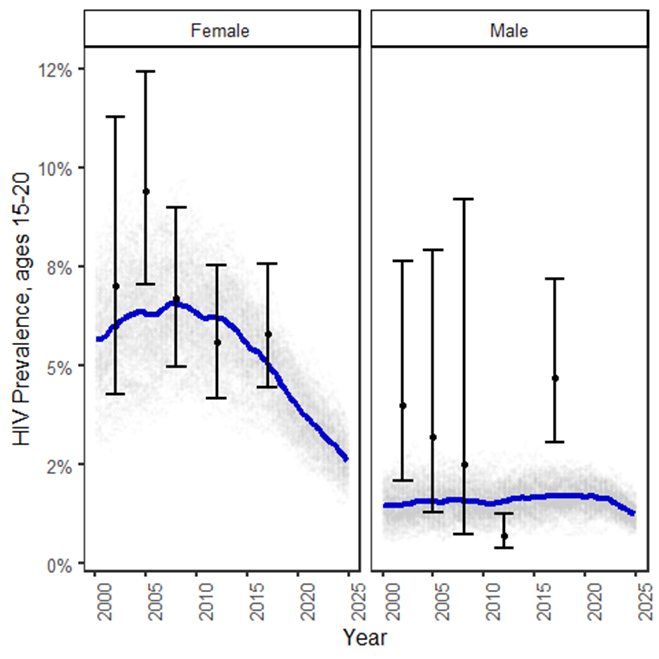

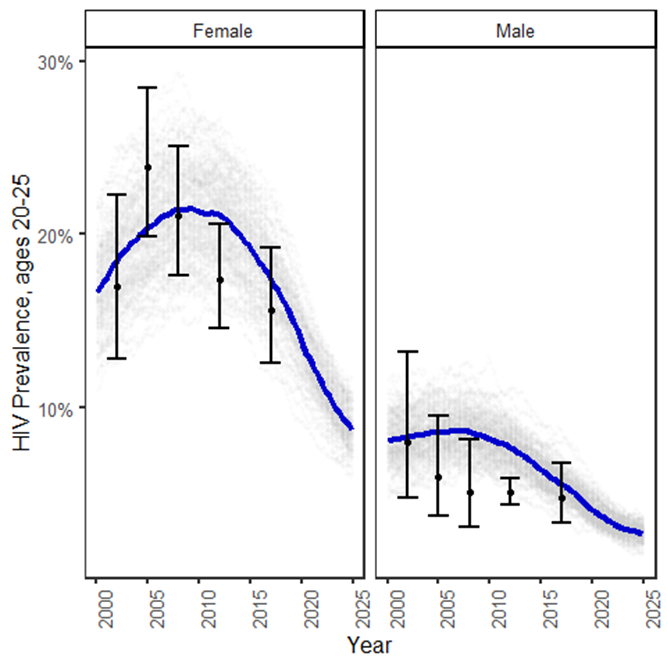

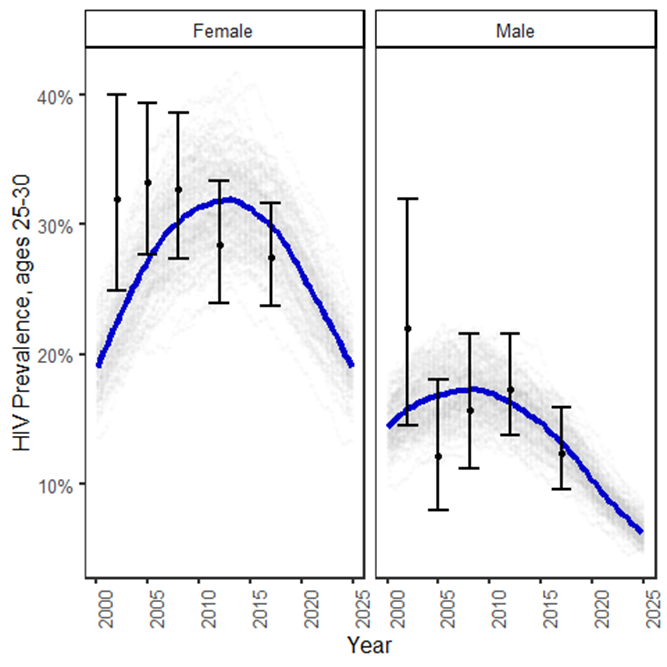

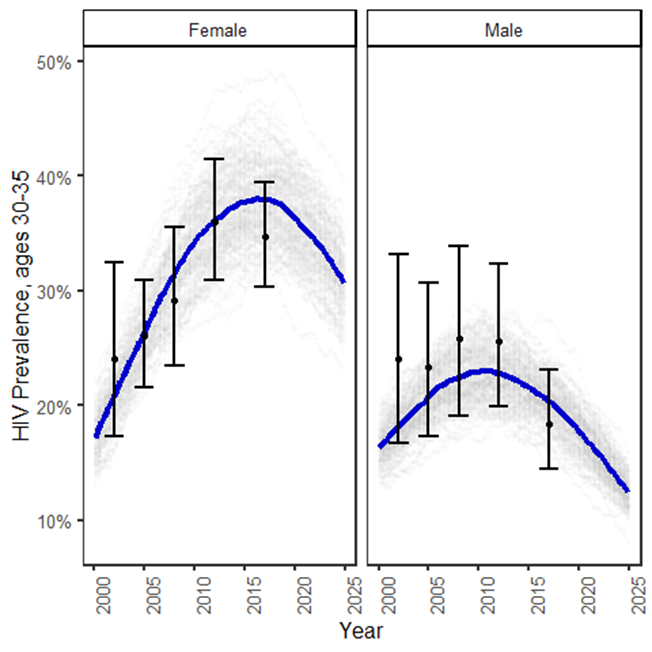

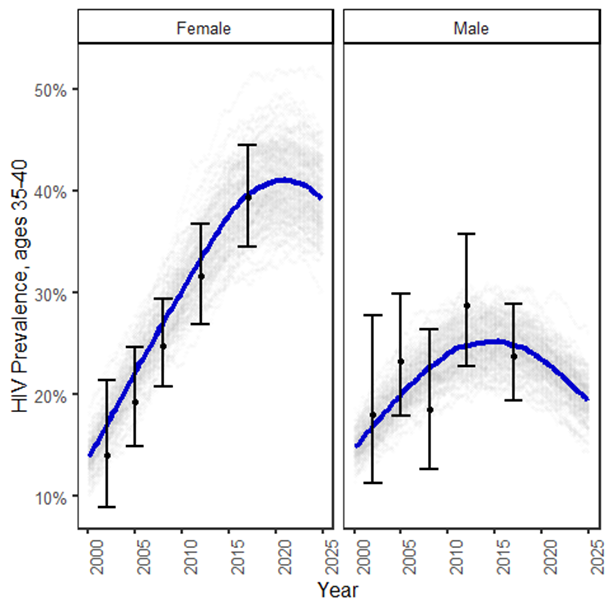

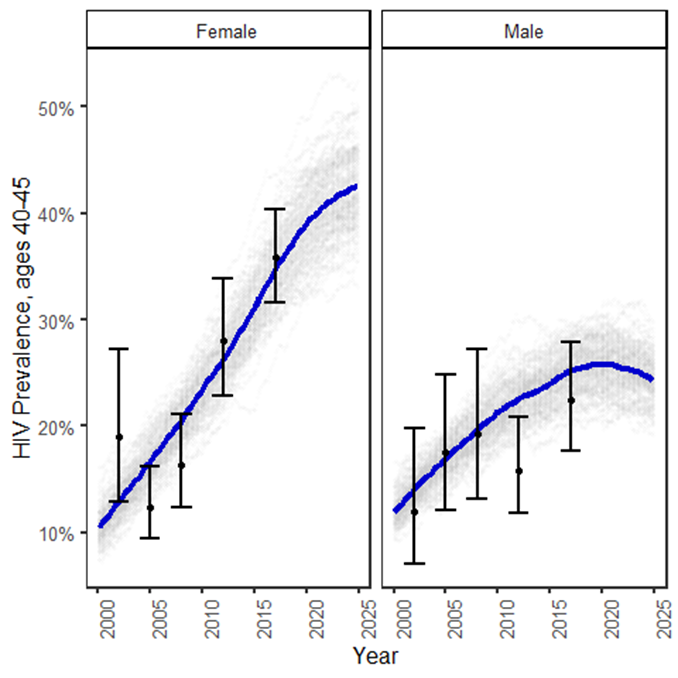

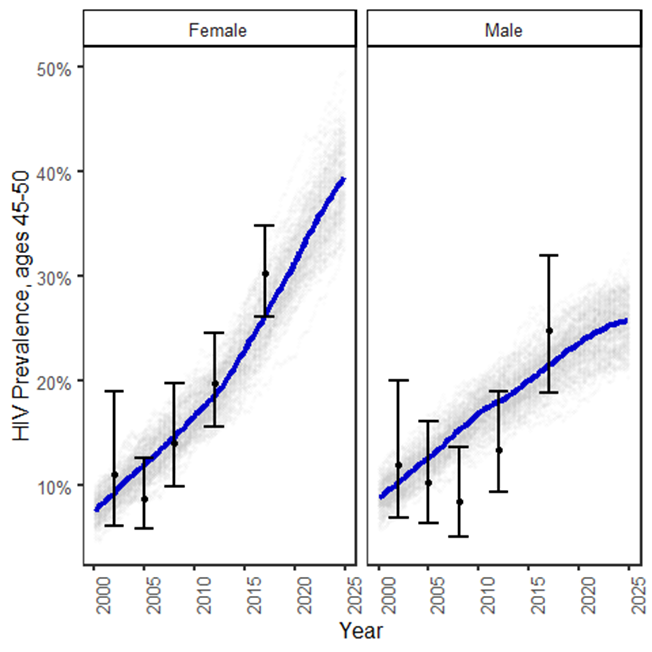

* Blue curves refer to the average over the 100 model simulations;; the error bars refer to the empirical estimates and 95% confidence intervals for HIV prevalence obtained from South African National HIV Prevalence, Incidence and Behaviour Surveys (2002, 2005, 2008, 2012 and 2017) from the Human Sciences Research Council (HSRC)

##### Model fit to age-specific and overall prevalence from population-based surveys by sex

###### Figure S2a Model fit to age-specific and overall ART coverage from population-based surveys by sex in Kenya^*^

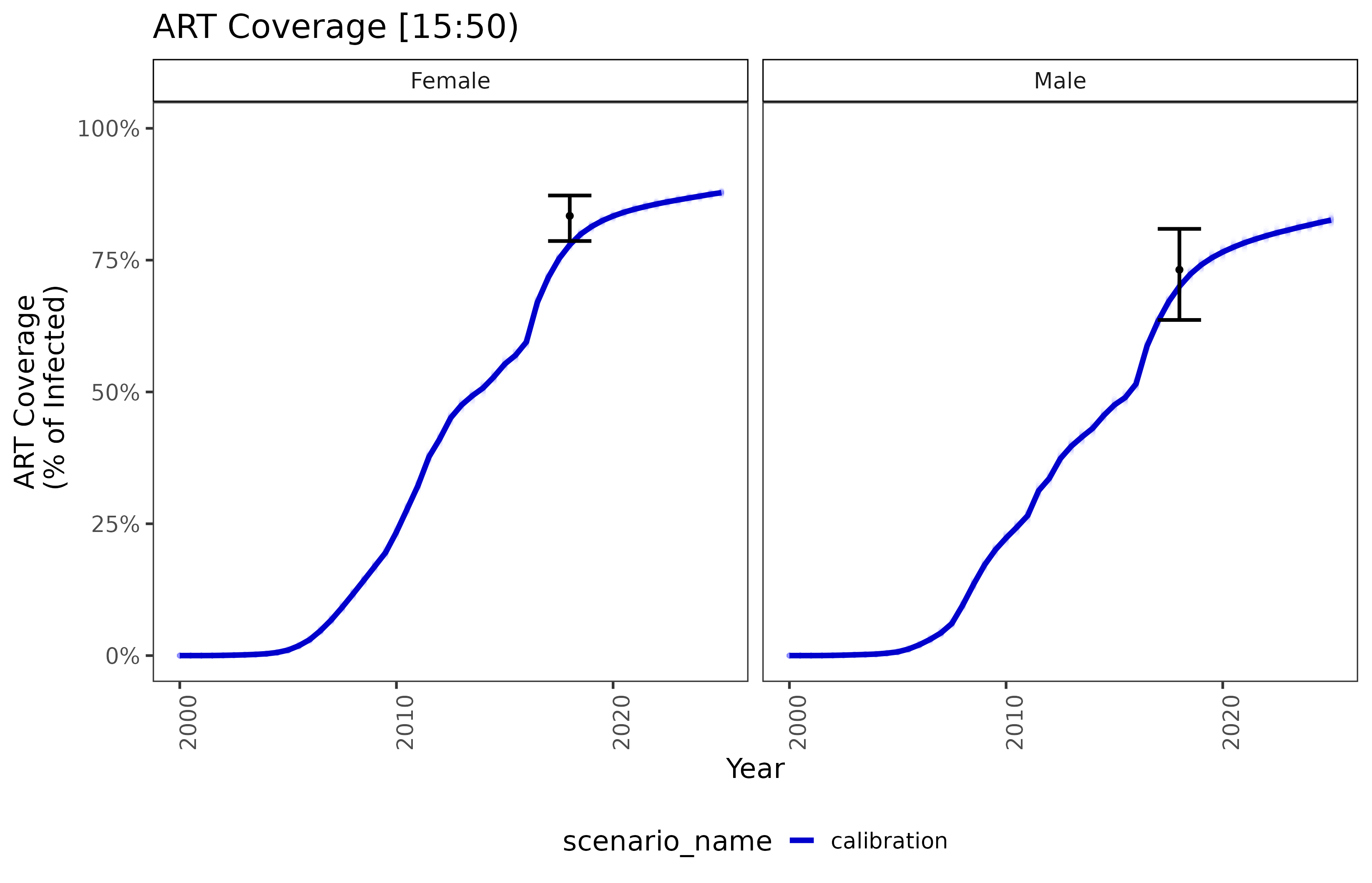

^*^ There is a lack of data on historic ART coverage but the model fits well to the empiric data point and is calibrated to number of people on ART over time by sex, which provides confidence about historical fit.

###### Figure S2b Model fit to age-specific and overall ART coverage from population-based surveys by sex in South Africa

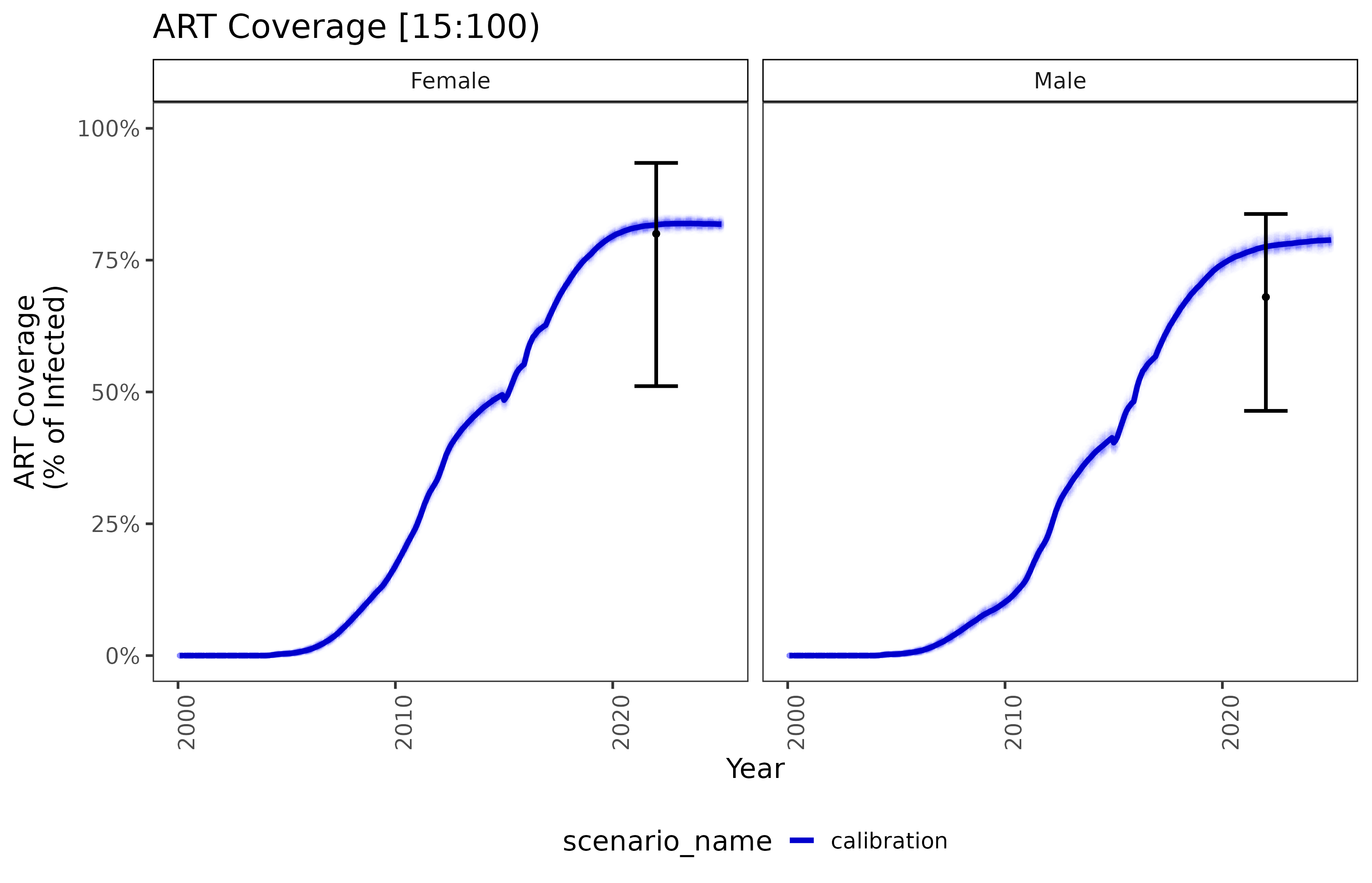

^*^ The model fits well to the empiric data point for current coverage and is calibrated to number of people on ART over time by sex, which provides confidence about historical fit.

#### Model Overview and Parameters

*Model initialization*

The simulations begin prior to the start of infection in the year 1960 to allow sufficient time for the epidemic to burn-in. During this time, individuals with demographic properties specified in the demographics file begin forming relationships; relationship formation rates for each sex and relationship type are updated daily using a relationship flow algorithm. Adjustment of pair formation entry rates is terminated at a specific timepoint (e.g., 1975) and the rates are fixed at that value for the remainder of the simulation. The age profile of the population is initialized using demographic data including population age distribution or age-specific fertility rates. A prior analysis of our model evaluated the age/sex pairings, partnership length and other sexual network characteristics and confirmed that these outputs reached equilibrium within 20 years, prior to the introduction of HIV into the model: <https://ieeexplore.ieee.org/abstract/document/6426573>. ^1^ HIV infections are seeded in 1980 and affect a certain proportion of the population based on the age and sex distribution reported from historical data. ART intervention is introduced in the year 2012. Eligible individuals enroll in ART based on historical eligibility criteria based on CD4 count, which changes over time based on WHO guidelines for ART initiation until the implementation of universal ART, which is assumed to remain the same until the end of the simulations (year 2050). Calibration and validation processes are performed to refine the initialization and ensure that the model aligns with observed HIV dynamics in the target population ^2,3^.

The following provides a more detailed description of the model initialization process: <https://docs.idmod.org/projects/emod-hiv/en/latest/sti-model-relationships.html>. ^2^

EMOD-HIV is open-source and publicly available online: <https://docs.idmod.org/projects/emod-hiv/en/2.20_a/>

*Modelled time step*

The model is implemented using a monthly time step, aggregating events and changes over the course of each month. Monthly updated information can then be ascertained regarding a range of activities and occurrences, including sexual mixing, relationship formation, stages of HIV infection, HIV testing and its results, and PrEP initiation and discontinuation.

*PrEP cascade*

HIV-negative individuals aged 18 to 49 years were eligible for PrEP if testing negative at the time of initiation or continuation, and if sexually active with at least one partner. PrEP discontinuation occurred if individuals are lost to follow-up or no longer met the eligibility criteria (i.e., an individual turned 50 years old, all partnerships end, or tested HIV-positive). Individuals who stopped PrEP can re-start at any time if they meet the eligibility criteria (i.e. start a new partnership).

##### Model Parameters

###### Table S9a. Select model parameters used to fit the EMOD-HIV transmission model to survey data on prevalence and ART coverage from Kenya.

| **Parameter** | **Parameter Description** | **Fitted median** | **(IQR)** |
| --- | --- | --- | --- |
| ARTLinkMax | Maximum probability of linkage to ART | 0.999 | (0.978, 1.000) |
| ARTLinkMid | Year of ART linkage (given eligibility), that is, time of the inflection point in the sigmoid trend. | 2003.008 | (2002.920, 2003.837) |
| AcuteDurationMonths | The time since infection, in months, over which the Acute_Stage_Infectivity_Multiplier is applied to coital acts occurring in that time period. | 1.000 | (1.000, 1.331) |
| CircumcisionReducedAcquire | The reduction of susceptibility to STI by voluntary male medical circumcision (VMMC). | 0.600 | (0.598, 0.600) |
| Homa_BayInfrmlCondomsMax | Maximum rate of condom use in informal relationships in Homa Bay | 0.231 | (0.230, 0.233) |
| Homa_BayLOWRisk | Proportion of the population that is low-risk in Homa Bay | 0.557 | (0.549, 0.559) |
| Homa_BayTrnsCondomsMax | Maximum rate of condom use in transitory relationships in Homa Bay | 0.244 | (0.232, 0.245) |
| InfmrlFormRate | Informal relationship formation rate | 0.000 | (0.000, 0.000) |
| InfrmlCondomMid | Year midpoint of logistic scale-up of condom use in informal relationships | 1993.007 | (1991.629, 1998.680) |
| InfrmlCondomRate | Rate of logistic scale-up of condom use in informal relationships | 2.661 | (1.953, 2.909) |
| InfrmlCondomsMax | Maximum rate of condom use in informal relationships | 0.216 | (0.215, 0.217) |
| InfrmlDurHet | Heterogeneity in duration of informal relationships | 0.750 | (0.750, 0.750) |
| KisiiInfrmlCondomsMax | Maximum rate of condom use in informal relationships in Kisii | 0.231 | (0.226, 0.232) |
| KisiiLOWRisk | Proportion of the population that is low-risk in Kisii | 0.938 | (0.935, 0.939) |
| KisiiTrnsCondomsMax | Maximum rate of condom use in transitory relationships in Kisii | 0.375 | (0.369, 0.376) |
| KisumuInfrmlCondomsMax | Maximum rate of condom use in informal relationships in Kisumu | 0.182 | (0.181, 0.183) |
| KisumuLOWRisk | Proportion of the population that is low-risk in Kisumu | 0.764 | (0.762, 0.765) |
| KisumuTrnsCondomsMax | Maximum rate of condom use in transitory relationships in Kisumu | 0.338 | (0.337, 0.347) |
| LogBaseInfectivity | The probability of transmission when none of the transmission multipliers apply to a particular coital act. | 0.002 | (0.002, 0.002) |
| MaleToFemaleOld | Male-to-female relative risk of infection among older individuals | 2.012 | (1.506, 2.132) |
| MaleToFemaleYoung | Male-to-female relative risk of infection among young individuals | 1.116 | (1.000, 1.242) |
| MaxInfrmlFLOW | Maximum number of informal relationships among low-risk females | 1.311 | (1.255, 1.315) |
| MaxInfrmlFMED | Maximum number of informal relationships among medium-risk females | 2.050 | (2.002, 2.351) |
| MaxInfrmlMLOW | Maximum number of informal relationships among low-risk males | 1.165 | (1.159, 1.203) |
| MaxInfrmlMMED | Maximum number of informal relationships among medium-risk males | 2.530 | (2.442, 2.735) |
| MaxMrtlFMED | Maximum number of marital relationship among medium-risk females | 1.139 | (1.136, 1.159) |
| MaxMrtlMMED | Maximum number of marital relationship among medium-risk males | 1.302 | (1.268, 1.316) |
| MaxTrnsFLOW | Maximum number of transitory relationships among low-risk females | 1.599 | (1.580, 1.625) |
| MaxTrnsFMED | Maximum number of transitory relationships among medium-risk females | 2.943 | (2.891, 3.000) |
| MaxTrnsMLOW | Maximum number of transitory relationships among low-risk males | 1.599 | (1.588, 1.677) |
| MaxTrnsMMED | Maximum number of transitory relationships among medium-risk males | 2.557 | (2.475, 2.879) |
| MigoriInfrmlCondomsMax | Maximum rate of condom use in informal relationships in Migori | 0.186 | (0.185, 0.189) |
| MigoriLOWRisk | Proportion of the population that is low-risk in Migori | 0.795 | (0.793, 0.797) |
| MigoriTrnsCondomsMax | Maximum rate of condom use in transitory relationships in Migori | 0.249 | (0.248, 0.257) |
| MrtlCondomMax | Maximum rate of condom use in marital relationships | 0.192 | (0.191, 0.193) |
| MrtlCondomMid | Year midpoint of logistic scale-up of condom use in marital relationships | 2003.131 | (1999.017, 2005.000) |
| MrtlCondomRate | Rate of logistic scale-up of condom use in marital relationships | 2.971 | (2.595, 3.000) |
| MrtlFormRate | Marital relationship formation rate | 0.000 | (0.000, 0.000) |
| NyamiraInfrmlCondomsMax | Maximum rate of condom use in informal relationships in Nyamira | 0.096 | (0.093, 0.097) |
| NyamiraLOWRisk | Proportion of the population that is low-risk in Nyamira | 0.902 | (0.900, 0.910) |
| NyamiraTrnsCondomsMax | Maximum rate of condom use in transitory relationships in Nyamira | 0.309 | (0.304, 0.313) |
| PrExInfrmlFemLOW | Probability of potential for extra-relational informal relationship among low-risk females | 0.366 | (0.364, 0.371) |
| PrExInfrmlFemMED | Probability of potential for extra-relational informal relationship among medium-risk females | 0.401 | (0.401, 0.407) |
| PrExInfrmlMaleLOW | Probability of potential for extra-relational informal relationship among low-risk males | 0.244 | (0.222, 0.249) |
| PrExInfrmlMaleMED | Probability of potential for extra-relational informal relationship among medium-risk males | 0.382 | (0.379, 0.389) |
| PrExTrnsFemLOW | Probability of potential for extra-relational transitory relationship among low-risk females | 0.033 | (0.033, 0.033) |
| PrExTrnsFemMED | Probability of potential for extra-relational transitory relationship among medium-risk females | 0.460 | (0.450, 0.468) |
| PrExTrnsMaleLOW | Probability of potential for extra-relational transitory relationship among low-risk males | 0.330 | (0.328, 0.332) |
| PrExTrnsMaleMED | Probability of potential for extra-relational transitory relationship among medium-risk males | 0.593 | (0.589, 0.611) |
| PreARTLinkMax | Maximum probability of linkage to pre-ART care | 0.715 | (0.708, 0.737) |
| PreARTLinkMid | Year midpoint of logistic scale-up of pre-ART linkage | 1996.709 | (1995.685, 1998.896) |
| PreARTLinkMin | Minimum probability of linkage to pre-ART care | 0.435 | (0.416, 0.442) |
| RiskAssortivity | Risk assortivity | 0.663 | (0.648, 0.673) |
| SeedYrHigh | Seed year | 1986.562 | (1982.000, 1988.000) |
| SexualDebutAgeFemaleWeibullHeterogeneity | Heterogeneity parameter of Weibull distribution of female age of sexual debut | 0.086 | (0.083, 0.086) |
| SexualDebutAgeFemaleWeibullScale | Scale parameter of Weibull distribution of female age of sexual debut | 16.013 | (15.540, 16.038) |
| SexualDebutAgeMaleWeibullHeterogeneity | Heterogeneity parameter of Weibull distribution of male age of sexual debut | 0.040 | (0.040, 0.040) |
| SexualDebutAgeMaleWeibullScale | Scale parameter of Weibull distribution of male age of sexual debut | 15.708 | (15.155, 16.090) |
| SiayaInfrmlCondomsMax | Maximum rate of condom use in informal relationships in Siaya | 0.160 | (0.148, 0.161) |
| SiayaLOWRisk | Proportion of the population that is low-risk in Siaya | 0.729 | (0.722, 0.731) |
| SiayaTrnsCondomsMax | Maximum rate of condom use in transitory relationships in Siaya | 0.300 | (0.293, 0.304) |
| TrnsCondomMax | Maximum rate of condom use in transitory relationships | 0.243 | (0.242, 0.253) |
| TrnsCondomMid | Year midpoint of logistic scale-up of condom use in transitory relationships | 1997.960 | (1996.962, 1999.273) |
| TrnsCondomRate | Rate of logistic scale-up of condom use in transitory relationships | 0.999 | (0.978, 1.000) |
| TrnsFormRate | Transitory relationship formation rate | 2003.008 | (2002.920, 2003.837) |

* Median and interquartile ranges (IQRs) reported for all dynamic parameters used in the calibration process from 100 best-fitting parameter sets. †

###### Table S9b. Select model parameters used to fit the EMOD-HIV transmission model to survey data on prevalence and ART coverage from South Africa.

| **Parameter** | **Description** | **Fitted Median** | **(IQR)** |
| --- | --- | --- | --- |
| ART Link Max | Maximum probability of linkage to ART | 1.000 | (0.997, 1.000) |
| ART Link Mid | Year of ART linkage (given eligibility), that is, time of the inflection point in the sigmoid trend. | 2,005.96 | (2,005.86, 2,006.08) |
| All: Infmrl Condom | Modern condom usage rate in informal relationships across all locations | 0.61 | (0.58, 0.64) |
| All: LOW Risk | Proportion of the population that is low-risk | 0.936 | (0.930, 0.942) |
| All: Trns Condom | Modern condom usage rate in transitory relationships across all locations | 0.2 | (0.16, 0.23) |
| Base Infectivity | The probability of transmission when none of the transmission multipliers apply to a coital act (or when all multipliers are set to 1). | 0.0015 | (0.0015, 0.0016) |
| Circumcision Reduced Acquire | The reduction of susceptibility to STI by voluntary male medical circumcision (VMMC). | 0.6 |  |
| Infmrl Condoms Late | Modern condom usage rate in informal relationships, by county | 0.37 | (0.35, 0.39) |
| Infmrl Form Rate | Informal relationship formation rate | 0.0009 | (0.0008, 0.0009) |
| Infrml Condom Mid | Year midpoint of logistic scale-up of condom use in informal relationships | 1,998.27 | (1,997.81, 1,999.03) |
| Infrml Condom Rate | Rate of logistic scale-up of condom use in informal relationships | 2.03 | (1.89, 2.18) |
| Infrml Dur Het | Heterogeneity in duration of informal relationships | 0.693 | (0.675, 0.711) |
| Male To Female Old | Male-to-female relative risk of infection among older individuals | 2.33 | (2.22, 2.49) |
| Male To Female Young | Male-to-female relative risk of infection among young individuals | 2.97 | (2.74, 3.37) |
| Max Infmrl F LOW | Maximum number of informal relationships among low-risk females | 1.67 | (1.61, 1.72) |
| Max Infmrl F MED | Maximum number of informal relationships among medium-risk females | 0.92 | (0.89, 0.94) |
| Max Infmrl M LOW | Maximum number of informal relationships among low-risk males | 1.73 | (1.69, 1.76) |
| Max Infmrl M MED | Maximum number of informal relationships among medium-risk males | 0.75 | (0.70, 0.79) |
| Max Mrtl F ME | Maximum number of marital relationships among medium-risk females | 1.23 | (1.16, 1.27) |
| Max Mrtl M ME | Maximum number of marital relationships among medium-risk males | 0.93 | (0.91, 0.96) |
| Max Trns F LOW | Maximum number of transitory relationships among low-risk females | 1.5 | (1.48, 1.52) |
| Max Trns F MED | Maximum number of transitory relationships among medium-risk females | 3.1 | (3.06, 3.14) |
| Max Trns M LOW | Maximum number of transitory relationships among low-risk males | 1.49 | (1.46, 1.53) |
| Max Trns M MED | Maximum number of transitory relationships among medium-risk males | 3.02 | (2.88, 3.10) |
| Mrtl Condom Max | Maximum rate of condom use in marital relationships | 0.191 | (0.184, 0.211) |
| Mrtl Condom Mid | Year midpoint of logistic scale-up of condom use in marital relationships | 1,994.92 | (1,994.19, 1,995.27) |
| Mrtl Condom Rate | Rate of logistic scale-up of condom use in marital relationships | 3.6 | (3.49, 3.69) |
| Mrtl Form Rate | Marital relationship formation rate | 0.0001 | (0.0001, 0.0001) |
| Pr Ex Infmrl Fem LOW | Probability of potential for extra-relational informal relationship among low-risk females | 0.08 | (0.06, 0.10) |
| Pr Ex Infmrl Fem MED | Probability of potential for extra-relational informal relationship among medium-risk females | 0.391 | (0.381, 0.404) |
| Pr Ex Infmrl Male LOW | Probability of potential for extra-relational informal relationship among low-risk males | 0.46 | (0.42, 0.50) |
| Pr Ex Infmrl Male MED | Probability of potential for extra-relational informal relationship among medium-risk males | 0.379 | (0.370, 0.390) |
| Pr Ex Trns Fem LOW | Probability of potential for extra-relational transitory relationship among low-risk females | 0.066 | (0.058, 0.081) |
| Pr Ex Trns Fem MED | Probability of potential for extra-relational transitory relationship among medium-risk females | 0.58 | (0.56, 0.62) |
| Pr Ex Trns Male LOW | Probability of potential for extra-relational transitory relationship among low-risk males | 0.17 | (0.12, 0.19) |
| Pr Ex Trns Male MED | Probability of potential for extra-relational transitory relationship among medium-risk males | 0.59 | (0.58, 0.62) |
| PreART Link Max | Maximum probability of linkage to pre-ART care | 0.89 | (0.85, 0.96) |
| PreART Link Mid | Year midpoint of logistic scale-up of pre-ART linkage | 1,998.28 | (1,997.76, 1,998.72) |
| PreART Link Min | Minimum probability of linkage to pre-ART care | 0.63 | (0.62, 0.65) |
| Risk Assortivity | Risk assortivity | 0.47 | (0.44, 0.49) |
| SeedYr HIGH | Seed year | 1991 |  |
| Sexual Debut Age Female Weibull Heterogeneity | Heterogeneity parameter of Weibull distribution of female age of sexual debut | 0.062 | (0.051, 0.067) |
| Sexual Debut Age Female Weibull Scale | Scale parameter of Weibull distribution of female age of sexual debut | 16.49 | (16.34, 16.62) |
| Sexual Debut Age Male Weibull Heterogeneity | Heterogeneity parameter of Weibull distribution of male age of sexual debut | 0.043 | (0.038, 0.050) |
| Sexual Debut Age Male Weibull Scale | Scale parameter of Weibull distribution of male age of sexual debut | 16.47 | (16.24, 16.62) |
| Trns Condom Late | Modern condom usage rate in transitory relationships | 0.61 | (0.59, 0.64) |
| Trns Condom Mid | Year midpoint of logistic scale-up of condom use in transitory relationships | 2,006.77 | (2,006.08, 2,007.21) |
| Trns Condom Rate | Rate of logistic scale-up of condom use in transitory relationships | 1.94 | (1.84, 2.01) |
| Trns Form Rate | Transitory relationship formation rate | 0.0013 | (0.0013, 0.0014) |

* Median and interquartile ranges (IQRs) reported for all dynamic parameters used in the calibration process from 100 best-fitting parameter sets. †

###### Table S11. Utility weights for estimating disability-adjusted life-years averted

| Health State | DALY Weight | Reference |
| --- | --- | --- |
| HIV-negative | 0 | Vos *et al* ^4^ |
| HIV and not on ART | 0.274 |  |
| HIV and on ART | 0.078 |  |

##### Costing parameters

###### Table S11 Western Kenya costing parameter calculations

| **Cost parameter** | **Estimate (USD)** | **Year** | **Data source** | **Calculation details, notes** |
| --- | --- | --- | --- | --- |
| **HIV costs** |  |  |  |  |
| Annual health care costs (among those not on ART) |  |  |  |  |
| HIV-positive CD4 < 200 | 110.3 | 2021 | Eaton 2014 ^5^ | Adjusted for inflation and GDP/capita ratio using this approach: **Step 1:** Adjust South Africa value in 2012 USD for inflation to be in 2021 USD ($374.08 = $167*2.24)  Cost of health care use, CD4 count <200 cells per μL, not in HIV care (per person-year) in South Africa= $167 USD Inflation Rate between time of costing (2012) and 2021: 2.24 =4.7/2.1 **Step 2:** Adjust South Africa 2021 USD value by multiplying by the Kenya GDP/cap ratio ($374.08*0.295) South Africa 2021 GDP per capita in $USD = 7,055; Kenya $USD = 2,082 Kenya GDP/ ZA GDP ratio adjustment: (2,082/7,055)= 0.295 |
| HIV-positive CD4 200 - 349 | 30.38 | 2021 | Eaton 2014 ^5^ | Adjusted for inflation and GDP/capita ratio (see steps above) |
| HIV-positive CD4 > 350 | 8.59 | 2021 | Eaton 2014 ^5^ | Adjusted for inflation and GDP/capita ratio (see steps above) |
| End of life care | 105.68 | 2021 | Eaton 2014 ^5^ | Adjusted for inflation and GDP/capita ratio (see steps above) |
| Annual ART provision costs ^6^ | 196.85 | 2020 | Long et al. (2010) ^6^ | 1st line ART delivery cost: $121 in 2016 USD → $131 in 2020 USD includes labs + staff encounters Cost of 1st line ART: $131 USD (delivery cost in 2020 USD) + $43.20 (ART)*1.2(additional 20% supply chain)=$ 182.84; The delivery cost ratio between 2nd and 1st ART is 2.4;  Cost of 2nd line ART: $131*2.4 USD (delivery cost in 2020 USD) + $279.60*1.2=$649.92; weighted average of 1st adn 2nd lines ART cost assuming 3% on 2nd line ART: $182.84*0.97 + $649.92*0.03=$196.85 |
| **Oral PrEP costs** |  |  |  |  |
| Oral PrEP per person month, facility | 10.88 | 2019 | Wanga et al. 2019 ^7^ | Data from micro-costing study, including viable (personnel, drug, lab and HIV testing, and other) and fixed (training, demand creation, personnel, capital, overhead) costs |
| Facility-based HIV-positive test | 3.68 | 2017 | Meisner et al. (2021) ^8^ | Data from micro-costing study. Inputs include screening test kit, other supply costs, and personnel costs. |
| Facility-based HIV-negative test | 2.64 | 2017 | Meisner et al. (2021) ^8^ | Data from micro-costing study. Inputs include screening test kit, other supply costs, and personnel costs. |
| **LA PrEP costs** |  |  |  |  |
| MK-8527 pill (monthly supply) | Low: $1.00, mid: $2.50, high: $5.00 |  |  | Assumption |
| LEN injection (bi-annual supply including accompanying oral dose) | $20.00 | 2025 |  | CHAI 2025^9^ |
| LEN loading dose | $12.75 | 2025 |  | CHAI 2025^9^ |
| Demand generation | 10% of LEN injection + loading dose price |  |  | Assumption (same for MK-8527 and LEN) |
| LEN Delivery | $8.55: $2.50 for HIV test, $0.05 for syringe, $6 for delivery overhead (facilities, staff, etc.) |  | Galactionova et al. (2015) ^10^  Ojal et al. (2019) ^11^  Mvundura et al. (2015) ^12^  Mangale et al. (2022) ^13^  UNICEF (2024) ^14^ | Syringe estimates were derived from UNICEF vaccine program estimates. Delivery overhead estimates were derived from the oral PrEP literature; provider time or overhead that is above and beyond resources needed to dispense oral PrEP are not included. |
| LEN Product wastage | 5% |  | Ojal et al. (2019) ^11^ |  |
| MK 8527 Delivery | $7.20 (overhead, personnel, HIV testing) |  | Wanga et al. (2021)^7^, Mangale et al. (2025)^15^, Bhardwaj (unpublished) | Obtained from oral PrEP provision costs minus oral PrEP drug costs and lab testing |
| MK 8527 Product wastage | 10% |  | Assumption | Assume higher wastage with pills |

###### Table S12 South Africa costing parameter calculations

| **South Africa** | |  | |  |  |  |
| --- | --- | --- | --- | --- | --- | --- |
| **Cost parameter** | | **Estimate (USD)** | | **Year** | **Data source** | **Calculation details, notes** |
| **HIV costs** | |  | |  |  |  |
| Annual health care costs  (among those not on ART) | |  | |  |  |  |
| HIV-positive CD4 < 200 | | 374.08 | | 2021 | Eaton 2014 ^5^ | Adjusted for inflation and GDP/capita ratio using this approach: Adjust South Africa value in 2012 USD for inflation to be in 2021 USD ($374.08 = $167*2.24)  Cost of health care use, CD4 count <200 cells per μL, not in HIV care (per person-year) in South Africa= $167 USD Inflation Rate between time of costing (2012) and 2021: 2.24 =4.7/2.1 |
| HIV-positive CD4 200 - 349 | | 102.95 | | 2021 | Eaton 2014 ^5^ | Adjusted for inflation and GDP/capita ratio (see above) |
| HIV-positive CD4 > 350 | | 29.10 | | 2021 | Eaton 2014 ^5^ | Adjusted for inflation and GDP/capita ratio (see above) |
| End of life care | | 358.10 | | 2021 | Eaton 2014 ^5^ | Adjust for inflation using World Bank CPI: the cost in 2021 USD for SA $160*4.7/2.1=$358.1 2021 USD. |
| Annual ART provision costs | | 189.56 | | 2020 | Long et al. (2010) ^6^ | 1st line ART delivery cost: $ 119 (including personnel, building, equipment, etc) in 2018 USD → $124 in 2020 USD  Cost of 1st line ART: $124 USD (delivery cost in 2020 USD) + $43.20 (ART)*1.2(additional 20% supply chain)=$ 175.84; The delivery cost ratio between 2nd and 1st ART is 2.4;  Cost of 2nd line ART: $124*2.4 USD (delivery cost in 2020 USD) + $279.60*1.2=$633.12 per year; weighted average of 1st and 2nd lines ART cost assuming 3% on 2nd line ART: $175.84*0.97 + $633.12*0.03=$189.56 |
| **Oral PrEP costs** | |  | |  |  |  |
| Oral PrEP per person month, facility | | 15.20 | | 2021 | Jamieson et al. (2022) ^16^  Jamieson et al. (2020) ^16^ | Ingredients-based approach, including rapid HIV testing, counselling, provision of condoms, syndromic screening with treatment referral, adherence counselling, traning, outreach, mobilization, monitoring and evaluation costs; Average cost of oral PrEP per person initiated:$76-78 across FSW, AGYW, and heterosexual men with a duration of 5 months use. This translates to a monthly cost of ~76/5=$15.20 |
| Facility-based HIV-positive test | | 5.62 | | 2018 | Meyer-Rath et al. (2019) ^17^ | Including supply, furniture, and staff salaries |
| Facility-based HIV-negative test | | 3.62 | | 2018 | Meyer-Rath et al. (2019) ^17^ | Including supply, furniture, and staff salaries |
| **LA PrEP costs** |  | |  | |  |  |
| MK-8527 pill (monthly supply) | Low: $1.00, mid: $2.50, high: $5.00 | |  | |  | Assumption |
| LEN injection (bi-annual supply including accompanying oral dose) | $20.00 | | 2025 | |  | CHAI 2025^9^ |
| LEN loading dose | $12.75 | | 2025 | |  | CHAI 2025^9^ |
| Demand generation | 10% of LEN injection + loading dose price | |  | |  | Assumption (same for MK-8527 and LEN) |
| LEN Delivery | $8.55: $2.50 for HIV test, $0.05 for syringe, $6 for delivery overhead (facilities, staff, etc.) | |  | | Galactionova et al. (2015) ^10^  Ojal et al. (2019) ^11^  Mvundura et al. (2015) ^12^  Mangale et al. (2022) ^13^  UNICEF (2024) ^14^ | Syringe estimates were derived from UNICEF vaccine program estimates. Delivery overhead estimates were derived from the oral PrEP literature; provider time or overhead that is above and beyond resources needed to dispense oral PrEP are not included. |
| LEN Product wastage | 5% | |  | | Ojal et al. (2019) ^11^ |  |
| MK 8527 Delivery | $7.20 (overhead, personnel, HIV testing) | |  | | Wanga et al. (2021)^7^, Mangale et al. (2025)^15^, Bhardwaj (unpublished) | Obtained from oral PrEP provision costs minus oral PrEP drug costs and lab testing |
| MK 8527 Product wastage | 10% | |  | | Assumption | Assume higher wastage with pills |

##### Total effective provision cost split by component over the course of LA PrEP (6 mo LEN, 3 mo MK)

##### S3a LA PrEP provision costs by component

|  | Drug price per course | Demand generation per course | Wastage per course | Delivery price per course |
| --- | --- | --- | --- | --- |
| LEN | 32.75 | 3.275 | 1.6375 | 8.55 |
| MK low | 3 | 1.6375 | 0.3 | 7.2 |
| MK mid | 7.5 | 1.6375 | 0.75 | 7.2 |
| MK high | 15 | 1.6375 | 1.5 | 7.2 |

##### S3b % Share of cost components for LA PrEP provision

|  | Drug price per course | Demand generation per course | Wastage per course | Delivery price per course |
| --- | --- | --- | --- | --- |
| LEN | 32.75 | 3.275 | 1.6375 | 8.55 |
| MK low | 3 | 1.6375 | 0.3 | 7.2 |
| MK mid | 7.5 | 1.6375 | 0.75 | 7.2 |
| MK high | 15 | 1.6375 | 1.5 | 7.2 |

##### Number of Oral PrEP initiations in Western Kenya and South Africa

###### Table S13 Number of PrEP initiations in Western Kenya and South Africa

|  | Western Kenya | South Africa |
| --- | --- | --- |
| 2016 | 655.2 | 722 |
| 2017 | 4376.4 | 3362 |
| 2018 | 13208.4 | 8476 |
| 2019 | 3370.8 | 45576 |
| 2020 | 11543.6 | 106402 |
| 2021 | 18007.2 | 205657 |
| 2022 | 68220.4 | 422239 |

Source: Global PrEP tracker, AVAC ^18,19^

##### FSW and male client of FSW estimation

The sizes of the FSW and male client populations in each setting were estimated based on Ministry of Health surveillance, FSW enumeration studies, and DHS data. Details of this process have been previously published paper (43). Briefly, we utilize primary data on mean and standard deviation age of FSWs and employ a 0 to 5 year delay from sexual debut to onset of female sex work using a Weibull distribution; based on data, we assume that FSWs engage in sex work for a for a mean of 5.4 years (95% CI 2 - 9 years), parameterized using a uniform distribution. Triangulating the number of females engaging in sex work at any one time and the duration of sex work, we estimated the lifetime probability that a female would engage in FSW by country. Similarly, we used DHS data on number of men reporting ever having paid for sex in their lifetime and in the last 12 months to inform estimates of ever being a male client of FSWs by setting.

###### Table S14: Lifetime probability of becoming a female sex worker or male client of FSW in western Kenya by county

|  | Homa Bay | Kisii | Kisumu | Migori | Nyamira | Siaya |
| --- | --- | --- | --- | --- | --- | --- |
| Prevalence of ever becoming FSW  (among females) | 0.0434 | 0.1302 | 0.14982 | 0.09936 | 0.05192 | 0.09406 |
| Prevalence of ever becoming a male client of FSWs (among males) | 0.063 | 0.2035 | 0.2341 | 0.1553 | 0.0811 | 0.147 |

###### Table S15: Lifetime probability of becoming a female sex worker or male client of FSW in South Africa

| South Africa (national) | |
| --- | --- |
| Prevalence of ever becoming FSW (among females) | 0.030 |
| Prevalence of ever becoming a male client of FSWs (among males) | 0.150 |

Data sources:

Odek WO, Githuka GN, Avery L, Njoroge PK, Kasonde L, Gorgens M, Kimani J, Gelmon L, Gakii G, Isac S, Faran E, Musyoki H, Maina W, Blanchard JF, Moses S. Estimating the size of the female sex worker population in Kenya to inform HIV prevention programming. PLoS One. 2014 Mar 3;9(3):e89180.

Fearon E, Chabata ST, Magutshwa S, Ndori-Mharadze T, Musemburi S, Chidawanyika H, Masendeke A, Napierala S, Gonese E, Herman Roloff A, et al. Estimating the Population Size of Female Sex Workers in Zimbabwe: Comparison of Estimates Obtained Using Different Methods in Twenty Sites and Development of a National-Level Estimate. J Acquir Immune Defic Syndr. 2020 Sep 1;85(1):30-38.

Konstant TL, Rangasami J, Stacey MJ, Stewart ML, Nogoduka C. Estimating the number of sex workers in South Africa: rapid population size estimation. AIDS Behav. 2015 Feb;19 Suppl 1:S3-15. doi: 10.1007/s10461-014-0981-y. PMID: 25582921.

South African Health Monitoring Survey (SAHMS): A Biological and Behavioural Survey among Female Sex Workers, South Africa 2018: Final Report. Accessed from: https://auruminstitute.org/images/Docs/SAHMS_II_FSW_BBS_Full_Report.pdf on August 29 2024.

South Africa Demographic and Health Survey. Accessed from: https://dhsprogram.com/publications/publication-fr337-dhs-final-reports.cfm on August 29 2024.

Kenya Demographic and Health Survey. Accessed from: https://dhsprogram.com/pubs/pdf/PR143/PR143.pdf on August 29 2024.

Kenya Demographic and Health Survey. Accessed from: https://dhsprogram.com/methodology/survey/survey-display-556.cfm August 29 2024.

### Supplemental Appendix Part II:

### Additional Results

#### Table S1: Five-year (2026-2030) budget impact analysis – MK = $1.00^*^

| Costs 2021 USD | ART | Oral PrEP | LEN | MK-8527 | Illness | Testing | Total |
| --- | --- | --- | --- | --- | --- | --- | --- |
| Western Kenya | | | | | | | |
| LEN only | $584,372 (-465,751 - 1,120,493) | -$343,150  (-393,085-  -301,838) | $46,199,705 (44,798,282 -47,883,774) | $- | -$545,595 (  -770,224-  -375,073) | $1,033,449 (899,449 – 1,202,895) | $46,928,781 (45,062,077- 48,471,427) |
| MK-8527 only | $1,022,220 (172,724 - 1,656,486) | $6,413 (-51,469 to 53,297) | $- | $19,545,164 (  18,949,364 -  20,178,355) | -$632,255 (  -841,884 to  -457,868) | $1,033,449 (899,449 – 1,202,895) | $20,974,991 (19,935,103 -  21,926,970) |
| MK-8527 + LEN | $719,203 (  -419,420 – 1,363,061) | -$410,329 (  -464,693 –  -365,442) | $51,752,568 (  50,094,830 -  53,372,683 ) | $5,825,436 (  5,650,183 -  6,026,006) | -$727,466 (  -934,226 –  -537,240) | $1,033,449 (899,449 – 1,202,895) | $58,192,861 (  56,077,518-  60,113,445) |
| Expanded MK-8527 + LEN | $681,212 (  -459,302 – 1,303,375) | -$410,921 (  -454,294 –  -363,123) | $51,480,968 (  49,745,440 -  53,214,907) | $12,931,391(  12,500,101 -  13,488,010) | -$758,684 (  -959,398 –  -601,596) | $1,033,449 (899,449 – 1,202,895) | $64,957,415 (62,058,301 – 66,899,042) |
| Expanded MK-8527 only | $1,137,564 (102,281 – 1,761,133) | $540 (-41,817 – 55,662) | $- | $31,415,618 (30,489,630 – 32,477,674) | -$790,501 (  -971,633 –  -606,242) | $1,033,449 (899,449 – 1,202,895) | $32,796,670 (30,940,988 – 34,348,378) |
| MK-8527 only at higher uptake | $1,258,588 (22,531 – 1,870,077) | $3,249 (  -52,056 – 40,676) | $- | $27,668,591 (26,819,471 – 28,505,704) | -$774,290 (-990,163 –  -620,736) | $1,033,449 (899,449 – 1,202,895) | $29,189,587 (27,682,012 – 30,449,491) |
| South Africa | | | | | | | |
| LEN only | -$18,460,905 (-29,811,719 –  -6,493,374) | -$4,510,266 (  -5,074,140 –  -3,898,231) | $241,026,855 (219,556,600 – 254,679,062) | $- | -$5,944,711 (  -15,726,170 –  3,581,794) | $28,375,324 (25,304,590 – 30,696,743) | $240,486,297 (221,797,906 - 263,738,286) |
| MK-8527 only | $28,121,340 (  -207,733,233 –  461,399,601) | $1,148,658 (  -537,924 –  5,848,673) | $- | $98,115,628 (89,722,162 – 104,049,876) | $13,129,618 (  -60,370,398 –  207,040,954) | $28,375,324 (25,304,590 – 30,696,743) | $168,890,568 (-94,685,016 - 765,896,295) |
| MK-8527 + LEN | -$19,517,613 (  -32,145,427 –  -10,785,412) | -$5,310,352 (-5,957,263 –  -4,716,033) | $262,486,188 (239,304,700 - 277,013,971) | $29,541,982 (26,752,093 –  31,253,246) | -$6,063,467 (  -15,496,083 -2,175,981) | $28,375,324 (25,304,590 – 30,696,743) | $289,512,062 (263,660,224 - 305,969,551) |
| Expanded MK-8527 + LEN | -$22,738,008 (  -34,278,803 –  -13,406,128) | -$5,333,848 (  -6,039,681 –  -4,732,160) | $261,203,994 (237,870,467 – 276,629,260) | $95,364,719 (87,996,874 –  100,664,763) | -$8,947,770 (  -16,557,485 – 426,863) | $28,375,324 (25,304,590 – 30,696,743) | $347,924,411 (319,480,490 - 370,271,405) |
| Expanded MK-8527 only | -$15,249,237 (  -25,733,589 –  -6,948,591) | $593,160 (  -118,555 – 1,218,966) | $- | $186,705,202 (172,054,865 – 196,309,669) | -$4,298,764 (  -11,500,986 –  4,228,103) | $28,375,324 (25,304,590 – 30,696,743) | $196,125,685 (180,388,856 - 210,525,927) |
| MK-8527 only at higher uptake | -$11,007,293 (  -24,845,580 – -3,121,896) | $704,485 (  -51,050 – 1,387,005) | $- | $136,439,581 (124,803,116 –  144,325,789) | -$3,701,944 (  -12,835,614 – 4,260,420) | $28,375,324 (25,304,590 – 30,696,743) | $150,810,153 (134,308,958 - 162,427,109) |

^*^Budget impact for each category is calculated as the total cost in the LA PrEP scenario minus the total cost in the standard of care scenario. Values in parathesis represent the 90% uncertainty interval across 100 simulations

#### Table S2: Five-year (2026-2030) budget impact analysis – MK = $2.50^*^

| Costs 2021 USD | ART | Oral PrEP | LEN | MK-8527 | Illness | Testing | Total |
| --- | --- | --- | --- | --- | --- | --- | --- |
| Western Kenya | | | | | | | |
| LEN only | $584,372 (-465,751 - 1,120,493) | -$343,150  (-393,085-  -301,838) | $46,199,705 (44,798,282 -47,883,774) | $- | -$545,595 (  -770,224-  -375,073) | $1,033,449 (899,449 – 1,202,895) | $46,928,781 (45,062,077 – 48,471,427) |
| MK-8527 only | $1,022,220 (172,724 - 1,656,486) | $6,413 (-51,469 to 53,297) | $- | $27,514,568 (  26,675,834 - 28,405,939 ) | -$632,255 (  -841,884 to  -457,868) | $1,033,449 (899,449 – 1,202,895) | $28,944,395 (27,674,019 – 30,228,913) |
| MK-8527 + LEN | $719,203 (  -419,420 – 1,363,061) | -$410,329 (  -464,693 –  -365,442) | $51,752,568 (  50,094,830 -  53,372,683 ) | $8,200,716 (  7,954,005 -  8,483,068 ) | -$727,466 (  -934,226 –  -537,240) | $1,033,449 (899,449 – 1,202,895) | $60,568,141 (58,398,850 – 62,543,938) |
| Expanded MK-8527 + LEN | $681,212 (  -459,302 – 1,303,375) | -$410,921 (  -454,294 –  -363,123) | $51,480,968 (  49,745,440 -  53,214,907) | $18,204,075 (17,596,930 - 18,987,652) | -$758,684 (  -959,398 –  -601,596) | $1,033,449 (899,449 – 1,202,895) | $70,230,099 (67,068,420 – 72,579,234) |
| Expanded MK-8527 only | $1,137,564 (102,281 – 1,761,133) | $540 (-41,817 – 55,662) | $- | $44,225,116 (42,921,563 – 45,720,218) | -$790,501 (  -971,633 –  -606,242) | $1,033,449 (899,449 – 1,202,895) | $45,606,168 (43,473,934 – 47,663,401) |
| MK-8527 only at higher uptake | $1,258,588 (22,531 – 1,870,077) | $3,249 (  -52,056 – 40,676) | $- | $38,950,265 (37,754,922 – 40,128,706) | -$774,290 (-990,163 –  -620,736) | $1,033,449 (899,449 – 1,202,895) | $40,471,261 (38,532,505 – 41,939,502) |
| South Africa | | | | | | | |
| LEN only | -$18,460,905 (-29,811,719 –  -6,493,374) | -$4,510,266 (  -5,074,140 –  -3,898,231) | $241,026,855 (219,556,600 – 254,679,062) | $- | -$5,944,711 (  -15,726,170 –  3,581,794) | $28,375,324 (25,304,590 – 30,696,743) | $240,486,297 (221,797,906 - 263,738,286) |
| MK-8527 only | $28,121,340 (  -207,733,233 –  461,399,601) | $1,148,658 (  -537,924 –  5,848,673) | $- | $138,121,589 (126,305,745 – 146,475,485) | $13,129,618 (  -60,370,398 –  207,040,954) | $28,375,324 (25,304,590 – 30,696,743) | $208,896,529 (-55,742,296 - 805,788,829) |
| MK-8527 + LEN | -$19,517,613 (  -32,145,427 –  -10,785,412) | -$5,310,352 (-5,957,263 –  -4,716,033) | $262,486,188 (239,304,700 - 277,013,971) | $41,587,518 (37,660,072 -43,996,539) | -$6,063,467 (  -15,496,083 -2,175,981) | $28,375,324 (25,304,590 – 30,696,743) | $301,557,598 (274,572,484 - 318,847,734) |
| Expanded MK-8527 + LEN | -$22,738,008 (  -34,278,803 –  -13,406,128) | -$5,333,848 (  -6,039,681 –  -4,732,160) | $261,203,994 (237,870,467 – 276,629,260) | $134,249,016 (123,876,984 – 141,710,115) | -$8,947,770 (  -16,557,485 – 426,863) | $28,375,324 (25,304,590 – 30,696,743) | $386,808,708 (356,041,921 - 409,337,965) |
| Expanded MK-8527 only | -$15,249,237 (  -25,733,589 –  -6,948,591) | $593,160 (  -118,555 – 1,218,966) | $- | $262,832,941 (242,209,032 – 276,353,563) | -$4,298,764 (  -11,500,986 –  4,228,103) | $28,375,324 (25,304,590 – 30,696,743) | $272,253,424 (251,800,206 - 291,441,793) |
| MK-8527 only at higher uptake | -$11,007,293 (  -24,845,580 – -3,121,896) | $704,485 (  -51,050 – 1,387,005) | $- | $192,071,865 175,690,712 – 203,173,619) | -$3,701,944 (  -12,835,614 – 4,260,420) | $28,375,324 (25,304,590 – 30,696,743) | $206,442,437 (187,318,786 - 220,252,557) |

^*^Budget impact for each category is calculated as the total cost in the LA PrEP scenario minus the total cost in the standard of care scenario. Values in parathesis represent the 90% uncertainty interval across 100 simulations

#### Table S3: Five-year (2026-2030) budget impact analysis – MK = $5.00^*^

| Costs 2021 USD | ART | Oral PrEP | LEN | MK-8527 | Illness | Testing | Total |
| --- | --- | --- | --- | --- | --- | --- | --- |
| Western Kenya | | | | | | | |
| LEN only | $584,372 (-465,751 - 1,120,493) | -$343,150  (-393,085-  -301,838) | $46,199,705 (44,798,282 -47,883,774) | $- | -$545,595 (  -770,224-  -375,073) | $1,033,449 (899,449 – 1,202,895) | $46,928,781 (45,062,077 – 48,471,427) |
| MK-8527 only | $1,022,220 (172,724 - 1,656,486) | $6,413 (-51,469 to 53,297) | $- | $40,796,907 (39,553,284 – 42,118,577) | -$632,255 (  -841,884 to  -457,868) | $1,033,449 (899,449 – 1,202,895) | $42,226,734 (40,648,568 -  43,760,121) |
| MK-8527 + LEN | $719,203 (  -419,420 – 1,363,061) | -$410,329 (  -464,693 –  -365,442) | $51,752,568 (  50,094,830 -  53,372,683 ) | $12,159,517 (11,793,709 – 12,578,171) | -$727,466 (  -934,226 –  -537,240) | $1,033,449 (899,449 – 1,202,895) | $64,526,942 (  62,260,172 -  66,594,758) |
| Expanded MK-8527 + LEN | $681,212 (  -459,302 – 1,303,375) | -$410,921 (  -454,294 –  -363,123) | $51,480,968 (  49,745,440 -  53,214,907) | $26,991,881 (26,091,644 – 28,153,721) | -$758,684 (  -959,398 –  -601,596) | $1,033,449 (899,449 – 1,202,895) | $79,017,905 (  75,574,918 -  82,046,221) |
| Expanded MK-8527 only | $1,137,564 (102,281 – 1,761,133) | $540 (-41,817 – 55,662) | $- | $65,574,280 (63,641,451 – 67,791,125) | -$790,501 (  -971,633 –  -606,242) | $1,033,449 (899,449 – 1,202,895) | $66,955,332 (  64,295,541 -  69,625,837) |
| MK-8527 only at higher uptake | $1,258,588 (22,531 – 1,870,077) | $3,249 (  -52,056 – 40,676) | $- | $57,753,056 (55,980,675 – 59,500,375) | -$774,290 (-990,163 –  -620,736) | $1,033,449 (899,449 – 1,202,895) | $59,274,052 (  56,511,509 -  61,105,934) |
| South Africa | | | | | | | |
| LEN only | -$18,460,905 (-29,811,719 –  -6,493,374) | -$4,510,266 (  -5,074,140 –  -3,898,231) | $241,026,855 (219,556,600 – 254,679,062) | $- | -$5,944,711 (  -15,726,170 –  3,581,794) | $28,375,324 (25,304,590 – 30,696,743) | $240,486,297 (221,797,906 - 263,738,286) |
| MK-8527 only | $28,121,340 (  -207,733,233 –  461,399,601) | $1,148,658 (  -537,924 –  5,848,673) | $- | $204,798,189 (187,278,384 – 217,184,833) | $13,129,618 (  -60,370,398 –  207,040,954) | $28,375,324 (25,304,590 – 30,696,743) | $275,573,129 (9,234,345 -  872,276,386) |
| MK-8527 + LEN | -$19,517,613 (  -32,145,427 –  -10,785,412) | -$5,310,352 (-5,957,263 –  -4,716,033) | $262,486,188 (239,304,700 - 277,013,971) | $61,663,412 (55,840,037 – 65,235,360) | -$6,063,467 (  -15,496,083 -2,175,981) | $28,375,324 (25,304,590 – 30,696,743) | $321,633,492 (292,915,956 - 340,311,373) |
| Expanded MK-8527 + LEN | -$22,738,008 (  -34,278,803 –  -13,406,128) | -$5,333,848 (  -6,039,681 –  -4,732,160) | $261,203,994 (237,870,467 – 276,629,260) | $199,056,177 (183,677,166 – 210,119,036) | -$8,947,770 (  -16,557,485 – 426,863) | $28,375,324 (25,304,590 – 30,696,743) | $451,615,869 (417,307,652 - 475,112,247) |
| Expanded MK-8527 only | -$15,249,237 (  -25,733,589 –  -6,948,591) | $593,160 (  -118,555 – 1,218,966) | $- | $389,712,506 (359,132,643 – 409,760,051) | -$4,298,764 (  -11,500,986 –  4,228,103) | $28,375,324 (25,304,590 – 30,696,743) | $399,132,989 (369,687,657 - 424,718,272) |
| MK-8527 only at higher uptake | -$11,007,293 (  -24,845,580 – -3,121,896) | $704,485 (  -51,050 – 1,387,005) | $- | $284,792,338 (260,503,373 – 301,253,335) | -$3,701,944 (  -12,835,614 – 4,260,420) | $28,375,324 (25,304,590 – 30,696,743) | $299,162,910 (274,005,660 - 316,933,574) |

^*^Budget impact for each category is calculated as the total cost in the LA PrEP scenario minus the total cost in the standard of care scenario. Values in parathesis represent the 90% uncertainty interval across 100 simulations

#### Table S4: Five-year (2026-2030) budget impact analysis – PEPFAR Interrupted scenario MK = $1.00^*^

| Costs 2021 USD | ART | Oral PrEP | LEN | MK-8527 | Illness | Testing | Total |
| --- | --- | --- | --- | --- | --- | --- | --- |
| Western Kenya | | | | | | | |
| LEN only | $5,243,039 (4,155,506 - 6,264,834) | -$346,453 (  -396,145 –  -300,710) | $45,627,175 (44,301,220 - 47,087,039) | $- | -$2,877,904 (  -3,328,366 –  -2,521,978) | $1,191,784 (1,033,835 – 1,388,001) | $48,837,641 (46,800,254 – 51,311,522) |
| MK-8527 only | $7,109,130 (5,898,666 - 8,387,071) | $9,065 (  -31,096 – 52,402) | $- | $19,242,463 (18,645,338 - 19,826,054) | -$3,312,985 (-3,800,733 –  -2,871,488) | $1,191,784 (1,033,835 – 1,388,001) | $24,239,457 (22,868,395 – 25,827,075) |
| MK-8527 + LEN | $6,965,872 (5,942,231 - 8,072,706) | -$417,075 (  -462,903 –  -368,940) | $51,069,338 (49,688,535 - 52,752,253) | $5,747,564 (5,539,924 - 5,922,891) | -$3,719,074 (-4,202,502 –  -3,323,961) | $1,191,784 (1,033,835 – 1,388,001) | $60,838,409 (58,562,914 -63,394,452) |
| Expanded MK-8527 + LEN | $7,207,391 (6,228,958 - 8,293,523) | -$412,882 (  -462,449 –  -360,968) | $50,790,658 (49,157,311 - 52,342,439) | $12,760,777 (12,261,945 - 13,369,999) | -$3,867,822 (  -4,341,604 –  -3,468,549) | $1,191,784 (1,033,835 – 1,388,001) | $67,669,906 (65,349,175 - 70,077,452) |
| Expanded MK-8527 only | $8,249,053 (7,085,987 - 9,439,087) | $8,281 (-42542 –53,779)_ | $- | $30,938,333 (30,036,974 - 31,975,411) | -$3,917,107 (  -4,400,780 –  -3,459,064) | $1,191,784 (1,033,835 – 1,388,001) | $36,470,344 (34,650,559 - 38,029,531) |
| MK-8527 only at higher uptake | $8,227,884 (9,951,491 - 12,821,437) | $9,193 (-32,822 – 56,787) | $- | $27,253,073 (26,401,144 - 28,048,938) | -$4,026,263 (  -4,461,410 –  -3,582,946) | $1,191,784 (1,033,835 – 1,388,001) | $32,655,671 (31,462,775 - 34,329,403 ) |
| South Africa | | | | | | | |
| LEN only | -$19,496,299 (-29,157,355 –  -7,119,397) | -$4,445,410 (  -5,260,895 –  -3,749,019) | $237,159,821 (215,478,514 – 250,918,361) | $- | -$12,379,882 (-21,249,114 – 1,762,983) | $30,065,699 (27,060,695 – 32,571,231) | $230,903,929 (206,673,849 - 254,534,060) |
| MK-8527 only | -$15,436,280 (  -20,974,007 –  1,885,942) | $588,274 (  -153,630 – 1,305,766) | $- | $96,011,783 (87,078,131 – 101,617,465) | -$9,969,226 (-15,781,912 – 4,733,505) | $30,065,699 (27,060,695 – 32,571,231) | $101,260,250 (95,722,724 - 122,134,316) |
| MK-8527 + LEN | -$24,699,565 (  -31,835,714 –  -12,127,859) | -$5,050,353 (  -5,852,216 –  -4,183,140) | $257,850,102 (233,186,981 – 273,052,051) | $29,039,815 (26,443,451 – 30,734,990) | -$14,569,281 (-23,934,419 – 1,954,319) | $30,065,699 (27,060,695 – 32,571,231) | $272,636,417 (247,303,554 - 296,216,046) |
| Expanded MK-8527 + LEN | -$28,238,833 (  -35,339,207 –  -17,698,124) | -$5,043,183 (  -5,762,689 –  -4,339,315) | $256,444,159 (233,159,572 – 271,965,164) | $93,681,318 (85,990,033 – 98,996,104) | -$17,249,863 (-24,844,396 – 5,956,359) | $30,065,699 (27,060,695 – 32,571,231) | $329,659,297 (  299,569,160 - 356,413,422) |
| Expanded MK-8527 only | -$18,746,692 (  -28,547,231 –  -5,412,188) | $718,544 (  -46,956 – 1,518,600) | $- | $182,838,094 (166,813,650 – 192,943,795) | -$10,004,319 (-18,431,337 – 1,971,202) | $30,065,699 (27,060,695 – 32,571,231) | $184,871,326 (167,224,166 - 202,893,861) |
| MK-8527 only at higher uptake | -$11,632,305 (  -25,736,962 –  -2,192,951) | $819,711 (  -1,424 – 1,448,597) | $- | $133,269,698 (120,690,085 – 140,811,982) | -$7,769,165 (-19,653,154 – 1,985,664) | $30,065,699 (27,060,695 – 32,571,231) | $144,753,638 (125,813,748 - 159,047,511) |

^*^Budget impact for each category is calculated as the total cost in the LA PrEP scenario minus the total cost in the standard of care scenario. Values in parathesis represent the 90% uncertainty interval across 100 simulations

#### Table S5: Five-year (2026-2030) budget impact analysis – Adherence to 1 out of 3 MK pills, MK = $1.00^*^

| Costs 2021 USD | ART | Oral PrEP | LEN | MK-8527 | Illness | Testing | Total |
| --- | --- | --- | --- | --- | --- | --- | --- |
| Western Kenya | | | | | | | |
| LEN only | $584,372 (-465,751 - 1,120,493) | -$343,150  (-393,085-  -301,838) | $46,199,705 (44,798,282 -47,883,774) | $- | -$545,595 (  -770,224-  -375,073) | $1,033,449 (899,449 – 1,202,895) | $46,928,781 (45,062,077 – 48,471,427) |
| MK-8527 only | $1,045,737 (77,967 – 1,677,222) | $3,383 (-46,122 – 51,436) | $- | $19,541,278 (18,974,016 – 20,195,339) | -$619,273 (  -806,250 –  -390,847) | $1,033,449 (899,449 – 1,202,895) | $21,464,371 (20,246,260 – 22,502,251) |
| MK-8527 + LEN | $1,047,350 (-429,335 – 1,317,681) | -$412,269 (  -454,553 –  -368,073) | $51,781,685 (50,194,800 – 53,456,353) | $5,826,886 (5,655,127 – 6,027,848) | -$723,258 (  -936,399 -  -546,046) | $1,033,449 (899,449 – 1,202,895) | $58,951,874 (56,490,599 – 60,814,161) |
| Expanded MK-8527 + LEN | $702,320 (  -442,315 – 1,469,034) | -$412,017 (  -463,960 –  -370,298) | $51,473,089 (49,846,961 – 53,183,800) | $12,922,044 (12,444,965 – 13,544,198) | -$756,571 (  -947,099 -  -572,575) | $1,033,449 (899,449 – 1,202,895) | $65,687,741 (63,052,215 – 68,006,112) |
| Expanded MK-8527 only | $1,231,923 (99,801 – 1,907,697) | $4,981 (-40,482 – 46,190) | $- | $31,412,278 (30,436,362 – 32,468,320) | -$774,308 (  -992,486 –  -611,613) | $1,033,449 (899,449 – 1,202,895) | $33,023,536 (31,334,856 – 34,483,834) |
| MK-8527 only at higher uptake | $1,314,992 (293,800 – 2,030,726) | $5,165 (-46,269 – 49,421) | $- | $27,678,912 (26,814,408 – 28,502870) | -$775,733 (  -951,898 –  -569,957) | $1,033,449 (899,449 – 1,202,895) | $28,933,136 (27,518,557 – 30,317,621) |
| South Africa | | | | | | | |
| LEN only | -$18,460,905 (  -29,811,719 –  -6,493,374) | -$4,510,266 (  -5,074,140 –  -3,898,231) | $241,026,855 (219,556,600 – 254,679,062) | $- | -$5,944,711 (  -15,726,170 –  3,581,794) | $28,375,324 (25,304,590 – 30,696,743) | $240,486,297 (221,797,906 - 263,738,286) |
| MK-8527 only | -$6,350,612 (  -20,446,066 –  -2,617,461) | $433,608 (  -367,905 – 982,338) | $- | $97,957,150 (89,530,041 – 103,588,232) | -$1,050,951 (-10,637,681 – 8,267,663) | $28,375,324 (25,304,590 – 30,696,743) | $119,364,519 (102,792,707 - 126,696,312) |
| MK-8527 + LEN | -$18,352,986 (  -30,152,202 –  -11,231,527) | -$5,352,160 (  -5,987,118 –  -4,732,906) | $262,399,859 (238,856,908 – 276,963,636) | $29,554,366 (26,930,033 – 31,329,985) | -$6,885,847 (  -16,996,066 – 610,045) | $28,375,324 (25,304,590 – 30,696,743) | $289,738,556 (263,801,010 - 307,732,124) |
| Expanded MK-8527 + LEN | -$22,034,677 (  -34,734,133 –  -12,606,340) | -$5,341,369 (  -5,889,080 –  -4,744,943) | $260,979,497 (238,137,690 -275,795,814) | $95,334,605 (87,700,832 – 101,016,448) | -$8,018,900 (-18,272,336 – 287,754) | $28,375,324 (25,304,590 – 30,696,743) | $349,294,480 (318,950,298 - 371,028,949) |
| Expanded MK-8527 only | -$8,644,684 (  -22,262,377 –  -3,663,472) | $480,020 (  -122,075 -982,819) | $- | $186,591,517 (171,557,361 – 196,203,690) | -$2,776,409 (  -11,243,627 – 3,916,216) | $28,375,324 (25,304,590 – 30,696,743) | $204,025,768 (181,241,373 - 214,196,335) |
| MK-8527 only at higher uptake | -$9,486,225 (  -23,479,171 –  -601,237) | $564,289 (  -54,315 – 1,120,259) | $- | $136,094,879 (124,042,990 – 143,823,234) | -$2,797,629 (-13,619,451– 5,759,987) | $28,375,324 (25,304,590 – 30,696,743) | $152,750,638 (136,793,805 - 163,290,808) |

^*^Budget impact for each category is calculated as the total cost in the LA PrEP scenario minus the total cost in the standard of care scenario. Values in parathesis represent the 90% uncertainty interval across 100 simulations

#### Table S6: Health and economic impact of LA PrEP scenarios – western Kenya

|  | LEN only | MK-8527 only | MK-8527 + LEN | Expanded MK-8527 + LEN | Expanded MK-8527 only | MK-8527 only at higher uptake |
| --- | --- | --- | --- | --- | --- | --- |
| Lenacapavir coverage | 2.4% (2.3 to 2.5) | - | 2.7% (2.6 to 2.8) | 2.7% (2.6 to 2.8) | - | - |
| MK coverage | - | 1.3% (1.3 to 1.4) | 0.4% (0.4 to 0.4) | 0.9% (0.9 to 0.9) | 2.2% (2.1 to 2.3) | 1.9% (1.8 to 1.9) |
| HIV infections averted (%) | 18.4% (14.8 to 22.8) | 13.9% (10.3 to 17.5) | 23.2% (19.3 to 27.3) | 24.8% (21.0 to 28.9) | 18.1% (13.7 to 22.6) | 17.2% (13.4 to 21.7) |
| HIV infections averted (n) | 9549 (6481 to 13046) | 7218 (4741 to 10489) | 12021 (8942 to 15570) | 12892 (10409 to 16402) | 9412 (6818 to 12780) | 8954 (6843 to 12285) |
| HIV-associated deaths averted (%) | 4.4% (1.4 to 9.1) | 4.2% (0.7 to 8.4) | 5.6% (2.3 to 8.7) | 5.6% (2.0 to 8.8) | 5.2% (1.8 to 8.6) | 5.0% (0.7 to 8.4) |
| HIV-associated deaths averted (n) | 1934 (584 to 4189) | 1874 (289 to 3694) | 2477 (923 to 3976) | 2472 (872 to 4115) | 2293 (772 to 3910) | 2213 (318 to 3883) |
| MK-8527 doses required | - | 1,439,565 (1,397,271 – 1,488,993) | 3,195,567 (3,089,742 – 3,333,012) | 4,829,943 (4,685,097 – 4,985,049) | 6,837,378 (6,632,718 – 7,043,646) | 7,763,331 (7,537,407 – 7,943,430) |
| Lenacapavir doses required | 999,777  (970,082 – 1,035,943) | 1,119,943  (1,084,771 – 1,154,824) | 1,114,066  (1,076,995 – 1,151,574) | - | - | - |
| 5-year budget impact (MK=$1.00) | $46,928,781 | $21,433,658 | $59,048,245 | $66,548,094 | $34,483,121 | $30,547,498 |
| 5-year budget impact (MK=$2.50) | $46,928,781 | $29,403,062 | $61,423,526 | $71,820,778 | $47,292,620 | $41,829,172 |
| 5-year budget impact (MK=$5.00) | $46,928,781 | $42,685,401 | $65,382,327 | $80,608,584 | $68,641,783 | $60,631,963 |
| 5-year LA PrEP drugs and wastage (MK=$1.00) | $34,382,339 | $5,312,936 | $40,098,361 | $41,827,836 | $8,539,666 | $7,521,116 |
| 5-year LA PrEP delivery and demand generation (MK=$1.00) | $11,817,367 | $14,232,228 | $17,479,642 | $22,584,523 | $22,875,953 | $20,147,475 |
| 5-year LA PrEP drugs and wastage (MK=$2.50) | $34,382,339 | $13,282,340 | $42,473,642 | $47,100,520 | $21,349,164 | $18,802,791 |
| 5-year LA PrEP delivery and demand generation (MK=$2.50) | $11,817,367 | $14,232,228 | $17,479,642 | $22,584,523 | $22,875,953 | $20,147,475 |
| 5-year LA PrEP drugs and wastage (MK=$5.00) | $34,382,339 | $26,564,679 | $46,432,443 | $55,888,327 | $42,698,328 | $37,605,581 |
| 5-year LA PrEP delivery and demand generation (MK=$5.00) | $11,817,367 | $14,232,228 | $17,479,642 | $22,584,523 | $22,875,953 | $20,147,475 |
| Modelled population size in 2026 | 4489080 (4480354 to 4495973) | | | | | |
| Median DALYs averted | 44288 (-39614 to 133729) | 46869 (-38717 to 127220) | 60808 (-23746 to 140454) | 55160 (-39068 to 133292) | 39928 (-50808 to 144123) | 40863 (-51216 to 130887) |
| ICER ($/DALY averted) at MK = $1.00 | $1306 (518 to 10697) | $467 (150 to 11778) | $1412 (658 to 9630) | $1542 (746 to 8699) | $887 (290 to 8424) | $748 (244 to 9144) |
| ICER ($/DALY averted) at MK = $2.50 | $1306 (518 to 10697) | $799 (316 to 18180) | $1489 (699 to 10191) | $1710 (830 to 9595) | $1436 (492 to 13542) | $1268 (431 to 14923) |
| ICER ($/DALY averted) at MK = $5.00 | $1306 (518 to 10697) | $1339 (577 to 28848) | $1613 (764 to 11126) | $1988 (971 to 11090) | $2348 (840 to 22072) | $2030 (782 to 24550) |
| Total costs averted over 35-year time horizon at MK = $1.00 | $74283709 (62659763 - 84852103) | $26057267 (16976270 - 35990931) | 93894984 (83174073 - 104561183) | 109321191 (98271579 - 119801327) | 50215412 (36573456 - 59864793) | 40498111 (32259506 - 49827512) |
| Total costs averted over 35-year time horizon at MK = $2.50 | 74283709 (62659763 - 84852103) | 43972315 (35263152 - 54044645) | 99169331 (88437149 - 109898179) | 121465335 (109903232 - 131951668) | 79587033 (65460750 - 89286835) | 66399956 (58092116 - 76419662) |
| Total costs averted over 35-year time horizon at MK = $5.00 | 74283709 (62659763 - 84852103) | 74778848 (65796561 - 84945151) | 108065555 (97208941 - 118793172) | 141588037 (129167041 - 151888200) | 130106388 (113606239 - 138564951) | 109907537 (101427332 - 120134235) |

DALY: Disability adjusted life year; ICER: incremental cost-effectiveness ratio; Health impacts are compared to the baseline scenario of daily oral pre-exposure prophylaxis only. HIV infections averted and long acting PrEP coverage are reported for people aged 15–65 years over 10 years of long-acting PrEP implementation; deaths averted are calculated over the 35-year time horizon. Values in parentheses show 90% uncertainty intervals representing the 5th and 95th percentiles across 100 parameter sets. Median ICER reported for non-dominated runs.

#### Table S7: Health and economic impact of LA PrEP scenarios – South Africa

|  | LEN only | MK-8527 only | MK-8527 + LEN | Expanded MK-8527 + LEN | Expanded MK-8527 only | MK-8527 only at higher uptake |
| --- | --- | --- | --- | --- | --- | --- |
| Lenacapavir coverage | 1.4% (1.3 to 1.5) | - | 1.5% (1.4 to 1.6) | 1.5% (1.4 to 1.6) | - | - |
| MK coverage | - | 0.7% (0.7 to 0.8) | 0.2% (0.2 to 0.3) | 0.7% (0.7 to 0.8) | 1.4% (1.3 to 1.5) | 1.1% (0.9 to 1.1) |
| HIV infections averted (%) | 10.7% (9.5 to 12.5) | 6.3% (4.5 to 7.8) | 13.2% (11.4 to 14.7) | 14.9% (13.4 to 16.4) | 9.3% (7.7 to 10.8) | 8.3% (6.5 to 10.2) |
| HIV infections averted (n) | 244364 (208967 to 281081) | 124425 (5808 to 182831) | 299938 (259427 to 337328) | 338866 (299234 to 378666) | 211850 (169181 to 252720) | 188958 (155520 to 229121) |
| HIV-associated deaths averted (%) | 2.5% (1.7 to 3.5) | 1.4% (0.4 to 2.3) | 3.1% (2.1 to 3.9) | 3.4% (2.3 to 4.4) | 2.1% (1.2 to 3.1) | 1.9% (0.9 to 2.6) |
| HIV-associated deaths averted (n) | 101154 (67430 to 148841) | 26750 (-309305 to 228854) | 124253 (85622 to 156191) | 139252 (92742 to 172082) | 85002 (46529 to 121047) | 76206 (35545 to 107632) |
| MK-8527 doses required | - | 7,300,326 (6,616,152 – 7,732,737) | 23,566,239 (21,750,912 – 24,974,679) | 24,246,036 (22,257,438 – 25,951,902) | 33,716,535 (30,871,443 – 35,826,219) | 46,138,023 (42,696,762 – 49,529,820) |
| Lenacapavir doses required | 5,215,903  (4,758,310 – 5,556,986) | 5,680,290  (5,183,193 – 6,025,272) | 5,652,543  (5,159,394 – 6,008,529) | - | - | - |
| 5-year budget impact (MK=$1.00) | $240,486,298 | $177,966,179 | $311,382,007 | $393,336,592 | $240,927,584 | $182,691,659 |
| 5-year budget impact (MK=$2.50) | $240,486,298 | $217,972,139 | $323,427,543 | $432,220,888 | $317,055,323 | $238,323,943 |
| 5-year budget impact (MK=$5.00) | $240,486,298 | $284,648,740 | $343,503,437 | $497,028,049 | $443,934,888 | $331,044,416 |
| 5-year LA PrEP drugs and wastage (MK=$1.00) | $179,374,888 | $26,670,640 | $203,375,521 | $220,313,805 | $50,751,826 | $37,088,189 |
| 5-year LA PrEP delivery and demand generation (MK=$1.00) | $61,651,968 | $71,444,988 | $88,652,649 | $136,254,909 | $135,953,376 | $99,351,392 |
| 5-year LA PrEP drugs and wastage (MK=$2.50) | $179,374,888 | $66,676,601 | $215,421,057 | $259,198,102 | $126,879,565 | $92,720,473 |
| 5-year LA PrEP delivery and demand generation (MK=$2.50) | $61,651,968 | $71,444,988 | $88,652,649 | $136,254,909 | $135,953,376 | $99,351,392 |
| 5-year LA PrEP drugs and wastage (MK=$5.00) | $179,374,888 | $133,353,201 | $235,496,951 | $324,005,262 | $253,759,130 | $185,440,946 |
| 5-year LA PrEP delivery and demand generation (MK=$5.00) | $61,651,968 | $71,444,988 | $88,652,649 | $136,254,909 | $135,953,376 | $99,351,392 |
| Modelled population size in 2026 | 42053495 (41952036 to 42140618) | | | | | |
| Median DALYs averted | 1063216 (582774 to 1575630) | 596784 (-4045619 to 3226212) | 1211756 (842546 to 1763895) | 1409592 (956461 to 1977848) | 881286 (311017 to 1411427) | 755629 (226815 to 1300021) |
| ICER ($/DALY averted) at MK = $1.00 | Cost-saving (100%) | Cost-saving (71%) | Cost-saving (100%) | Cost-saving (98%) | Cost-saving (92%) | Cost-saving (98%) |
| ICER ($/DALY averted) at MK = $2.50 | Cost-saving (100%) | Cost-saving (59%) | Cost-saving (99%) | Cost-saving (92%) | $118 (13 to 556) | Cost-saving (83%) |
| ICER ($/DALY averted) at MK = $5.00 | Cost-saving (100%) | $160 (9 to 1425) | Cost-saving (99%) | Cost-saving (52%) | $378 (125 to 1171) | $182 (19 to 923) |
| Total costs averted over 35-year time horizon at MK = $1.00 | -289312135 (-414995903 - -146508562) | -183906913 (-1014631052 - 2525164207) | -315744065 (-486032683 - -147688944) | -232383581 (-395109644 - -41892585) | -124463701 (-304135590 - 10965532) | -205666492 (-369450872 - -76633639) |
| Total costs averted over 35-year time horizon at MK = $2.50 | -289312135 (-414995903 - -146508562) | -92949578 (-928365354 - 2616488397) | -289369117 (-460440675 - -119774244) | -145928507 (-312104255 - 46196827) | 48786855 (-133822496 - 184760297) | -77465332 (-248284911 - 50888664) |
| Total costs averted over 35-year time horizon at MK = $5.00 | -289312135 (-414995903 - -146508562) | 55810648 (-784589191 - 2768695379) | -245410870 (-417401530 - -73249746) | -5485260 (-165967771 - 193820914) | 328462039 (136935100 - 476656267) | 129607387 (-48114923 - 267616931) |

DALY: Disability adjusted life year; ICER: incremental cost-effectiveness ratio; Health impacts are compared to the baseline scenario of daily oral pre-exposure prophylaxis only. HIV infections averted and long acting PrEP coverage are reported for people aged 15–65 years over 10 years of long-acting PrEP implementation; deaths averted are calculated over the 35-year time horizon. Values in parentheses show 90% uncertainty intervals representing the 5th and 95th percentiles across 100 parameter sets. Median ICER reported for non-dominated runs.

#### Table S8: Health and economic impact of LA PrEP scenarios: PEPFAR interruption – western Kenya ^*^

|  | LEN only | MK-8527 only | MK-8527 + LEN | Expanded MK-8527 + LEN | Expanded MK-8527 only | MK-8527 only at higher uptake |
| --- | --- | --- | --- | --- | --- | --- |
| Lenacapavir coverage | 2.4% (2.3 to 2.5) | - | 2.7% (2.6 to 2.8) | 2.7% (2.6 to 2.8) | - | - |
| MK coverage | - | 1.3% (1.3 to 1.4) | 0.4% (0.4 to 0.4) | 0.9% (0.9 to 0.9) | 2.1% (2.1 to 2.2) | 1.9% (1.8 to 1.9) |
| HIV infections averted (%) | 29.2% (26.6 to 31.6) | 25.4% (22.2 to 28.7) | 35.1% (32.1 to 37.4) | 36.0% (33.3 to 38.4) | 29.8% (27.7 to 32.8) | 29.7% (27.4 to 33.2) |
| HIV infections averted (n) | 36743 (31389 to 42137) | 31988 (26512 to 37757) | 44210 (37899 to 50640) | 45403 (38731 to 52145) | 37593 (31817 to 44662) | 37428 (31156 to 45506) |
| HIV-associated deaths averted (%) | 19.2% (17.2 to 21.5) | 18.8% (16.3 to 21.2) | 22.3% (20.0 to 24.8) | 22.7% (20.6 to 24.6) | 20.9% (19.0 to 23.1) | 20.7% (18.1 to 23.2) |
| HIV-associated deaths averted (n) | 47003 (39182 to 55754) | 45905 (37889 to 53368) | 54579 (47429 to 63412) | 55440 (46272 to 64909) | 51202 (43873 to 61285) | 50590 (41497 to 61365) |
| 5-year budget impact (MK=$1.00) | $48,837,641 | $24,774,367 | $61,803,208 | $69,400,875 | $38,341,818 | $34,199,295 |
| 5-year budget impact (MK=$2.50) | $48,837,641 | $32,620,347 | $64,146,737 | $74,603,993 | $50,956,707 | $45,311,545 |
| 5-year budget impact (MK=$5.00) | $48,837,641 | $45,696,980 | $68,052,619 | $83,275,855 | $71,981,521 | $63,831,961 |
| 5-year LA PrEP drugs and wastage (MK=$1.00) | $33,956,255 | $5,230,653 | $39,568,726 | $41,267,722 | $8,409,926 | $7,408,166 |
| 5-year LA PrEP delivery and demand generation (MK=$1.00) | $11,670,920 | $14,011,810 | $17,248,176 | $22,283,713 | $22,528,407 | $19,844,906 |
| 5-year LA PrEP drugs and wastage (MK=$2.50) | $33,956,255 | $13,076,633 | $41,912,255 | $46,470,840 | $21,024,814 | $18,520,416 |
| 5-year LA PrEP delivery and demand generation (MK=$2.50) | $11,670,920 | $14,011,810 | $17,248,176 | $22,283,713 | $22,528,407 | $19,844,906 |
| 5-year LA PrEP drugs and wastage (MK=$5.00) | $33,956,255 | $26,153,266 | $45,818,137 | $55,142,702 | $42,049,629 | $37,040,832 |
| 5-year LA PrEP delivery and demand generation (MK=$5.00) | $11,670,920 | $14,011,810 | $17,248,176 | $22,283,713 | $22,528,407 | $19,844,906 |
| Modelled population size in 2026 | 4489080 (4480354 to 4495973) | | | | | |
| Median DALYs averted | 478027 (379090 to 569919) | 469906 (384865 to 580283) | 552303 (447859 to 652258) | 577905 (462323 to 684264) | 525014 (412071 to 642224) | 539440 (447171 to 622594) |
| ICER ($/DALY averted) at MK = $1.00 | $31 (4 to 68) | Cost-saving (97%) | $40 (5 to 83) | $67 (23 to 109) | Cost-saving (73%) | Cost-saving (96%) |
| ICER ($/DALY averted) at MK = $2.50 | $31 (4 to 68) | Cost-saving (76%) | $45 (6 to 93) | $90 (40 to 133) | $44 (9 to 89) | $22 (2 to 52) |
| ICER ($/DALY averted) at MK = $5.00 | $31 (4 to 68) | $55 (12 to 107) | $62 (19 to 112) | $122 (67 to 175) | $137 (84 to 198) | $95 (40 to 132) |
| Total costs averted over 35-year time horizon at MK = $1.00 | 10356535 (-8507637 - 27802373) | -24125090 (-46228224 - -5087643) | 19376400 (-3101607 - 39868184) | 35495536 (13996526 - 54777482) | -7085205 (-26323970 - 11041664) | -17876969 (-43181812 - -793755) |
| Total costs averted over 35-year time horizon at MK = $2.50 | 10356535 (-8507637 - 27802373) | -5915870 (-28350810 - 12528733) | 24495732 (2093638 - 45152928) | 47343365 (25922824 - 66500566) | 22058605 (2663545 - 40551619) | 7608226 (-17776509 - 24829011) |
| Total costs averted over 35-year time horizon at MK = $5.00 | 10356535 (-8507637 - 27802373) | 24679432 (1738186 - 41889360) | 33088849 (10800757 - 53952835) | 67541379 (45720066 - 86188616) | 70040580 (50384825 - 89508651) | 50313641 (24565661 - 67452033) |

DALY: Disability adjusted life year; ICER: incremental cost-effectiveness ratio; Health impacts are compared to the baseline scenario of daily oral pre-exposure prophylaxis only. HIV infections averted and long acting PrEP coverage are reported for people aged 15–65 years over 10 years of long-acting PrEP implementation; deaths averted are calculated over the 35-year time horizon. Values in parentheses show 90% uncertainty intervals representing the 5th and 95th percentiles across 100 parameter sets. Median ICER reported for non-dominated runs.

#### Table S9: Health and economic impact of LA PrEP scenarios: PEPFAR interruption – South Africa^*^

|  | LEN only | MK-8527 only | MK-8527 + LEN | Expanded MK-8527 + LEN | Expanded MK-8527 only | MK-8527 only at higher uptake |
| --- | --- | --- | --- | --- | --- | --- |
| Lenacapavir coverage | 1.4% (1.3 to 1.5) | - | 1.5% (1.4 to 1.6) | 1.5% (1.4 to 1.6) | - | - |
| MK coverage | - | 0.7% (0.7 to 0.8) | 0.2% (0.2 to 0.3) | 0.7% (0.7 to 0.7) | 1.4% (1.3 to 1.5) | 1.0% (0.9 to 1.1) |
| HIV infections averted (%) | 10.6% (9.3 to 11.9) | 5.9% (4.6 to 7.5) | 13.1% (11.8 to 14.6) | 14.8% (13.3 to 16.3) | 8.8% (7.4 to 10.2) | 8.0% (6.6 to 9.5) |
| HIV infections averted (n) | 315054 (271009 to 363647) | 175841 (142708 to 220326) | 388740 (345011 to 435750) | 441271 (390317 to 489733) | 261280 (299759 to 219301) | 238263 (199368 to 277163) |
| HIV-associated deaths averted (%) | 3.1% (2.2 to 4.0) | 1.6% (0.8 to 2.5) | 3.8% (3.1 to 4.6) | 4.2% (3.3 to 5.0) | 2.4% (1.6 to 3.2) | 2.3% (1.5 to 3.2) |
| HIV-associated deaths averted (n) | 184922 (133409 to 239682) | 97596 (49363 to 154009 | 228448 (187894 to 287455) | 255989 (203501 to 313202) | 143846 (93674 to 205229) | 140331 (93317 to 193673) |
| 5-year budget impact (MK=$1.00) | $230,903,929 | $111,742,266 | $295,548,802 | $376,881,699 | $231,676,397 | $178,213,376 |
| 5-year budget impact (MK=$2.50) | $230,903,929 | $150,890,399 | $307,389,583 | $415,079,601 | $306,227,350 | $232,553,162 |
| 5-year budget impact (MK=$5.00) | $230,903,929 | $216,137,286 | $327,124,218 | $478,742,770 | $430,478,938 | $323,119,472 |
| 5-year LA PrEP drugs and wastage (MK=$1.00) | $176,496,997 | $26,098,755 | $199,788,790 | $216,313,885 | $49,700,635 | $36,226,524 |
| 5-year LA PrEP delivery and demand generation (MK=$1.00) | $60,662,824 | $69,913,028 | $87,101,126 | $133,811,591 | $133,137,459 | $97,043,174 |
| 5-year LA PrEP drugs and wastage (MK=$2.50) | $176,496,997 | $65,246,887 | $211,629,571 | $254,511,787 | $124,251,588 | $90,566,310 |
| 5-year LA PrEP delivery and demand generation (MK=$2.50) | $60,662,824 | $69,913,028 | $87,101,126 | $133,811,591 | $133,137,459 | $97,043,174 |
| 5-year LA PrEP drugs and wastage (MK=$5.00) | $176,496,997 | $130,493,774 | $231,364,207 | $318,174,956 | $248,503,176 | $181,132,621 |
| 5-year LA PrEP delivery and demand generation (MK=$5.00) | $60,662,824 | $69,913,028 | $87,101,126 | $133,811,591 | $133,137,459 | $97,043,174 |
| Modelled population size in 2026 | 42053495 (41952036 to 42140618) | | | | | |
| Median DALYs averted | 1817578 (1310128 to 2376653) | 969653 (356604 to 1594372) | 2302078 (1706204 to 2916090) | 2536264 (1968060 to 3128253) | 1470998 (804707 to 2079227) | 1417395 (942409 to 2014663) |
| ICER ($/DALY averted) at MK = $1.00 | Cost-saving (100%) | Cost-saving (100%) | Cost-saving (100%) | Cost-saving (100%) | Cost-saving (100%) | Cost-saving (99%) |
| ICER ($/DALY averted) at MK = $2.50 | Cost-saving (100%) | Cost-saving (98%) | Cost-saving (100%) | Cost-saving (100%) | Cost-saving (85%) | Cost-saving (99%) |
| ICER ($/DALY averted) at MK = $5.00 | Cost-saving (100%) | Cost-saving (67%) | Cost-saving (100%) | Cost-saving (100%) | $148 (24 to 329) | Cost-saving (65%) |
| Total costs averted over 35-year time horizon at MK = $1.00 | -510867256 (-728738933 - -335989862) | -283459603 (-497650680 - -117996926) | -615713896 (-852684529 - -441566458) | -566702187 (-805006201 - -359231600) | -258057068 (-484577571 - -131327400) | -376508506 (-544600263 - -212953954) |
| Total costs averted over 35-year time horizon at MK = $2.50 | -510867256 (-728738933 - -335989862) | -195456637 (-416050255 - -29072691) | -591065941 (-826099100 - -414911785) | -485338670 (-722714221 - -271751643) | -89990127 (-325728080 - 39720236) | -256476328 (-421636932 - -88049114) |
| Total costs averted over 35-year time horizon at MK = $5.00 | -510867256 (-728738933 - -335989862) | -48785026 (-280049546 - 118910853) | -548852018 (-785695926 - -370487331) | -343942617 (-585673994 - -127201817) | 189267565 (-59095969 - 332744176) | -60355678 (-222107951 - 118025590) |

DALY: Disability adjusted life year; ICER: incremental cost-effectiveness ratio; Health impacts are compared to the baseline scenario of daily oral pre-exposure prophylaxis only. HIV infections averted and long acting PrEP coverage are reported for people aged 15–65 years over 10 years of long-acting PrEP implementation; deaths averted are calculated over the 35-year time horizon. Values in parentheses show 90% uncertainty intervals representing the 5th and 95th percentiles across 100 parameter sets. Median ICER reported for non-dominated runs.

#### Table S10: Health and economic impact of LA PrEP scenarios: Assuming adherence to one out of three MK-8527 pills – Western Kenya ^*^

|  | LEN only | MK-8527 only | MK-8527 + LEN | Expanded MK-8527 + LEN | Expanded MK-8527 only | MK-8527 only at higher uptake |
| --- | --- | --- | --- | --- | --- | --- |
| Lenacapavir coverage | 2.4% (2.3 to 2.5) | - | 2.7% (2.6 to 2.8) | 2.7% (2.6 to 2.8) | - | - |
| MK coverage | - | 0.7% (0.7 to 0.7) | 0.2% (0.2 to 0.2) | 0.4% (0.4 to 0.5) | 1.1% (1.1 to 1.1) | 1.0% (0.9 to 1.0) |
| HIV infections averted (%) | 18.4% (14.8 to 22.8) | 12.1% (8.5 to 16.4) | 23.1% (19.4 to 27.1) | 24.3% (19.8 to 29.1) | 16.0% (11.4 to 20.3) | 15.5% (11.4 to 20.0) |
| HIV infections averted (n) | 9549 (6481 to 13046) | 8070 (5574 to 11193) | 11976 (9483 to 14910) | 12634 (10117 to 16518) | 8287 (5641 to 11248) | 8070 (5574 to 11193) |
| HIV-associated deaths averted (%) | 4.4% (1.4 to 9.1) | 3.9% (0.3 to 8.0) | 5.6% (2.2 to 9.0) | 5.5% (1.7 to 8.7) | 4.9% (1.5 to 9.0) | 4.8% (1.1 to 8.5) |
| HIV-associated deaths averted (n) | 1934 (584 to 4189) | 2103 (500 to 3896) | 2460 (923 to 4149) | 2436 (732 to 3938) | 2178 (643 to 4200) | 2103 (500 to 3896) |
| MK-8527 doses required | - | 1,439,565 (1,397,271 – 1,488,993) | 3,195,567 (3,089,742 – 3,333,012) | 4,829,943 (4,685,097 – 4,985,049) | 6,837,378 (6,632,718 – 7,043,646) | 7,763,331 (7,537,407 – 7,943,430) |
| Lenacapavir doses required | 999,777  (970,082 – 1,035,943) | 1,119,943  (1,084,771 – 1,154,824) | 1,114,066  (1,076,995 – 1,151,574) | - | - | - |
| 5-year budget impact (MK=$1.00) | $46,928,781 | $21,460,754 | $59,412,003 | $66,545,236 | $34,600,314 | $30,626,088 |
| 5-year budget impact (MK=$2.50) | $46,928,781 | $29,428,573 | $61,787,875 | $71,814,109 | $47,408,451 | $41,911,971 |
| 5-year budget impact (MK=$5.00) | $46,928,781 | $42,708,272 | $65,747,662 | $80,595,565 | $68,755,345 | $60,721,775 |
| 5-year LA PrEP drugs and wastage (MK=$1.00) | $34,382,339 | $5,311,880 | $40,120,426 | $41,819,432 | $8,538,758 | $7,523,922 |
| 5-year LA PrEP delivery and demand generation (MK=$1.00) | $11,817,367 | $14,229,399 | $17,488,146 | $22,575,701 | $22,873,521 | $20,154,990 |
| 5-year LA PrEP drugs and wastage (MK=$2.50) | $34,382,339 | $13,279,699 | $42,496,298 | $47,088,305 | $21,346,894 | $18,809,804 |
| 5-year LA PrEP delivery and demand generation (MK=$2.50) | $11,817,367 | $14,229,399 | $17,488,146 | $22,575,701 | $22,873,521 | $20,154,990 |
| 5-year LA PrEP drugs and wastage (MK=$5.00) | $34,382,339 | $26,559,398 | $46,456,085 | $55,869,760 | $42,693,788 | $37,619,608 |
| 5-year LA PrEP delivery and demand generation (MK=$5.00) | $11,817,367 | $14,229,399 | $17,488,146 | $22,575,701 | $22,873,521 | $20,154,990 |
| Modelled population size in 2026 | 4489080 (4480354 to 4495973) | | | | | |
| Median DALYs averted | 44288 (-39614 to 133729) | 32371 (-46577 to 125670) | 54467 (-24816 to 126646) | 45436 (-23529 to 141353) | 45928 (-59015 to 128308) | 40155 (-44686 to 113910) |
| ICER ($/DALY averted) at MK = $1.00 | $1306 (518 to 10697) | $549 (177 to 4297) | $1621 (688 to 7422) | $1777 (782 to 13845) | $876 (419 to 3213) | $814 (329 to 9003) |
| ICER ($/DALY averted) at MK = $2.50 | $1306 (518 to 10697) | $864 (333 to 7000) | $1714 (733 to 7875) | $1977 (857 to 15307) | $1423 (650 to 4892) | $1288 (490 to 13890) |
| ICER ($/DALY averted) at MK = $5.00 | $1306 (518 to 10697) | $1374 (570 to 11509) | $1869 (808 to 8631) | $2311 (991 to 17744) | $2210 (1026 to 7805) | $2061 (796 to 21851) |
| Total costs averted over 35-year time horizon at MK = $1.00 | 74283709 (62659763 - 84852103) | 28653070 (20213624 - 39211911) | 93886114 (83161751 - 103936358) | 109175061 (96431325 - 122354031) | 53723009 (44467029 - 62579317) | 44494005 (33175387 - 54349744) |
| Total costs averted over 35-year time horizon at MK = $2.50 | 74283709 (62659763 - 84852103) | 47321963 (38620865 - 57468754) | 99281720 (88460400 - 109398152) | 121286641 (108173279 - 134401551) | 83589695 (73991788 - 92646157) | 70338902 (59019207 - 80935024) |
| Total costs averted over 35-year time horizon at MK = $5.00 | 74283709 (62659763 - 84852103) | 77891861 (69162646 - 88257516) | 108306322 (97291481 - 118618831) | 141223154 (127743201 - 154477930) | 132830467 (121994549 - 142706140) | 113880615 (102750024 - 125149668) |

DALY: Disability adjusted life year; ICER: incremental cost-effectiveness ratio; Health impacts are compared to the baseline scenario of daily oral pre-exposure prophylaxis only. HIV infections averted and long acting PrEP coverage are reported for people aged 15–65 years over 10 years of long-acting PrEP implementation; deaths averted are calculated over the 35-year time horizon. Values in parentheses show 90% uncertainty intervals representing the 5th and 95th percentiles across 100 parameter sets. Median ICER reported for non-dominated runs.

#### Table S11: Health and economic impact of LA PrEP scenarios: Assuming adherence to one out of three MK-8527 pills – South Africa^*^

|  | LEN only | MK-8527 only | MK-8527 + LEN | Expanded MK-8527 + LEN | Expanded MK-8527 only | MK-8527 only at higher uptake |
| --- | --- | --- | --- | --- | --- | --- |
| Lenacapavir coverage | 1.4% (1.3 to 1.5) | - | 1.5% (1.4 to 1.6) | 1.5% (1.4 to 1.6) | - | - |
| MK coverage | - | 0.4% (0.3 to 0.4) | 0.1% (0.1 to 0.1) | 0.4% (0.3 to 0.4) | 0.7% (0.7 to 0.8) | 0.5% (0.5 to 0.6) |
| HIV infections averted (%) | 10.7% (9.5 to 12.5) | 5.0% (3.3 to 6.6) | 12.9% (11.4 to 14.6) | 14.3% (12.7 to 16.0) | 7.3% (5.8 to 9.1) | 6.6% (5.3 to 8.2 |
| HIV infections averted (n) | 244364 (208967 to 281081) | 113005 (73543 to 152610) | 294380 (253455 to 337500) | 326222 (287714 to 367560) | 166735 (133553 to 208811) | 150526 (115638 to 190263) |
| HIV-associated deaths averted (%) | 2.5% (1.7 to 3.5) | 1.1% (0.2 to 2.0) | 3.1% (2.2 to 3.9) | 3.3% (2.5 to 4.3) | 1.6% (0.5 to 2.6) | 1.6% (0.7 to 2.3) |
| HIV-associated deaths averted (n) | 101154 (67430 to 148841) | 45624 (7579 to 76816) | 124230 (91697 to 155893) | 135035 (96714 to 177671) | 65294 (20653 to 104816) | 62740 (30240 to 94452) |
| MK-8527 doses required | - | 7,300,326 (6,616,152 – 7,732,737) | 23,566,239 (21,750,912 – 24,974,679) | 24,246,036 (22,257,438 – 25,951,902) | 33,716,535 (30,871,443 – 35,826,219) | 46,138,023 (42,696,762 – 49,529,820) |
| Lenacapavir doses required | 5,215,903  (4,758,310 – 5,556,986) | 5,680,290  (5,183,193 – 6,025,272) | 5,652,543  (5,159,394 – 6,008,529) | - | - | - |
| 5-year budget impact (MK=$1.00) | $240,486,298 | $129,164,285 | $311,628,948 | $394,752,942 | $249,095,585 | $184,894,465 |
| 5-year budget impact (MK=$2.50) | $240,486,298 | $169,105,626 | $323,679,533 | $433,624,960 | $325,176,969 | $240,386,199 |
| 5-year budget impact (MK=$5.00) | $240,486,298 | $235,674,530 | $343,763,843 | $498,411,656 | $451,979,277 | $332,872,422 |
| 5-year LA PrEP drugs and wastage (MK=$1.00) | $179,374,888 | $26,627,561 | $203,314,640 | $220,138,545 | $50,720,923 | $36,994,489 |
| 5-year LA PrEP delivery and demand generation (MK=$1.00) | $61,651,968 | $71,329,588 | $88,639,585 | $136,175,556 | $135,870,594 | $99,100,390 |
| 5-year LA PrEP drugs and wastage (MK=$2.50) | $179,374,888 | $66,568,903 | $215,365,226 | $259,010,563 | $126,802,308 | $92,486,223 |
| 5-year LA PrEP delivery and demand generation (MK=$2.50) | $61,651,968 | $71,329,588 | $88,639,585 | $136,175,556 | $135,870,594 | $99,100,390 |
| 5-year LA PrEP drugs and wastage (MK=$5.00) | $179,374,888 | $133,137,806 | $235,449,536 | $323,797,259 | $253,604,616 | $184,972,447 |
| 5-year LA PrEP delivery and demand generation (MK=$5.00) | $61,651,968 | $71,329,588 | $88,639,585 | $136,175,556 | $135,870,594 | $99,100,390 |
| Modelled population size in 2026 | 42053495 (41952036 to 42140618) | | | | | |
| Median DALYs averted | 1063216 (582774 to 1575630) | 491419 (-111438 to 945143) | 1224506 (758439 to 1800035) | 1350311 (895667 to 1908272) | 649298 (172489 to 1198507) | 604115 (168215 to 1109785) |
| ICER ($/DALY averted) at MK = $1.00 | Cost-saving (100%) | Cost-saving (76%) | Cost-saving (100%) | Cost-saving (95%) | $119 (11 to 644) | Cost-saving (82%) |
| ICER ($/DALY averted) at MK = $2.50 | Cost-saving (100%) | $152 (11 to 725) | Cost-saving (100%) | Cost-saving (87%) | $317 (71 to 1383) | $164 (13 to 669) |
| ICER ($/DALY averted) at MK = $5.00 | Cost-saving (100%) | $284 (26 to 1527) | Cost-saving (98%) | $68 (7 to 209) | $728 (270 to 2716) | $401 (101 to 1543) |
| Total costs averted over 35-year time horizon at MK = $1.00 | -289312135 (-414995903 - -146508562) | -96335079 (-238408180 - 87488566) | -309469715 (-462918790 - -114319109) | -187080935 (-376790737 - 4371508) | 29178404 (-194195302 - 155830985) | -106244896 (-257199168 - 76638754) |
| Total costs averted over 35-year time horizon at MK = $2.50 | -289312135 (-414995903 - -146508562) | -3374373 (-149097453 - 175648098) | -283163155 (-436927935 - -88823110) | -102332068 (-292916097 - 91481825) | 205175803 (-28574905 - 329037805) | 15882326 (-130227384 - 203656875) |
| Total costs averted over 35-year time horizon at MK = $5.00 | -289312135 (-414995903 - -146508562) | 151560136 (-3738251 - 329392995) | -241333705 (-391692370 - -45637210) | 38916042 (-150578098 - 241112772) | 481792036 (257357578 - 620130526) | 220389546 (81063339 - 419809194) |

DALY: Disability adjusted life year; ICER: incremental cost-effectiveness ratio; Health impacts are compared to the baseline scenario of daily oral pre-exposure prophylaxis only. HIV infections averted and long acting PrEP coverage are reported for people aged 15–65 years over 10 years of long-acting PrEP implementation; deaths averted are calculated over the 35-year time horizon. Values in parentheses show 90% uncertainty intervals representing the 5th and 95th percentiles across 100 parameter sets. Median ICER reported for non-dominated runs.
